# Alternative approaches to standard inpatient mental health care: development of a typology of service models

**DOI:** 10.1101/2023.12.13.23298812

**Authors:** Jessica Griffiths, Helen Baldwin, Jerusaa Vasikaran, Ruby Jarvis, Ramya Pillutla, Katherine R. K. Saunders, Ruth Cooper, Una Foye, Luke Sheridan Rains, Molly Lusted-Challen, Phoebe Barnett, Geoff Brennan, Steven Pryjmachuk, Karen Newbigging, Jo Lomani, Rachel Rowan Olive, Lizzie Mitchell, Patrick Nyikavaranda, Chris Lynch, Karen Persaud, Brynmor Lloyd-Evans, Alan Simpson, Sonia Johnson

**Affiliations:** Mental Health Policy Research Unit (MHPRU), Department of Health Services and Population Research (HSPR), Institute of Psychiatry, Psychology & Neuroscience, King’s College London, London, United Kingdom; Mental Health Policy Research Unit (MHPRU), Division of Psychiatry, University College London, London, United Kingdom; Independent Researcher; National Health Service (NHS) England, United Kingdom; Centre for Outcomes Research and Effectiveness, Research Department of Clinical, Educational, & Health Psychology, University College London, London, UK; National Collaborating Centre for Mental Health, Royal College of Psychiatrists, London, UK; Department of Health Services and Population Research (HSPR), Institute of Psychiatry, Psychology & Neuroscience, King’s College London, London, United Kingdom; Division of Nursing, Midwifery and Social Work, University of Manchester, Manchester, United Kingdom; School of Social Policy, University of Birmingham, Birmingham, United Kingdom; NIHR Mental Health Policy Research Unit Lived Experience Working Group, Division of Psychiatry, University College London, London, United Kingdom; Department of Primary Care & Public Health, Brighton & Sussex Medical School, University of Sussex, Brighton, United Kingdom; Camden and Islington NHS Foundation Trust, London, United Kingdom

**Keywords:** Inpatient mental health care alternatives, typology development, crisis care, community care

## Abstract

**Background:** Inpatient mental health care is a challenging component of the mental health services system, with frequent reports of negative and coercive experiences and doubts about its therapeutic value. As such, alternative approaches for individuals experiencing a mental health crisis are highly desirable. This research aimed to identify models which offer an alternative to standard inpatient mental health care across all age groups, both nationally and internationally, and to develop a typology for these alternative models.

**Methods:** A dual literature search and expert consultation research methodology was adopted to identify relevant models. Three typologies of models were developed according to age group and acuity, including: alternatives to standard acute inpatient services for adults; alternatives to longer-stay inpatient services for adults, including rehabilitation and forensic inpatient services; and alternatives to standard inpatient services for children and young people.

**Results:** We identified an array of service models in each typology, some in community settings, some hospital-based and some working across settings. Models varied greatly in characteristics, extent of implementation and supporting evidence.

**Conclusions:** Through this mapping exercise, we have developed three novel typologies of alternatives to standard inpatient care. A range of community-based, hospital-based and cross-setting approaches were identified. The identification of services providing inpatient care in a substantially different way to the standard suggests that some improvements could be provided within existing structures. Potential inequities in access to alternatives were identified for certain groups, such as people who are compulsorily detained, younger children and young people transitioning between children’s and adult services. These typologies can inform future description, evaluation and comparison of different service models. This research also yields some key considerations for the design, development and implementation of alternative mental health service models and service arrays.

## 1. Introduction

Inpatient mental health care is seen frequently as a problematic component of the mental health services system, both within the United Kingdom (UK) (1) and globally (2). Inpatient services aim to offer intensive support and treatment of all modalities. Acute inpatient services admit people in crisis; longer-term wards are usually intended to focus on people with a high level of continuing need and/or risk. Stays are overnight and can result from either voluntary admission or involuntary admission in accordance with national law. However, standard inpatient mental health care has been widely criticised due to high rates of restrictive practices (3,4), safety concerns, including risks of abuse (5,6), limited treatment choices (7) and strained staff-service user relationships (8–10). Inpatient mental health care is routinely reported as coercive, dominated by involuntary hospitalisations, overly reliant on medication use (11,12) and failing to meet the needs of minority ethnic groups (13–15). Service user critiques of hospitalisation highlight concerns about the potential for standard inpatient mental health care to violate service users’ human rights and freedoms (16).

Inpatient care is also costly; even though only 3% of people in England accessing mental health care in 2018/19 received inpatient mental health care, National Health Service (NHS) trusts in England still invest more in inpatient than community services (11). Involuntary admissions have risen for several decades in England and some other high-income countries (17). Thus, service user dissatisfaction and activism, concerns with justice and human rights, doubts about inpatient service effectiveness and cost pressures are among the drivers for the search for effective, cost-effective and acceptable alternatives to standard inpatient care.

In light of these challenges, the World Health Organisation has proposed substantial changes in global mental health systems to deliver care that is person-centred, rights-based, recovery-oriented, and addresses social determinants of health (18). The quest for effective alternatives to standard hospital admission dates back to at least the mid-20^th^ century, and has been a central concern in policy, service development and research for many decades, including both hospital-based approaches and community alternatives to both acute and long-term inpatient services (19,20). However, the availability of alternative models continues to vary greatly between and within countries, and innovative services are often small in scale, remain underequipped and underfunded (11).

Currently, there is no extended typology which identifies alternative approaches to inpatient mental health care internationally and across all settings and age groups. Such a typology has potential benefits to researchers, service planners and clinicians in increasing the extent to which alternatives may be systematically introduced in contexts where they are appropriate, evaluated and implemented. Including alternatives to inpatient care for children and young people (CYP) and in longer-term settings is also an advance on the focus of much previous literature focusing solely on alternatives to acute adult mental health care: very significant clinical, ethical and economic disadvantages have been identified both for hospital admissions for under 18s (21,22) and for longer inpatient stays (23), so these are also valuable targets for the development, implementation of alternatives to standard inpatient care.

The National Institute for Health and Care Research (NIHR) Mental Health Policy Research Unit (MHPRU) (24), a research team funded to deliver research evidence to inform mental health policy, has carried out this study following a request from policy makers in NHS England and the Department of Health and Social Care. We sought to inform development and testing of inpatient and community alternatives to standard inpatient services and of integrated and comprehensive catchment area acute care systems by identifying, mapping out and categorising alternatives nationally and internationally, across all age groups. We aimed to develop three typologies of alternative approaches to standard inpatient mental health care according to: adult acute inpatient alternatives; adult long-term rehabilitation and forensic inpatient alternatives; and inpatient alternatives for CYP.

## 2. Methods

### 2.1. Study design

This study’s methodology was informed by scoping review principles (25). We used two simultaneous approaches for gathering data: literature scoping and a call for evidence from international experts (Figure 1). Alongside this, an expert working group was established to inform the direction of the research and offer iterative consultation throughout. The working group comprised academics and researchers from the NIHR MHPRU with relevant experience in acute care, as well as clinical and professional experts (including psychiatrists, mental health nurses, clinical psychologists and social workers), lived experience researchers selected from the MHPRU’s Lived Experience Working Group (LEWG) who had experiences of different mental health difficulties and accessing different services, and policy maker representatives from NHS England.

**Figure 1.** A flow diagram showing the research methodology used for typology development.

**Figure 2 (see Additional File 4).** A typology of alternative service models for acute inpatient mental health care for adults.

**Figure 3 (see Additional File 5).** A typology of alternatives service models for long-term inpatient care for adults (including rehabilitation and forensic services).

**Figure 4 (see Additional File 6).** A typology of alternative service models for acute, long-term and forensic inpatient mental health care for children and young people.

### 2.2. Eligibility criteria

Our criteria for defining ‘alternatives to standard inpatient mental health care’ were adapted from those used in a previous study by members of our team (26). To be considered an alternative, each model had to aim to serve people who would otherwise be admitted to an acute psychiatric ward or receive longer-term inpatient psychiatric care (e.g., in secure or rehabilitation services), and meet **at least one** of the following criteria:

- Based outside of a hospital setting, including services that support people intensively at home, in day care settings or in community residential settings with the aim of reducing pressure on hospital services;
- Dedicated to delivering specialised care for a specific diagnostic or sociodemographic group;
- Have a fixed maximum length of stay;
- Have implemented a specific therapeutic model involving changes in the practice of more than one profession within a hospital or hospital alternative service, or that involves different types of workers in care;
- Have implemented a significant change in practice in the management of risk.

We included models of care which fitted these criteria regardless of age range or geographical location, but, in the interests of feasibility, limited our scope through the exclusion of services specialising in care for perinatal populations, people with drug and alcohol problems, autistic people and people with intellectual disabilities, people living with dementia or other organic conditions, and solely prison-based services.

#### 2.2.1. Literature searching

##### Academic database searching

We conducted a broad initial search of relevant systematic and non-systematic reviews across three academic databases (PubMed, PsycINFO via Ovid and the Cochrane Central Register of Systematic Reviews) from the date of database initiation to 19^th^ December 2022. Key words for inpatient alternatives and specific service models already identified by the working group were used (see Supplementary A, Additional File 1). We aimed to offer a broad scope of the literature at the review level and as such, the search strategy was thorough though not exhaustive. There were no limits on the type of source that could be included for extraction.

Database records were exported to EndNote and screened for relevance by two researchers (HB, JG). Given the interest in international models, non-English language documents were included and translated using Google Translate. Extracted information was checked by someone with a knowledge of the language.

##### *Grey* literature *searching*

The academic literature searching was supplemented by grey literature searching using Google and nine other grey literature databases (see Supplementary B, Additional File 1) identified by the expert working group. Grey literature searches were conducted by an independent researcher (RP) between 16^th^ December 2022 and 1^st^ February 2023.

##### Supplementary searches

Supplemental searches of academic databases and grey literature were conducted for specific service models where there were gaps or insufficient detail in the accrued resources.

#### 2.2.2. Expert consultation

The literature scoping workstream was supplemented with expert consultation. Minimal risk ethics was obtained from the King’s College London ethics committee (Approval Number: MRA- 22/23-34963).

The expert working group identified key international experts (see Acknowledgements) to be contacted with a call for information (see Supplementary C, Additional File 1). 120 experts (including health care professionals, academics, charity leaders, experts by experience, and policymakers) were asked via email to provide information relating to relevant models, any literature associated with these models, and recommendations for other experts to contact. Using this snowballing approach, a further 18 experts were contacted, resulting in a total of 138 experts being individually contacted. The call for information was also distributed to 26 specialist clinical, academic, charitable and lived experience networks operating internationally and identified by the expert working group.

The call for information was delivered to the identified experts via email, and responses were received either via video call or email. Monetary compensation (£50) was provided to contributors from the voluntary/third sector who provided information via video expert consultation.

#### 2.2.3. Data extraction and synthesis

All relevant models and services identified from the literature scoping were extracted in Microsoft Excel by one of the research team (HB, JG, RJ, JV, KS, RC, RP). This data extraction form was designed following consultation with the expert working group and piloted on a set of studies initially in case modifications were needed. A range of key model descriptor variables were extracted, including: setting; funding; location; brief description of the model; date of establishment; target population; occupational roles of staff involved in the model; typical duration of support; access routes; whether the service can accept compulsorily detained individuals; and the details of any associated literature.

Where identified, additional information was also extracted regarding quantitative service use outcomes, including inpatient admissions/readmissions; number of inpatient bed days; length of stay; and satisfaction with care (see Additional File 2). It should be noted that the evidence presented was not subjected to quality appraisal and is not exhaustive.

Extracted models were then synthesised into three separate typologies where they were broadly categorised according to their settings, approaches and target populations. A decision was made to stratify the typologies by age and target population, rather than presenting a single comprehensive typology. This was for ease of interpretation, given the large number of models included, and to facilitate comparisons across the different populations. The typology categories were iteratively developed through consultation with the expert working group. Where knowledge gaps in the expert working group were identified, additional external experts were consulted.

## 3. Results

### 3.1. Model identification and typology development

#### Literature searching and expert consultation

Database searches returned 4,253 studies and a total of 78 experts responded to our call for information, from a range of countries. Their professional backgrounds included: academics, health care professionals, clinical academics, experts by experience, policy makers, charity workers and service leaders (see Supplementary D, Additional File 1 for more detailed breakdown). Screening of database search results and literature recommendations from experts for eligibility, combined with supplementary searches, resulted in the inclusion of 449 relevant sources. These sources ranged from published peer-reviewed literature to policy documents, websites, books and videos.

#### Typology mapping

Identified models were categorised in three typology maps: i) alternatives to adult standard acute inpatient care, ii) alternatives to adult standard long-term inpatient care (including rehabilitation and forensic inpatient care), and iii) alternatives to standard inpatient care for CYP.

Models in each typology were broadly categorised into community-based alternatives, hospital-based alternatives or cross-setting approaches. Community-based approaches encompass models which are home-based or located within another community setting, rather than in a traditional hospital setting. Hospital-based models are offered within a hospital setting, including inpatient wards operating in a substantially different way to standard inpatient care according to our criteria. Finally, cross-setting approaches are broader frameworks or philosophies that can be implemented across different types of settings whilst maintaining their core principles and values and often take a systemic perspective. There is some overlap in models featuring in each of the three typologies.

A description of the typologies follows below. Table 1 provides a high-level overview of service model comparisons across the three typologies. A more in-depth description of the models and identified quantitative evidence is provided in Additional File 2 and Additional File 3.

**Table 1.**
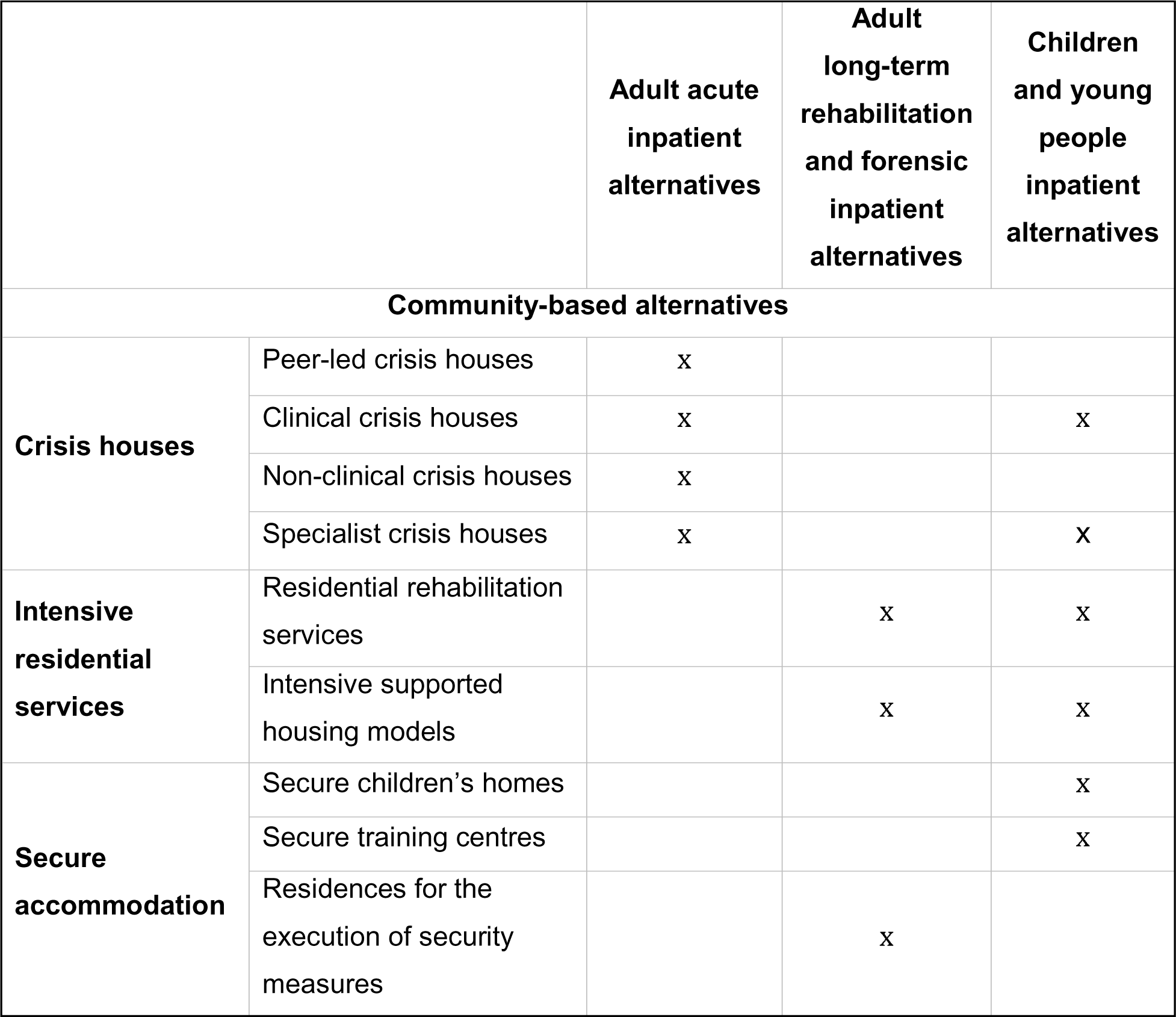
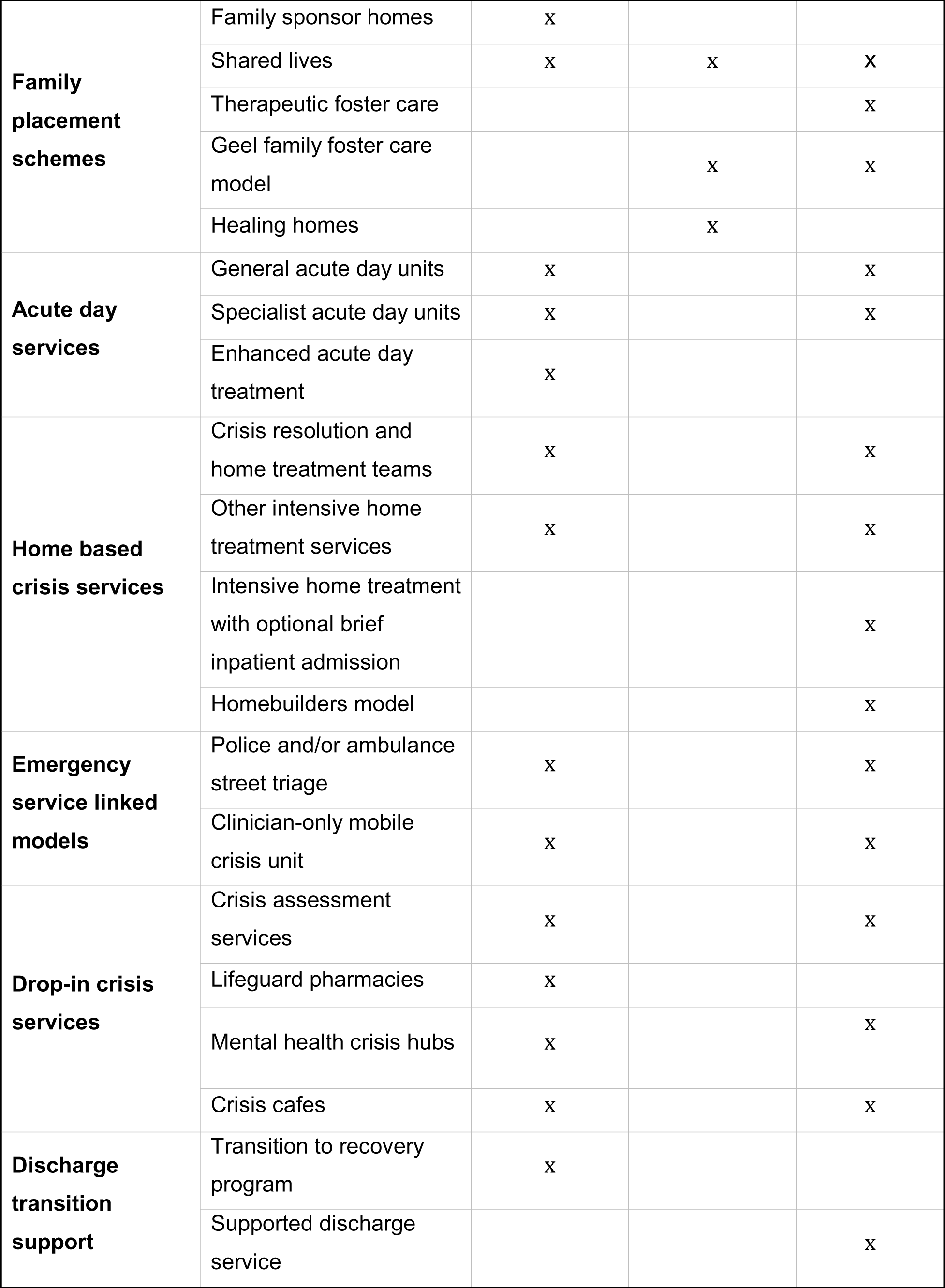
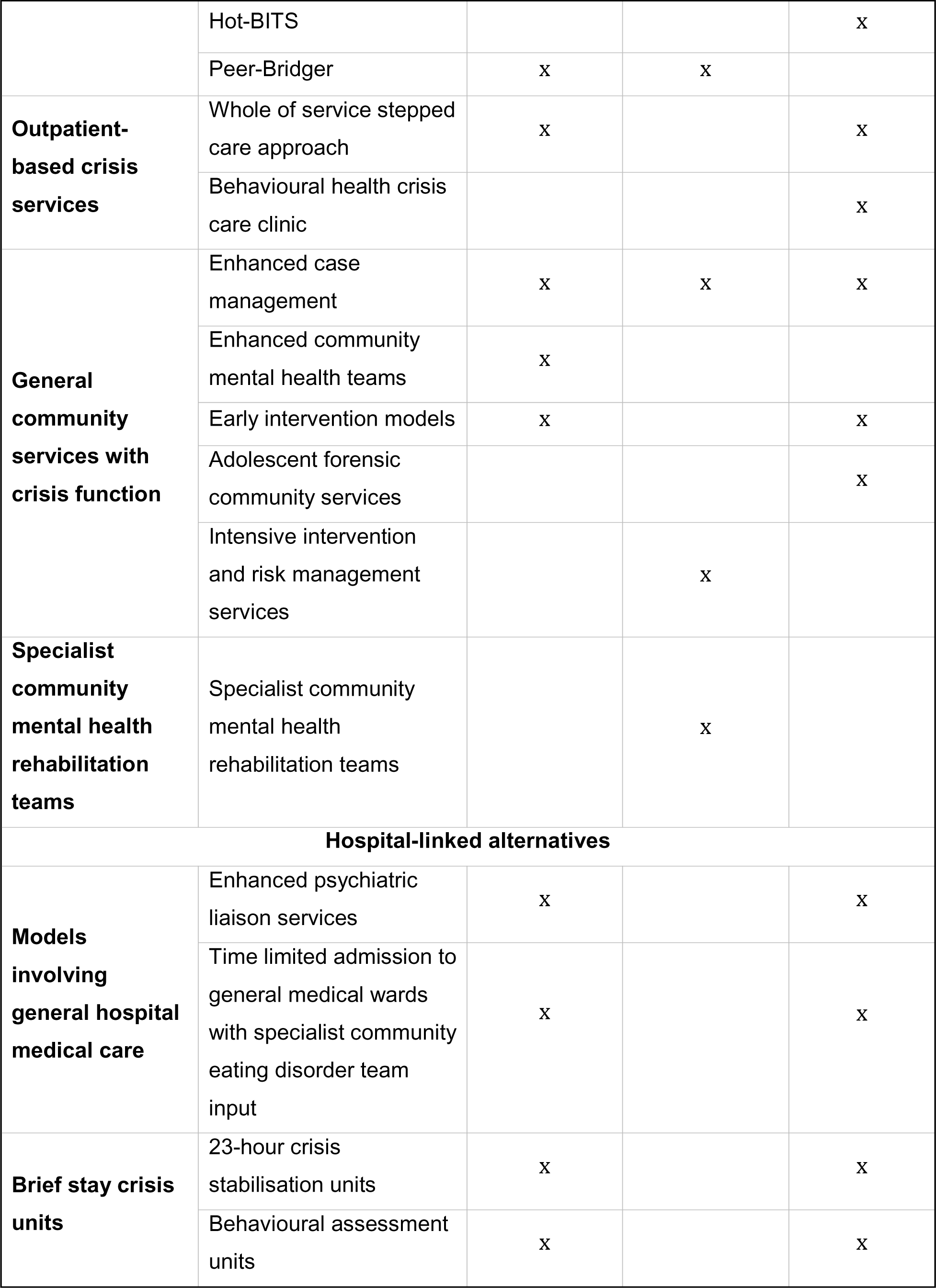
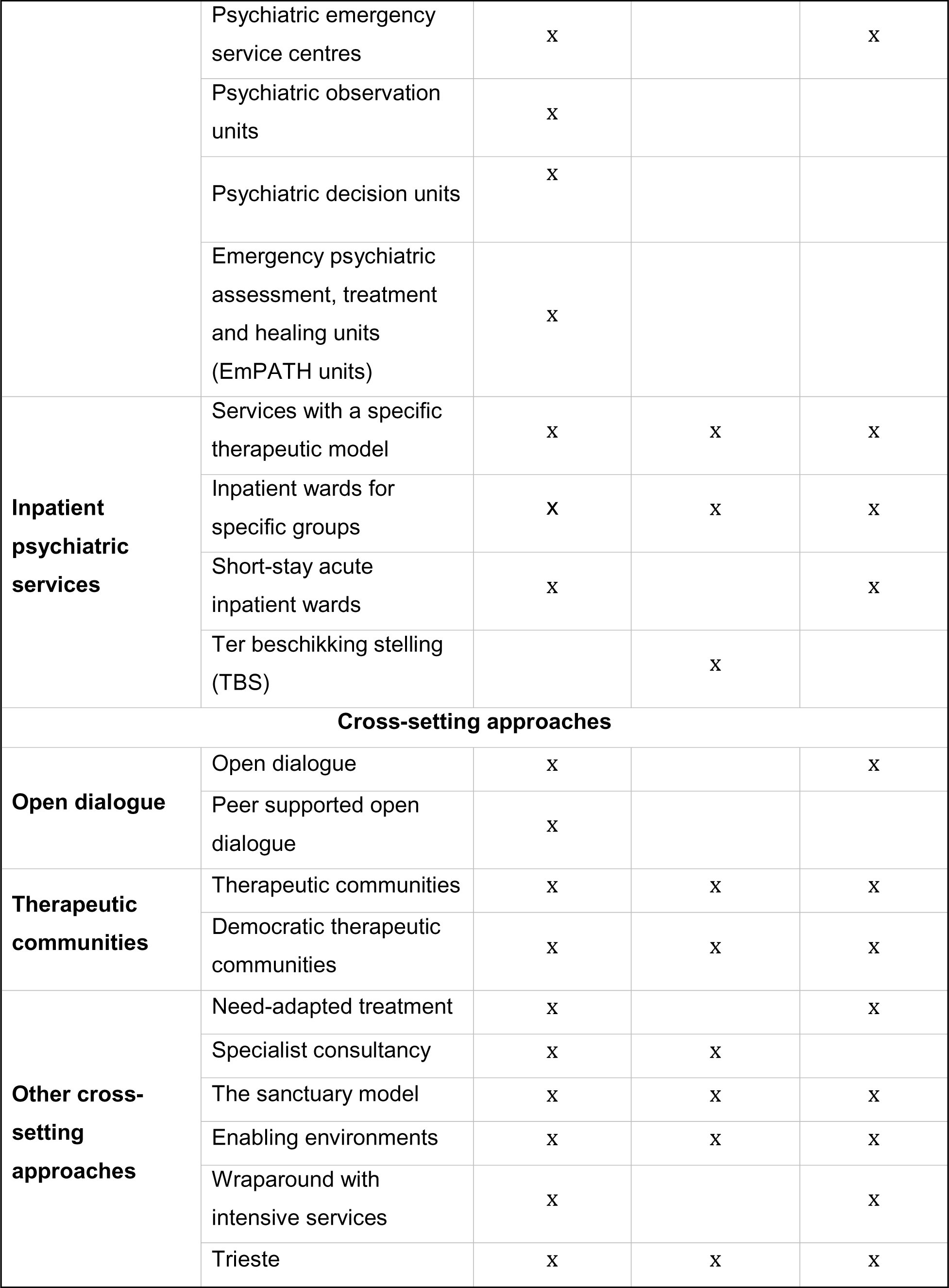
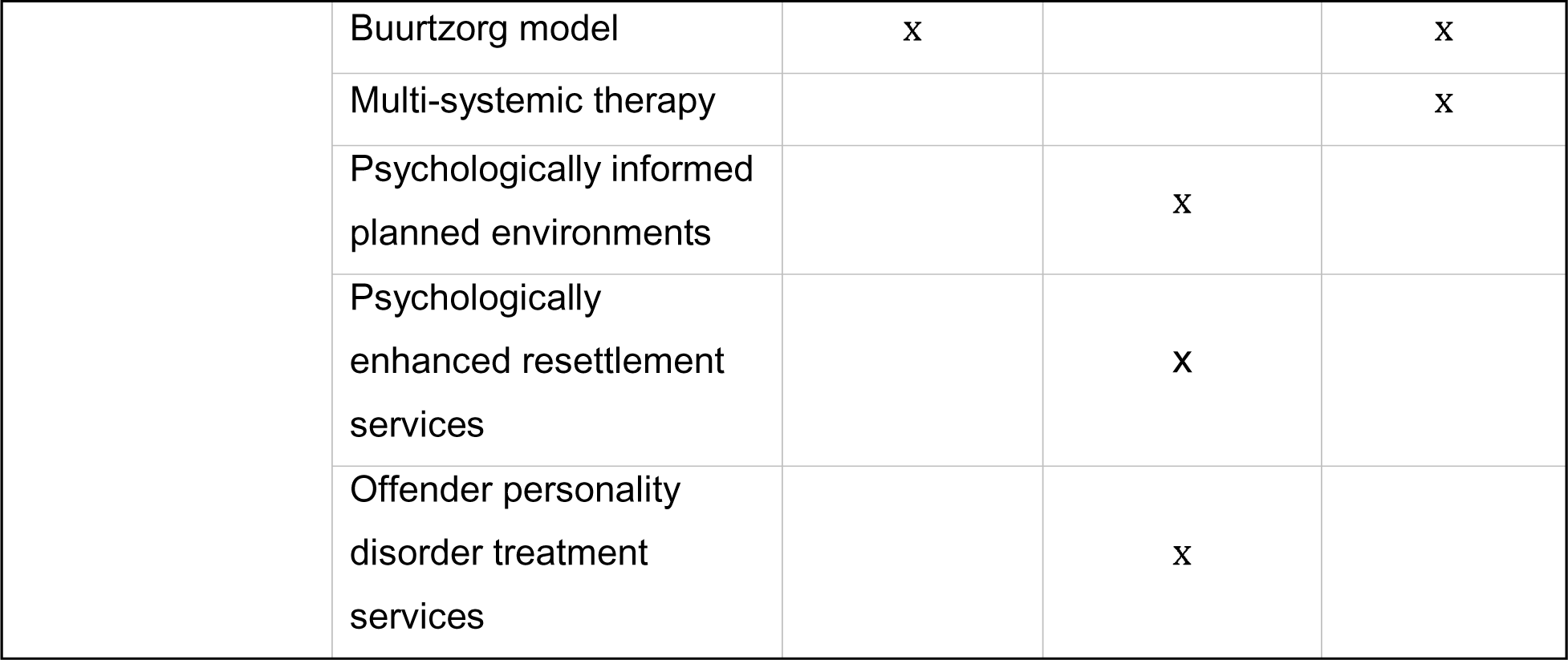
A high-level summary and comparison of the models of care across each of the typologies.

|  |  | Adult acute inpatient alternatives | Adult long-term rehabilitation and forensic inpatient alternatives | Children and young people inpatient alternatives |
| --- | --- | --- | --- | --- |
| **Community-based alternatives** |  |  |  |  |
| **Crisis houses** | Peer-led crisis houses | x |  |  |
|  | Clinical crisis houses | x |  | x |
|  | Non-clinical crisis houses | x |  |  |
|  | Specialist crisis houses | x |  | x |
| **Intensive residential services** | Residential rehabilitation services |  | x | x |
|  | Intensive supported housing models |  | x | x |
| **Secure accommodation** | Secure children's homes |  |  | x |
|  | Secure training centres |  |  | x |
|  | Residences for the execution of security measures |  | x |  |
| **Family placement schemes** | Family sponsor homes | x |  |  |
|  | Shared lives | x | x | x |
|  | Therapeutic foster care |  |  | x |
|  | Geel family foster care model |  | x | x |
|  | Healing homes |  | x |  |
| **Acute day services** | General acute day units | x |  | x |
|  | Specialist acute day units | x |  | x |
|  | Enhanced acute day treatment | x |  |  |
| **Home based crisis services** | Crisis resolution and home treatment teams | x |  | x |
|  | Other intensive home treatment services | x |  | x |
|  | Intensive home treatment with optional brief inpatient admission |  |  | x |
|  | Homebuilders model |  |  | x |
| **Emergency service linked models** | Police and/or ambulance street triage | x |  | x |
|  | Clinician-only mobile crisis unit | x |  | x |
| **Drop-in crisis services** | Crisis assessment services | x |  | x |
|  | Lifeguard pharmacies | x |  |  |
|  | Mental health crisis hubs | x |  | x |
|  | Crisis cafes | x |  | x |
| **Discharge transition support** | Transition to recovery program | x |  |  |
|  | Supported discharge service |  |  | x |
|  | Hot-BITS |  |  | X |
|  | Peer-Bridger | X | X |  |
| **Outpatient-based crisis services** | Whole of service stepped care approach | X |  | X |
|  | Behavioural health crisis care clinic |  |  | X |
| **General community services with crisis function** | Enhanced case management | X | X | X |
|  | Enhanced community mental health teams | X |  |  |
|  | Early intervention models | X |  | X |
|  | Adolescent forensic community services |  |  | X |
|  | Intensive intervention and risk management services |  | X |  |
| **Specialist community mental health rehabilitation teams** | Specialist community mental health rehabilitation teams |  | X |  |
| **Hospital-linked alternatives** |  |  |  |  |
| **Models involving general hospital medical care** | Enhanced psychiatric liaison services | X |  | X |
|  | Time limited admission to general medical wards with specialist community eating disorder team input | X |  | X |
| **Brief stay crisis units** | 23-hour crisis stabilisation units | X |  | X |
|  | Behavioural assessment units | X |  | X |
|  | Psychiatric emergency service centres | X |  | X |
|  | Psychiatric observation units | X |  |  |
|  | Psychiatric decision units | X |  |  |
|  | Emergency psychiatric assessment, treatment and healing units (EmPATH units) | X |  |  |
| **Inpatient psychiatric services** | Services with a specific therapeutic model | X | X | X |
|  | Inpatient wards for specific groups | X | X | X |
|  | Short-stay acute inpatient wards | X |  | X |
|  | Ter beschikking stelling (TBS) |  | X |  |
| **Cross-setting approaches** |  |  |  |  |
| **Open dialogue** | Open dialogue | X |  | X |
|  | Peer supported open dialogue | X |  |  |
| **Therapeutic communities** | Therapeutic communities | X | X | X |
|  | Democratic therapeutic communities | X | X | X |
| **Other cross-setting approaches** | Need-adapted treatment | X |  | X |
|  | Specialist consultancy | X | X |  |
|  | The sanctuary model | X | X | X |
|  | Enabling environments | X | X | X |
|  | Wraparound with intensive services | X |  | X |
|  | Trieste | X | X | X |
|  | Buurtzorg model | x |  | x |
|  | Multi-systemic therapy |  |  | x |
|  | Psychologically informed planned environments |  | x |  |
|  | Psychologically enhanced resettlement services |  | x |  |
|  | Offender personality disorder treatment services |  | x |  |

### 3.2. Adult acute inpatient alternatives

This section describes services intended to support adults in a mental health crisis that may otherwise result in admission to a standard acute inpatient ward (see Additional File 4). Further information, and associated evidence, for each model is provided in Additional File 2 and Additional File 3.

#### 3.2.1. Community-based alternatives

##### Crisis residential services

‘Crisis residential services’ provide temporary accommodation and support to people in crisis, typically for periods ranging from a few days to a few weeks. They include ‘Crisis houses’, which vary in approach, ranging from clinical to non-clinical. ‘Non-clinical crisis houses’ are usually run by voluntary sector organisations; they have fewer clinically qualified staff and employ approaches more clearly distinct from those in standard acute settings than ‘Clinical crisis houses’ (27). Crisis houses can be peer-led and can cater to specific sociodemographic groups, such as women or veterans, or specific clinical groups, such as people experiencing suicidality or early psychosis (e.g., Soteria houses) (27–31).

This category also includes ‘Family placement schemes’ (including ‘Family sponsor homes’ and ‘Shared Lives’) which place individuals with volunteer host families or carers at times of crisis. ‘Family sponsor homes’ support people for up to four weeks (32) whereas ‘Shared Lives’ can offer long-term, short-term or day-support placements (33).

##### Acute day units

‘Acute day units’ offer intensive, non-residential treatment and support for people facing acute mental health issues, offering an intermediate level of care between inpatient hospitalisation and outpatient services. ‘Enhanced acute day unit treatment’ models augment the standard model in some way, for example by providing additional respite or outreach services, crisis beds, or extended hours programmes (34). Acute day units tend to follow a structured schedule, normally operating during regular business hours (35). The intensity of programmes varies, and they tend to include a combination of individual therapy, group therapy, medication management, and support with practical issues (e.g., housing, welfare benefits, legal issues) and physical health (35). Some acute day units are designed for specific groups, such as people with diagnoses of eating disorders or “personality disorders” (36). ADUs may aim to avoid inpatient admissions or shorten inpatient stays (35). They can be part of step-down programmes, where people engage in brief intensive acute day unit treatment followed by a period of longer-term therapy (37,38).

##### Home-based crisis services

‘Home-based crisis models’ include ‘Crisis Resolution and Home Treatment Teams’ (CRHTTs) that provide intensive support and treatment to people in crisis, operate 24/7 and offer a rapid response (39). They often act as a gatekeeper for inpatient admissions (39). They are intended to avoid inpatient admissions and facilitate earlier discharge from hospital. They are staffed by multidisciplinary teams which can provide medication management, safety planning and psychological and social interventions (40). CRHTTs can provide both assessment and treatment, or these functions can be performed by separate teams. ‘Other intensive home treatment services’ can provide similar support, which may be staffed by psychiatrists, nurses, psychologists, social workers, therapists or peer workers (41–46).

##### Emergency service linked models

‘Paramedic and/or ambulance street triage teams’ are specialised units staffed by mental health clinicians and emergency service staff. Sometimes peer workers are also integrated into these teams. They provide immediate assessment and intervention to people in crisis who come into contact with law enforcement or emergency services in community settings (32,43,47). They aim to divert people from unnecessary arrest or incarceration. These teams can be ‘Police co-response teams’ or ‘Paramedic street triage teams’, and their formats can vary, with some providing in-person support from a mental health clinician during calls, and others where clinicians provide telephone advice only (32,43). ‘Clinician-only mobile crisis units’ perform similar functions but are only staffed by mental and physical health clinicians.

##### Drop-in crisis services

‘Drop-in crisis services’ provide immediate support to people experiencing a mental health crisis without the need for an appointment, as well as signposting and triage to further crisis support for those requiring urgent care. Some operate during extended hours, including evenings and weekends.

‘Crisis assessment services’ are designed to provide people in crisis with rapid assessment and referral on to other acute mental health services where deemed appropriate (32). ‘Lifeguard Pharmacies’ offer people experiencing suicidal thoughts and/or domestic abuse discreet consultations with pharmacists, who can signpost them to appropriate community support (48).

‘Mental health crisis hubs’ focus on assessment, stabilisation and connecting individuals to appropriate support, while ‘Crisis cafés’, which are usually voluntary sector run and can be peer-led, provide informal spaces for support and connection (32,49). These walk-in services are intended to divert people from hospital emergency departments, which can be potentially distressing and unsuitable locations for assessment for people in crisis, associated with a higher risk of inpatient admission (1,50).

##### Discharge transition support

‘Discharge transition support’ models aim to assist individuals with transitioning from inpatient care to the community, promoting recovery, independence, and reducing readmissions. The ‘Transition to Recovery Program’ includes psychosocial education, practical assistance, symptom management, social support and connecting individuals with appropriate services (51,52). The ‘Peer Bridger Project’ involves peer workers who offer goal setting, skills teaching, emotional support and advocacy through one-to-one interactions and support groups for an average of 12 months around the time of discharge from inpatient care (53).

##### Outpatient-based crisis services

The ‘Whole-of-service stepped-care’ model is a brief outpatient intervention approach for people with a “personality disorder” diagnosis. It offers immediate brief psychological intervention to people in crisis, followed by options for longer-term treatment (54).

##### Community services with crisis function

This category captures community-based mental health services which provide secondary mental health care generally and include some capacity for managing crises among people on their caseloads.

#### Enhanced community mental health teams

‘Enhanced community mental health teams’ (CMHT), also known as ‘extended hours’ teams, aim to proactively prevent crises and offer crisis intervention where needed, in addition to performing the standard ongoing case management functions typical of a standard CMHT (55).

#### Early intervention models

‘Early intervention models’ in mental health focus on specialist interventions, including detection, assessment, individualised treatment planning and psychosocial interventions, delivered by multidisciplinary teams. ‘Early intervention in psychosis’ (EIP) services support individuals aged 14-65 for up to three years after their first episode of psychosis. They offer assessment, pharmacological and psychological interventions, family support, employment/education assistance, crisis planning and help with social care issues, with the aim of reducing treatment delays, promoting recovery, avoiding hospitalisation and preventing relapse (56–58). Another early intervention model, ‘First episode and Rapid Early intervention for Eating Disorders’ (FREED), addresses eating disorders in young people aged 16-25, providing rapid access to specialised treatment and addressing unique needs related to this age group (59–63).

#### Enhanced case management models

‘Enhanced case management’ models for adults include ‘Assertive community treatment’ (ACT), ‘Intensive case management’ (ICM), ‘Flexible assertive community treatment’ (FACT) and ‘Peer-delivered case management’. They focus on long-term care coordination to ensure individuals receive appropriate services, treatment adherence support and crisis management. These models involve multidisciplinary teams with small caseloads, providing support in individuals’ home environments. In ACT, staff caseloads are shared whereas in ICM they are not. In FACT, care-coordinators manage individual caseloads but can also provide shared care at times of increased need (64,65). Peer-delivered case management involves peer workers alongside health care professionals (66–69).

### 3.2.2. Hospital-based alternatives

#### Models involving general hospital medical care

##### Enhanced psychiatric liaison services

‘Enhanced psychiatric liaison services’ aim to provide psychiatric assessment, treatment and support to people who are admitted to general hospitals or emergency departments with comorbid physical and mental health needs, including people in crisis. They ensure appropriate follow-up care post-discharge and are typically staffed by multidisciplinary teams. Enhanced models (including ‘Comprehensive’ and ‘Enhanced 24’ models e.g., ‘Rapid Assessment Interface Discharge’) offer more specialised care than standard psychiatric liaison services, increased psychiatric consultant input, and enhanced follow-up support (47,70–74), in part aiming to prevent avoidable inpatient admissions (73).

##### Time-limited admission to general medical wards with specialist community eating disorder team input

People with eating disorders may sometimes be admitted to general medical wards for medical stabilisation, with input from specialist community eating disorder teams. This approach is designed for people who have medical needs requiring inpatient care, including those who are too physically unwell for specialist eating disorder units (75,76).

#### Brief-stay crisis units

‘Brief-stay crisis units’ offer assessment, short-term treatment, and stabilisation for people in crisis outside of an inpatient setting (77). They are generally linked to hospital emergency departments. The environment is less restrictive than inpatient psychiatric units, and length of stay typically ranges from a few hours to a few days (77). They include ‘Psychiatric observation units’, ‘Psychiatric decision units’, ‘Emergency psychiatric assessment, treating and healing (EmPATH) units’, ‘Psychiatric emergency service centres’, ‘23-hour crisis stabilisation units’ and ‘Behavioural assessment units’.

#### Inpatient psychiatric services

##### Short-stay acute inpatient wards

‘Short-stay acute inpatient psychiatric wards’ provide assessment, stabilisation and intensive interventions for people requiring intensive psychiatric care for a limited period of time, usually ranging from a day up to a week, with the aim of facilitating discharge home within the admission time period wherever possible (27,32).

##### Inpatient psychiatric services with a specific therapeutic model

‘Safewards’, ‘Star Wards’, ‘Talk 1^st^, the ‘Tidal Model’, the ‘Bradford Refocusing Model’, ‘Six Core Strategies’ and the ‘HOPES model’ are specific therapeutic models aimed at improving care in inpatient mental health settings (27,78–90). They prioritise person-centred care, collaboration, prevention of crises and minimisation of risk. These models emphasise therapeutic relationships, recovery-oriented environments and staff training. ‘Drug-free and minimal medication wards’ in Norway also offer an alternative therapeutic approach (91).

‘Brief Admission’ (BA) and ‘Preventative Admission’ (PA) are two distinct models which have been implemented on standard psychiatric wards, which aim to proactively reduce crises and inpatient admissions through contracting admissions. The BA self-referral model allows individuals with emotional instability and/or a history of repeated self-harm to hospitalise themselves in a pre-negotiated way, in line with a contract created with clinicians at a time of non-crisis (92). In contrast, the PA model involves pre-arranged admissions over a longer period of time (93). As such, these two service models are distinct from ‘short stay acute inpatient wards’ (described above).

##### Inpatient wards for specific groups

Specialist inpatient services are designed to meet the needs of specific groups such as people who are deaf (94) or people with specific psychiatric diagnoses (e.g., eating disorders, “personality disorders”, early psychosis). Inpatient services for people diagnosed with a “personality disorder” employ therapies like dialectical behavioural therapy or mentalisation based treatment (95–97). Specialist eating disorder wards focus on medical stabilisation, nutritional rehabilitation, psychotherapy and developing effective coping skills (36,98). Early psychosis wards aim to provide timely intervention and comprehensive treatment (99,100).

#### 3.2.3. Cross-setting approaches

These approaches can be applied across a range of settings and stages in care pathways and include elements that are expected to reduce dependence on acute care.

‘The Sanctuary model’, ‘Enabling Environments’, ‘Therapeutic Communities’ and ‘Democratic Therapeutic Communities’ (DTCs) are approaches that aim to create supportive environments for people experiencing mental health difficulties, including those in crisis. The ‘Sanctuary Model’ focuses on creating trauma-informed environments (89,101), whilst ‘Therapeutic Communities’ are structured environments based on shared values such as attachment and respect, promoting recovery through social relationships and activities (102). DTCs are a type of therapeutic community based on the principles of democratic decision making, communal sharing, permissiveness with consequences, and reality confrontation (103,104). ‘Enabling Environments’ similarly aim to prioritise positive social environments for recovery and community integration (105). Each of these approaches can be applied in a range of settings, including acute inpatient settings.

‘Need-adapted treatment’ focuses on providing personalised care tailored to individuals’ needs, guided by a psychotherapeutic and family-centred approach (106). It can be applied across different contexts. Two examples of its implementation were identified - the ‘Swedish Parachute Project’ (107–109) and ‘Acute Psychosis Integrated Treatment’ (110,111).

‘Open Dialogue’, developed from need-adapted treatment, is a person-centred and network-centred approach to mental health care that involves systemic family therapy and psychodynamic principles. It emphasises open and transparent relationships between staff and service users as well as involving families and support networks, and there is a peer-supported variant (112,113).

‘Wraparound with intensive services’ similarly aims to engage multiple systems (e.g., health, social, education and youth justice services) to support and empower people aged 0-20 years old with complex needs and their families (114–116).

The ‘Trieste model’ in Italy and ‘Buurtzorg model’ from the Netherlands are whole-system approaches to organising care. The ‘Trieste model’ is a community-based approach to mental health care. It prioritises human rights, social inclusion and recovery. There are no locked doors and restrictive care is avoided (117,118). The ‘Buurtzorg model’ involves self-managed nursing teams providing holistic and person-centred care. Specialised ‘Buurtzorg T’ teams, which include mental health professionals, implement the model in the treatment of psychiatric patients (119,120).

Finally, there are a number of ‘Specialist consultancy’ models in operation that aim to promote alternative approaches to standard inpatient care and reduce inpatient admissions by offering consultation to relevant organisations. This consultancy may be provided by survivor-led, not-for-profit, statutory or private organisations (121–126).

### 3.3. Adult long-term inpatient care alternatives

Along with the array of services we identified which aim to offer support in times of acute crisis, service models were also identified that offer an alternative to standard long-term term hospital wards, including inpatient rehabilitation or forensic mental health care (see Additional File 5). These services largely aim to reduce the length of hospitalisation, support people to reside in the community, or offer a therapeutic approach that differs substantially from standard long-term inpatient care. Though most secondary community mental health models have a role in avoidance of long-term hospitalisation, the focus here is on models that are more obviously intensive forms of support that could substitute for an inpatient admission. These service models were similarly categorised into community-based, hospital-based and cross-setting approaches. Further information, and associated evidence, is provided about each model in Additional File 2 and Additional File 3.

#### 3.3.1. Community-based alternatives

##### Intensive residential services

This category encompasses ‘Residential rehabilitation services’, and ‘Intensive supported housing’ models (127). Individuals typically move through the components of this pathway with the ultimate aim of successfully managing an independent tenancy and avoiding re-hospitalisation.

‘Residential rehabilitation services’ offer communal residential living for adults with chronic mental health conditions and significant impairment in social functioning whose needs cannot be well-served in the community (128,129). Residential rehabilitation services are staffed 24-hours a day and provide people with a high level of support. People typically stay for several years (127), however some shorter-term residential facilities also exist. Some residential rehabilitation services aim to be specialised for specific diagnostic groups, such as individuals with a primary diagnosis of an eating disorder or psychosis.

‘Intensive supported housing’ represents a step-down from 24-hour residential rehabilitation care, offering time-limited tenancies in shared or individual self-contained units with high-level support, sometimes including 24-hours a day staffing (128,129). ‘Intensive supported housing’ can be offered with integrated care offered by the accommodation provider or with support outsourced and offered by external services (128,129). ‘Housing First’ and ‘Full Service Partnership’ represent two models of ‘supported housing with external intensive community support’ which typically serve communities experiencing or at-risk of homelessness and who also experience mental health difficulties (127,130–132).

##### Family placement schemes

Some family placement schemes can offer longer-term support to people with mental health difficulties who would otherwise not be able to manage living independently in the community. In the ‘Geel family foster care’ model, people of any age with various mental health difficulties are accommodated by foster families and are encouraged to participate in daily household activities and family life (133). The average length of stay with a Geel placement is approximately 30 years (133). ‘Healing Homes’, a similar model derived from Sweden, supports people with psychosis to live in host families for approximately 1-2 years (134). As well as offering support in an acute crisis (see *Section 3.2.1*.) the ‘Shared Lives’ model also operates as an alternative for individuals requiring long-term placements (33).

##### Enhanced case management models

These models aim to avoid the need for long-term inpatient admissions by providing intensive support to people with complex mental health needs within the community. ‘Specialist community mental health rehabilitation teams’ are a variation of traditional CMHTs which offer specialised rehabilitation care in the community, often working with people in intensive residential services or ensuring safe transitions between settings (128,129). Designated care coordinators oversee service users’ progression through the rehabilitation pathway and liaise with other providers in the statutory and voluntary sectors (128,129). Likewise, ACT, FACT and intensive case management models, as described in *Section 3.2.1*, can offer long-term enhanced case management support.

##### Discharge transition support

The ‘Peer Bridger Project’, described in *Section 3.2.1*, often works with people who have been hospitalised for long periods of time or who have experienced repeated hospitalisations, aiming to support successful transitions from inpatient care back into the community (53).

#### 3.3.2. Hospital-based alternatives

##### Inpatient psychiatric services

###### Inpatient psychiatric services with a specific therapeutic model

‘Hostel wards’ provide residential inpatient care for long-stay patients in inpatient mental health services who are unable to manage in the community and require 24/7 nursing care. Residents remain inpatients legally and staffing levels are similar to standard inpatient care settings. However, they are distinct from standard long-term wards in that the style of accommodation is more domestic, and residents are given a programme of domestic chores and self-care activities judged to be within their abilities (135).

As described in relation to acute settings in *Section 3.2.2*., the ‘Safewards’ programme has also been implemented in medium- and long-term inpatient settings (136).

###### Inpatient wards for people with specific psychiatric conditions

Specialist longer-term inpatient wards may offer tailored care for specific clinical groups, such as people with “personality disorder”, or other groups with specific needs, such as deaf people. An example identified in our consultation was the Springbank Unit, an inpatient unit for women with a diagnosis of “borderline personality disorder”, which also aimed to implement less restrictive risk management procedures (e.g., by offering service users an optional conversation with staff in the place of a formal risk assessment checklist when leaving the ward) (96).

#### 3.3.3. Cross-setting approaches

A range of cross-setting approaches were also identified in relation to long-term rehabilitation. Firstly, as described in *Section 3.2.3*, ‘Therapeutic communities’ and ‘DTCs’ may also offer an alternative approach to standard longer term inpatient care, in both inpatient rehabilitation, residential and day hospital settings (102,137).

Second, as described in *Section 3.2.3*, the ‘Trieste model’ also operates as a community alternative for longer term rehabilitation. For example, it provides people with supported housing, and there is a residential rehabilitation service which offers up to six months of support (138).

Third, as described above in *Section 3.2.3*, ‘Enabling Environments’ can be applied in a range of settings, including long-term rehabilitation settings for people with chronic mental health difficulties (105).

Finally, as described in *Section 3.2.3*, there are ‘Specialist consultancy’ models which aim to provide expert consultancy to health care organisations and intensive intervention to individuals to avoid and reduce long-term mental health hospitalisations (121,123–126).

### 3.4. Adult long-term inpatient care alternatives: forensic

A range of alternative service models were identified which operate in forensic settings across a variety of countries (see Additional File 5). Some of the service models identified below represent components of the Offender Personality Disorder (OPD) pathway in the UK, which operates exclusively in forensic contexts. This aims to offer a pathway of psychologically informed services for offenders with likely “severe personality disorder” (139). Further information, and identified evidence, about each model is provided in Additional File 2 and Additional File 3.

#### 3.4.1. Community-based alternatives

##### Residential community care

Similar to the long-term rehabilitation findings above (see *Section 3.3.1*), residential community care also serves offending populations in the form of either ‘intensive residential services’ or ‘secure accommodation’.

‘Intensive residential service’ options identified overlap with those for non-forensic populations. They include ‘Residential rehabilitation services’, ‘Integrated supported housing’ and ‘Housing with external intensive community support’ models (e.g., ‘Housing First’ and ‘Full Service Partnership’).

‘Housing and Accommodation Support Services’ (HASS) are specific to forensic populations. They are an ‘integrated supported housing’ component of the OPD pathway, specifically offering support and accommodation to people meeting OPD pathway criteria on probation who have been released from prison and other secure health care settings (e.g., inpatient care), or those who are transitioning on from other residential settings housing ex-offenders in the community (140,141).

‘Residences for the Execution of Security Measures’ (REMS), based in Italy, are a type of secure accommodation offering an alternative residential approach for forensic populations who have been given a custodial order. REMS are intended to be a therapeutic space with an aim for rehabilitation, without the involvement of police officers (142).

As described above in *Section 3.2.1* and *Section 3.3.1*, ‘Shared Lives’ is a family placement scheme which can offer support to offenders as well as non-offending populations (143).

#### Enhanced case management models

‘Forensic ACT’ teams are an adaptation of the general ACT model, involving co-ordination with criminal justice entities, provisions of legal advocacy and assistance with applying for financial support, and with a primary aim of reducing criminal justice involvement and reoffending (144). Forensic ACT teams operate in a similar manner to ACT teams, but with the added provision of forensic psychiatrists and probation officers usually (145).

##### Intensive Intervention and Risk Management Services (IIRMS)

‘IIRMS’ are a component of the OPD pathway which may be based in either prisons or the community. They offer psychologically-informed case management, working with individuals who are being released from a prison or secure environment into the community. IIRMS provide both in-reach to prison settings and outreach to community settings and aim to enhance skills and self-management necessary for a successful resettlement (146).

‘Transitional support and liaison services’ are a specialist type of IIRMS which aim to help people access support offered by statutory and voluntary organisations, both during and after their transition from custody to the community (147). The support duration is typically shorter, typically spanning just a few months, compared to standard IIRMS support, which typically extends for a year or longer (147).

#### 3.4.2. Hospital-based alternatives

##### Inpatient services

As described in relation to acute inpatient settings (*Section 3.2.2),* some forensic inpatient settings have employed therapeutic models such as the ‘Tidal Model’ (87), ‘Six Core Strategies’ (148,149) and ‘Safewards’ (83,136,150). Secure wards also exist which specifically accept offenders from certain groups, for example, deaf people or people with a diagnosis of “personality disorder”.

Another forensic inpatient model distinct to standard care are ‘Ter Beschikking Stelling’ (TBS) hospitals in the Netherlands. Dutch courts can impose combination verdicts on offenders with severe mental health difficulties and high risk of recidivism (151). These offenders can be detained in specialist TBS hospitals, which aim to reintegrate them back into the community (151). In contrast to forensic hospitals in the UK, TBS hospitals have wards with different levels of security within the same institution, offering greater continuity of care when people move between them, and are less restrictive (e.g., offering more people unsupervised leave and family visits) (152).

#### 3.4.3. Cross-setting approaches

Finally, there are also a range of approaches which can be applied across multiple settings. As described above in relation to acute contexts (*Section 3.2.3)* and long-term rehabilitation (*Section 3.3.3),* ‘Therapeutic Communities’ have been implemented in forensic settings, such as forensic inpatient and prison services, as have ‘DTCs’, which offer a whole-system approach to rehabilitation (103).

A further approach, ‘Psychologically Informed Planned Environments’ (PIPES) are another component of the OPD pathway. Whilst PIPES are primarily adopted in prison settings, they are also evidenced in alternative settings, such as community-based hostels for individuals who have recently been released from prison. PIPES offer training to staff to facilitate a safe and supportive environment according to several key principles: the development of an ‘enabling environment’; structured groups between staff and residents; socially creative sessions and training; and the provision of supervision and reflective spaces for staff (153). In a similar vein, as described above in relation to acute care (*Section 3.2.3*) and long-term rehabilitation *(Section 3.3.3*), ‘Enabling Environments’ can be found in a range of settings, including forensic services.

Some OPD services can operate in multiple settings. ‘OPD treatment services’ may be offered either in secure settings or in the community for individuals who are subject to probation supervision. For example, these may include Mentalisation Based Treatment services or Male-Trauma Recovery Empowerment Model services (154). ‘Psychologically enhanced resettlement services’, another component of the OPD pathway, also offer support either within prisons or within the community, and aim to support the transition-in, graduation, and transition-out of open conditions (155).

Finally, as listed in relation to the acute crisis care (*Section 3.2.3*), the ‘Sanctuary Model’ also operates in forensic settings (89), as do some ‘Specialist consultancies’ and the ‘Trieste model’.

### 3.5 CYP alternatives: Acute, long-term and forensic

A range of service models were also identified for CYP (see Additional File 6). Some were unique to this population, whilst others were also identified during the adult typology mapping and, as such, are described in greater detail in prior sections. Further information, and identified evidence, is provided about each model in Additional File 2 and Additional File 3.

#### 3.5.1 Community-based alternatives

##### Residential services

Similar to provision for adults – although not as widely implemented – crisis houses were identified for CYP aged 11 and over. Soteria houses and ‘Shared Lives’ are adult crisis residential models that can also accept CYP aged 16 and over (33).

A family placement scheme specific to CYP (aged three and over) is ‘Therapeutic foster care’, which involves structured therapy within a foster family setting, usually for 6-9 months (156–158). It can support forensic and non-forensic CYP populations and can be initiated at a time of crisis (158). It aims to provide CYP in foster care and youth justice programs with an alternative to more restrictive placements (158). The ‘Geel family foster care’ model is another family placement scheme, previously described for adults in *Section 3.3.1*, that can also provide long-term support to CYP of all ages.

Two further residential models of secure accommodation for CYP populations include ‘Secure children’s homes’ and ‘Secure training centres’ (STCs). Secure children’s homes are specifically tailored for CYP aged 10 to 17 years and offer full residential care, formal on-site education, and health care provision. On-site mental health professionals ultimately aim to successfully re-integrate service users back into the community (159,160). They accept CYP who are sentenced or on remand through the justice system, or placed due to local authority concerns that no other type of placement could keep them safe (159). Whereas STCs are detention facilities for CYP aged 12-17 years old who have been convicted of a criminal offence or are awaiting trial only. They are usually operated by private companies and their ethos is more punitive than in secure children’s homes, with lower staff ratios and less extensively trained staff (161).

As detailed in the adult section above, ‘Residential rehabilitation services’ and ‘Full Service Partnership’ - a ‘Housing with external intensive community support’ model - can also support CYP.

##### Acute day units

Both ‘General acute ADUs’ and ‘Specialist ADUs’ for eating disorders also exist for CYP, where support is tailored to the needs of CYP at the time of a crisis. ADU programmes for CYP may provide a combination of individual therapy, family therapy, medication management and educational services (157,162,163).

##### Home-based crisis services

‘CRHTTs’ and ‘Other intensive home treatment models’ can also provide support to CYP, sometimes with dedicated teams for CYP. A model specifically for CYP aged 11-18 years old is ‘Intensive home treatment with optional brief inpatient admission’. This involves providing a CRHTT approach with an optional short admission to a psychiatric intensive care unit, where the family is supported by the same professionals as in the community (164). Other CYP-specific models also include ‘Homebuilders’ and ‘Enhanced Homebuilders’, which offer short-term, intensive, home-based services aimed at resolving crises, improving family relationships and connecting families with necessary support (including financial support and respite care) (165). ‘Enhanced Homebuilders’ focuses on cultural competence and addressing violence (165).

##### Emergency service linked models

‘Police and/or ambulance street triage teams’ and ‘Clinician-only mobile crisis units’, previously described, can also provide support to CYP (43).

##### Drop-in crisis services

Similar to the ‘Crisis café’ model identified for adult populations (see *Section 3.2.1*), a few examples of crisis cafés for CYP were identified (157), alongside ‘Crisis assessment services’ and ‘Mental health crisis hubs’ (166,167).

##### Discharge transition support

‘Supported Discharge Service’ and ‘Hot-BITS’ models are discharge transition support models for CYP. Both models involve the provision of psychiatric reviews, psychological interventions, support with education, social issues and physical health, and out-of-hours support (168–171) to facilitate successful transition from inpatient care to the community.

##### Outpatient-based crisis services

The ‘Behavioural health crisis care clinic’ is a brief outpatient intervention model for CYP providing assessment, safety planning, coping skills and coordinating further care for those experiencing suicidal thoughts or recent suicide attempts (172). The ‘Whole-of-service stepped-care’ model, previously described for adults with a diagnosis of “borderline personality disorder”, can also support CYP aged 12 and over (54).

##### Community services with crisis function

###### Enhanced case management models

‘Crisis case management’ (CCM) is an enhanced case management model for CYP aged 10 and over. It provides shorter-term crisis support for families and additional resources like respite care (173). ACT has been used with CYP aged 10 and over, and FACT with CYP aged 12 and over (64,174,175).

###### Early intervention models

EIP services can accept CYP aged 14 years old and over, whilst FREED can accept CYP aged 16-25 (59,176).

###### Adolescent forensic community services

‘Adolescent forensic community services’ provide assessment and intervention for young people with complex mental health needs and a high risk of offending behaviour, and sometimes also provide in-reach to juvenile secure estates and children’s homes (157). They aim in part to avoid inpatient admissions and shorten inpatient lengths of stay (157).

#### 3.5.2. Hospital-based alternatives

##### Models involving general hospital medical care

###### Enhanced psychiatric liaison services

The ‘Enhanced psychiatric liaison models’ (‘Enhanced 24’ and ‘Comprehensive’ models) described above serve both adults and CYP aged 16 and over (177). ‘Paediatric psychiatric liaison’ services specifically provide support to CYP of all ages (178).

###### Time-limited admission to general medical wards with specialist community eating disorder team input

As described above in the adult section, this model also exists for CYP with eating disorders requiring medical stabilisation (179).

##### Brief-stay crisis units

Some types of ‘Brief-stay crisis units’ can also provide care to CYP, including ‘Psychiatric emergency service centres’, ‘23-hour crisis stabilisation units’ and ‘Behavioural assessment units’ (12,180).

##### Short-stay acute inpatient wards

‘Short-stay acute inpatient psychiatric wards’ also exist which can accept CYP aged 16 and over (27).

##### Inpatient psychiatric services with a specific therapeutic model

Some therapeutic models implemented in adult inpatient services have also been implemented in inpatient services for CYP, including ‘Safewards’ (181,182), ‘Six Core Strategies’ and the ‘HOPES’ model (90). Furthermore, an evaluation of the ‘BA’ model described in *Section 3.2.2*, adapted for adolescents, is currently in progress (183).

##### Inpatient wards for specific groups

As for adults, there are ‘Specialist eating disorder wards’ for CYP, and ‘Early psychosis wards’ which can accept CYP aged 16 and over. Specialist wards also exist for deaf CYP (184). There are also inpatient wards specifically for children under the age of 13, which may provide schooling, support to parents/carers and age-appropriate therapeutic activities (185).

#### 3.5.3 Cross-setting approaches

Multisystemic therapy is a cross-setting model specific to CYP aged 10 and over. It aims to engage multiple systems in order to provide intensive support to CYP and their families in their natural environments (157). It has been implemented with both forensic and non-forensic populations. ‘Wraparound with intensive services’, previously described, is for CYP aged 0-20 (116). Likewise, ‘Open Dialogue’ (but not peer-supported open dialogue), ‘Therapeutic Communities’, ‘DTCs’, the ‘Sanctuary model’, ‘Enabling Environments’, ‘Need-adapted Treatment’, the ‘Trieste model’, and the ‘Buurtzorg model’ have also been implemented for CYP populations.

## 4. Discussion

### 4.1. Key findings

The typologies we have presented show that a wide variety of approaches exist internationally which may act as an alternative to standard acute inpatient mental health care for adults and CYP, and to standard long-term and forensic inpatient care across a range of settings.

While we identified a range of community-based approaches, including some peer-led approaches, we also noted services which provide inpatient care in a substantially different way to the standard, including time-limited approaches, inpatient services with a particular therapeutic model (including trauma-informed approaches), and service models which aimed to reduce coercion and restrictive practice. These service models demonstrate that some improvements to care can be provided within existing structures. However, the fundamental needs of services, such as adequate resourcing and staffing, must be met if good quality crisis or rehabilitation care is to be provided, and we cannot assume that these innovations eliminate coercive or otherwise harmful elements of hospitalisation.

We also observed that exclusion criteria for the alternative models seemed to vary between individual services (see Additional File 3) and were often not stated in the literature. However, where this information was available, compulsory detention was a common exclusion criterion. Other exclusion criteria for some services included: people assessed as presenting with high risk to themselves or others, people with substance abuse difficulties and people with intellectual disabilities. This inequitable access to alternative models could have several disadvantages for these individuals, including limiting their choice of care, prolonging institutionalisation in more restrictive settings, reinforcing feelings of stigmatisation and disempowerment, and exacerbating distress. Furthermore, given that these are common comorbidities in mental health populations, this poses the question as to whether the identified models truly act as alternatives to inpatient care, if substantial numbers of those receiving inpatient care are excluded from accessing them. This is compounded by further observations that certain groups were not fully considered in existing service provisions. For example, there appeared to be relatively fewer models providing support to younger CYP, and CYP transitioning from children’s services to adult services. In line with previous literature (186,187), these observations reiterate that key gaps exist in service provisions for these populations. Mental health care services should not necessarily be considered as a one-size-fits-all approach – more granular consideration of sociodemographic and diagnostic factors in service delivery may help to improve the quality of care (188). Taken together, this necessitates the implementation of alternative models which can serve these groups, including services which can intervene before the point of detention for people at high risk.

We also identified a range of voluntary sector led models (such as ‘non-clinical crisis houses’, and ‘crisis cafés’) that make an important contribution to supporting individuals in crisis by providing an immediate response, contributing to prevention and recovery, and through the provision of a social rather than clinical approach (see Additional File 3). Evidence suggests that voluntary sector organisations are attractive and acceptable to people in crisis and can be more attractive to minoritized groups than statutory services (189). However, there are still some documented issues with geographical variability in service availability and inequalities in access for certain minoritized groups (189). However, voluntary and third sector organisations often lack resources to conduct rigorous research needed for larger-scale implementation of their services. As such, further quantitative and qualitative research is needed to evaluate different voluntary sector models, including their outcomes, their impact on promoting equality and their partnerships with public sector services (189).

### 4.2. Strengths and limitations

In this research, a comprehensive approach was taken to identify various inpatient alternatives. The search process involved reviewing literature and consulting experts to identify a wide range of alternative models. The inclusion criteria were broad, allowing for the inclusion of alternatives from any country, time period, and for people of any age. Models were also included irrespective of the level of evidence available for them, ensuring a comprehensive scoping of alternatives. Furthermore, key stakeholders were consulted throughout the project to ensure the real-world applicability of the resulting typology. Indeed, the development of international typologies of alternative service models offers a clear framework for understanding and categorising different types of mental health support. This can help to improve our understanding of international mental health care provision and, in turn, inform service planning, delivery and policymaking. These typologies can also help to identify gaps in local service provision, and to drive research and evaluation efforts for models which have received less funding and attention. These typologies may therefore be helpful for researchers, mental health professionals, policymakers and service users alike.

However, there are also limitations to consider in both the research process and the interpretability of our findings. It should be acknowledged our scoping exercise aimed to be broad and rapid, and its aim was to identify and describe existing models, not to evaluate their effectiveness. Though key information and literature associated with each model is provided, full systematic reviews which follow PRISMA guidelines (190) are required for a comprehensive description of each model and its associated evidence to better inform funding decisions and resource allocations.

Although experts from a range of countries responded to our call for information, the majority, including members of our expert working group, were based in England, so international models may not have been adequately captured. The typologies also do not capture models that fall outside of the scope of this study, including perinatal services, addiction services, services specifically for autistic people or people with intellectual disabilities, neurorehabilitation services, services for people living with dementia, and solely prison-based services. Furthermore, the typology development involved the categorisation of complex and diverse models into distinct groups. This can oversimplify how these models operate in practice and the heterogeneity within each model. In practice, the boundaries between different models may be more blurred. Similarly, whilst presenting the three separate typologies offered greater clarity for ease of interpretation, there was a large degree of overlap between both the CYP and adult models of care, and the acute and long-term models of care. Therefore, the distinctions between the different typologies can also be blurred. In future, alternative approaches to typology development could be taken – for example, with different scope or categorising models on the basis of different features (e.g., function versus form). Mental health service delivery is constantly changing, and so new models and approaches are likely to emerge over time; this typology provides a strong foundation which future research can build upon.

The focus of our typology on crisis provision constrains its ability to adopt a comprehensive system-wide approach that considers broader care pathways. It is important to consider the conceptualisation of mental health crises as ‘biographical disruptions’ – intense and extreme experiences which disrupt everyday life and potentially have far-reaching consequences – rather than episodes requiring an urgent response (189). This perspective highlights the importance of providing support not only during crises, but also before and after. It could be argued that effective mental health care at earlier stages, such as primary and preventative care, has the potential to prevent crises from occurring. Additionally, action to address systemic contributors to distress (e.g., social and economic inequalities, trauma, discrimination and marginalisation, limited community resources and social support) could help to promote wellbeing and prevent crises. Addressing such factors could involve policy changes, social reforms, advocacy and community empowerment. This highlights the importance of taking action to prevent crises and reduce the social determinants of mental ill health, not just focusing on crisis care provision.

### 4.3. Implications for research, policy and practice

It is apparent from this work, and existing literature, that crisis alternatives have proliferated in recent decades and crisis systems have typically become more complex (32). However, there is a limited evidence base to inform service planners’ decision-making. We have mapped out three international typologies of service models which may offer an alternative to standard acute and long-term inpatient mental health care (including in inpatient rehabilitation and forensic settings) for adults and CYP. Our typologies can help service planners and commissioners consider the whole range of options, when attempting to decide which crisis system improvements to prioritise, as well as guiding researchers in future attempts to explore the critical ingredients of crisis care systems.

Whilst we did not systematically assess the availability of the included models in this study, existing research suggests that the availability of many crisis services varies substantially geographically (32). A key constraint about the usefulness of models is the degree to which it has been feasible to implement them. Future research could further investigate the feasibility of implementing models in different contexts, including exploring barriers and facilitators to effective implementation. However, geographical variation in availability may also result from services being commissioned to meet specific local needs. Recent research reports few associations between any particular community crisis model and system-level outcomes – perhaps reflecting that the quality of care is most important (191). Some efforts have been made to define standards, key components and best practices of effective crisis services and pathways (192,193), which could help to improve the quality of crisis care. For example, research has shown that increasing model fidelity in CRHTTs leads to reductions in inpatient admissions (194). Future research could continue to develop and refine these best practice standards, investigate methods to enhance fidelity to them, and examine the impact of this upon outcomes and experiences of care.

Future research efforts should continue to evaluate these alternative models to better understand their effectiveness in practice, factors influencing their outcomes, and how they can be most effectively integrated into a crisis response system, and what works best for whom, when and how - prioritising models with an established and/or promising evidence base. Crucially, a focus on coproduced research is needed with people with lived experience of mental health difficulties, their families, clinicians, policymakers and commissioners to ensure specific service models in specific areas address the priorities of these key stakeholder groups.

#### Lived Experience Commentary, written by ROO, LM and KP

As compulsory hospital admissions rise in the UK, providing alternatives is vital. However, diverting someone’s care pathway around the most intense crisis point is not the only, or necessarily the best, way to reduce admissions – many admissions result from unmet needs elsewhere. It is vital to consider the whole ecosystem of care, including social safety nets, social work, and social care, which have been drastically eroded over the last decade and a half by austerity and a global pandemic. We are tired of continually suffering, and watching others suffer, avoidable crises triggered by punitive benefits reviews, workplace discrimination, housing problems, and/or lack of compassionate community services. For carers, watching loved ones be traumatised by repeated - sometimes punitive - admissions leads to hopelessness and relationship breakdowns. This is compounded when carers are not identified or included in care by those arranging admissions.

Disparities in detention rates suggest racialised groups, especially Black patients, cannot access alternatives to hospital equally or at the right time. Nor are racialised groups as likely as White patients to receive therapy (195–197) despite the World Health Organisation emphasising everyone’s rights to choice and meaningful support. Bearing this in mind, institutional racism will continually influence access to admission alternatives unless it is actively and consciously countered with culturally sensitive commissioning, valuing service user and carer voices. While it is tempting to seek scalable models to roll out nationwide, we also need local flexibility to work with local communities’ needs. This is a constant tension in policy work: the flipside of flexibility is a postcode lottery. It has been eye-opening to read about alternatives unavailable in our areas and disappointing to realise how much some groups are missing.

For the services that do currently exist, accessibility is an issue. Many people who need these services will not know they exist, and many services are ‘gatekept’, requiring a professional referral in order to receive support - but if you are not under a professional’s care, or the professional does not think you are suitable, then you won’t be able to access the service. Finally, a system at breaking point cannot deliver admission alternatives as intended. We have seen cracks in the UK system open wide enough to swallow us and our loved ones whole: people waiting weeks in a 24-hour assessment unit; living in “supported” housing offering little meaningful support; unable to work with crisis teams failing to provide basic consistency. We are left wondering how new admission alternatives can possibly be implemented in this climate; how such services can provide accountability to users and carers; and worried that overstretched providers might not resource them sufficiently to be safe.

## Data Availability

The Excel dataset generated via expert consultation and literature searching is available within this manuscript’s additional materials. Unpublished data and identifiable information from expert responses have not been made publicly available to protect participants’ confidentiality.

## Abbreviations

ACT: Assertive Community Treatment
BA: Brief Admission
CCM: Crisis Case Management
CMHT: Community Mental Health Team
CRHTT: Crisis Resolution and Home Treatment Team
CYP: Children and Young People
DTC: Democratic Therapeutic Communities
EIP: Early Intervention in Psychosis
FACT: Flexible Assertive Community Treatment
FREED: First episode and Rapid Early intervention for Eating Disorders HASS Housing and Accommodation Support Services
ICM: Intensive Case Management
IIRMS: Intensive Intervention and Risk Management Services
LEWG: Lived Experience Working Group
NHS: National Health Service
NICE: National Institute for Care Excellence
OPD: Offender Personality Disorder
PA: Preventative Admission
PIPEs: Psychologically Informed Planned Environments
REMS: Residences for the Execution of Security Measures
STC: Secure Training Centre
TBS: Ter Beschikking Stelling
UK: United Kingdom
USA: United States of America

## Acknowledgements

We are very appreciative for the many contributions of the academic, clinical and lived experience experts who provided valuable information, resources and time towards this project. Those who provided consent to acknowledgement are listed here.

We would like to thank Aisling Clifford, Amy Trunnell-Morris, Akiko Hart, Alessandra Martinelli, Ania Scigala-Ali, Arnhild Lauveng, Ben Hannigan, Bernadette McSherry, Bethan Thibaut, Bob Hinshelwood, Brian Martindale, Caroline Fox, Catriona Connell, Cherene Allen-Caraco, Christian Dalton-Locke, Claire Fraser, Crick Lund, Crystal Blyer, Daniel Eichert, Dennis Ougrin, Eamonn Flynn, Francis Silvestri, George Roycroft, Graham Thornicroft, Guiseppe Tibaldi, Harry Dyson, Helen Killaspy, Himali Pandya, Hollie Berrigan, Holly Salazar, Indira Vinjamuri, James Scurry, Jenny Molin, Joanna Monaghan, John Baker, John Dawson, Jorge Zimbron, Judith Moss, Keir Harding, Kenneth Minkoff, Kirk Turner, Klaus Lehtinen, Lars Kjellin, Lisa Neos, Lisette van der Meer, Lucy Fernandes, Lucy Maconick, Margaret Turner, Michael Kazich, Miram Berges, Natasha Tyler, Niel van Cleynenbreugel, Nigel Blackwood, Oliver Dale, Peter Cockersell, Pien Leendertse, Rachel Hastings-Caplan, Rafaella Pocobello, Rex Haigh, Robert Koch, Samuel Grenny, Sam Timimi, Shirley McNicholas, Shubulade Smith, Siobain Bonfield, Steve Miccio, Susie Walker, Tim Kendall, Vincenzo Passante, Jessica Griffiths, Wayne Lindstrom, Liz Durrant, Prathiba Chitsabesan and Kate Lorrimer.

## Author Contribution

All authors, except KP and MLC, substantially contributed to the conception or design of this study. Literature searching was conducted by JG, HB, RJ, JV, RP, KS. Expert consultations were conducted by JG and HB. Data extraction was conducted by JG, HB, RJ, JV, RP, RC, KS. JG and HB led on developing the typologies and drafting the manuscript with input and/or editing by all authors. All authors read and approved the final manuscript.

---

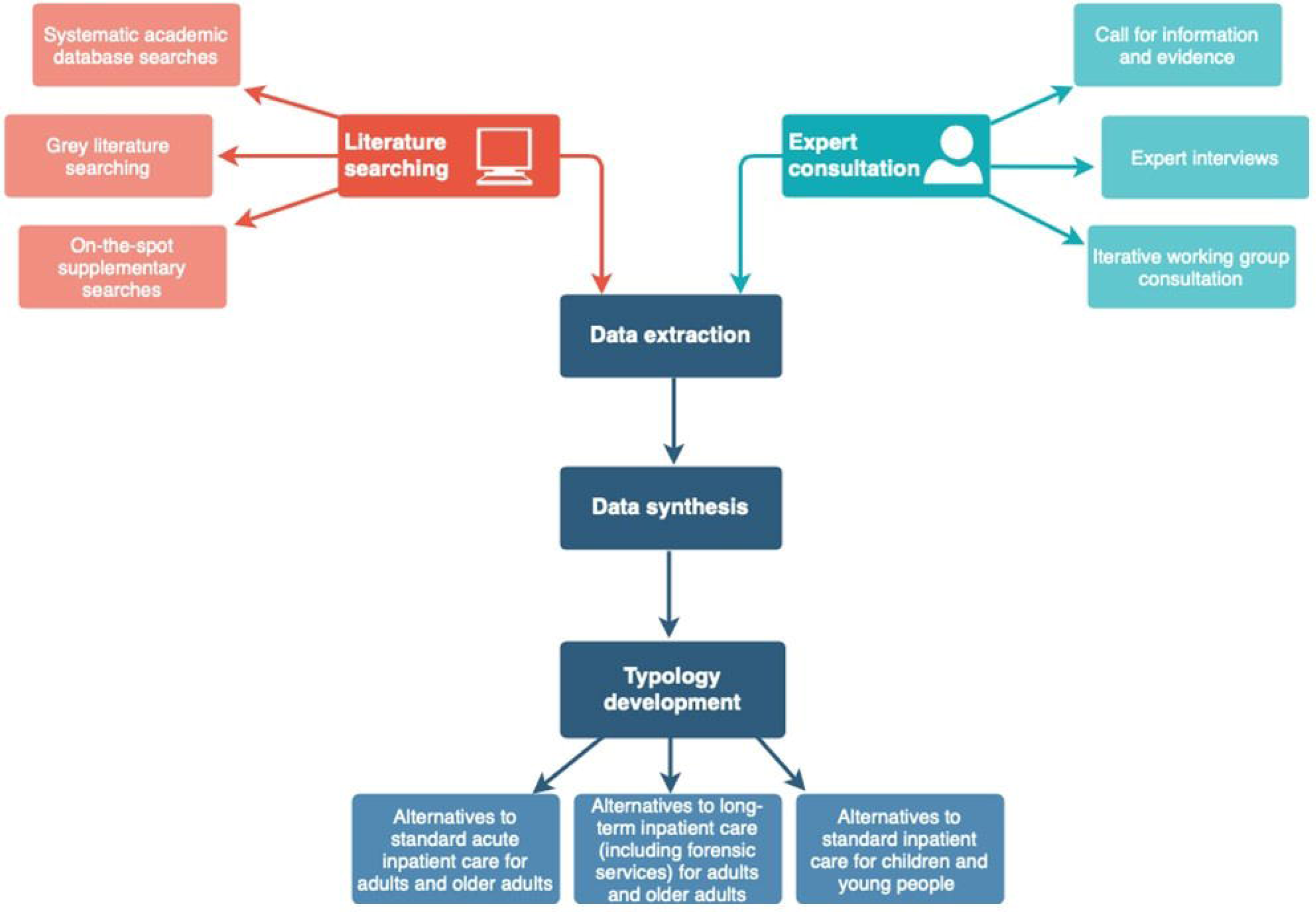

