## Additional File 1 for "Alternative approaches to standard inpatient mental health care: development of a typology of service models"

**Supplementary A: Database Search Strings**

| 1. | inpatient.ab. |
| --- | --- |
| 2. | admission.ab. |
| 3. | hospital*.ab. |
| 4. | 1 or 2 or 3 |
| 5. | alternative*.ab. |
| 6. | emerging*.ab. |
| 7. | differ*.ab. |
| 8. | 5 or 6 or 7 |
| 9. | "mental health".ab. |
| 10. | psychiatr*.ab. |
| 11. | psycholog*.ab. |
| 12. | psychosis*.ab. |
| 13. | schizophreni*.ab. |
| 14. | depressi*.ab. |
| 15. | anxi*.ab. |
| 16. | "substance use".ab. |
| 17. | addict*.ab. |
| 18. | suicid*.ab. |
| 19. | self-harm*.ab. |
| 20. | bipolar.ab. |
| 21. | bulimi*.ab. |
| 22. | anorexi*.ab. |
| 23. | "eating disorder*".ab. |
| 24. | autis*.ab. |
| 25. | ptsd.ab. |
| 26. | "post-traumatic stress disorder".ab. |
| 27. | adhd.ab. |
| 28. | "attention deficit hyperactivity disorder".ab. |
| 29. | "conduct disorder".ab. |
| 30. | "personality disorder*".ab. |
| 31. | forensic.ab. |
| 32. | "intellectual disabilit*".ab. |
| 33. | crisis.ab. |
| 34. | 9 or 10 or 11 or 12 or 13 or 14 or 15 or 16 or 17 or 18 or 19 or 20 or 21 or 22 or 23 or 24 or 25 or 26 or 27 or 28 or 29 or 30 or 31 or 32 or 33 |
| 35. | "systematic review".ab. |
| 36. | 4 and 8 and 34 and 35 |

**PsycInfo (via Ovid) String B:**

| 1. | "mental health".ti,ab. |
| --- | --- |
| 2. | "mental disorder*".ti,ab. |
| 3. | psychiatr*.ti,ab. |
| 4. | psycholog*.ti,ab. |
| 5. | psychosis.ti,ab. |
| 6. | schizophreni*.ti,ab. |
| 7. | depressi*.ti,ab. |
| 8. | anxi*.ti,ab. |
| 9. | "substance abuse".ti,ab. |
| 10. | addict*.ti,ab. |
| 11. | suicid*.ti,ab. |
| 12. | "self-harm".ti,ab. |
| 13. | bipolar*.ti,ab. |
| 14. | bulimi*.ti,ab. |
| 15. | anorexi*.ti,ab. |
| 16. | "eating disorder*".ti,ab. |
| 17. | autis*.ti,ab. |
| 18. | ptsd.ti,ab. |
| 19. | "post-traumatic stress disorder".ti,ab. |
| 20. | adhd.ti,ab. |
| 21. | "attention deficit hyperactivity disorder".ti,ab. |
| 22. | "conduct disorder".ti,ab. |
| 23. | "personality disorder*".ti,ab. |
| 24. | forensic.ti,ab. |
| 25. | "intellectual disabilit*".ti,ab. |
| 26. | crisis.ti,ab. |
| 27. | 1 or 2 or 3 or 4 or 5 or 6 or 7 or 8 or 9 or 10 or 11 or 12 or 13 or 14 or 15 or 16 or 17 or 18 or 19 or 20 or 21 or 22 or 23 or 24 or 25 or 26 |
| 28. | "acute day unit*".ti,ab. |
| 29. | adu.ti,ab. |
| 30. | "home treatment team*".ti,ab. |
| 31. | "home treatment crisis team*".ti,ab. |
| 32. | "crisis resolution team*".ti,ab. |
| 33. | "mobile crisis".ti,ab. |
| 34. | "crisis team*".ti,ab. |
| 35. | "community crisis".ti,ab. |
| 36. | "day hospital*".ti,ab. |
| 37. | "psychiatric decision unit*".ti,ab. |
| 38. | pdu.ti,ab. |
| 39. | safewards.ti,ab. |
| 40. | "crisis house*".ti,ab. |
| 41. | "crisis cafe*".ti,ab. |
| 42. | "peer-led service*".ti,ab. |
| 43. | "survivor-led service*".ti,ab. |
| 44. | starwards.ti,ab. |
| 45. | trieste.ti,ab. |
| 46. | soteria.ti,ab. |
| 47. | "medication-free".ti,ab. |
| 48. | "drug-free".ti,ab. |
| 49. | "mother and baby unit*".ti,ab. |
| 50. | mbu.ti,ab. |
| 51. | "peer respite".ti,ab. |
| 52. | "Short-stay hospital*".ti,ab. |
| 53. | "short-stay ward*".ti,ab. |
| 54. | "community-based crisis".ti,ab. |
| 55. | Haven*.ti,ab. |
| 56. | "family based".ti,ab. |
| 57. | "emergency accommodation".ti,ab. |
| 58. | "open dialogue".ti,ab. |
| 59. | 28 or 29 or 30 or 31 or 32 or 33 or 34 or 35 or 36 or 37 or 38 or 39 or 40 or 41 or 42 or 43 or 44 or 45 or 46 or 47 or 48 or 49 or 50 or 51 or 52 or 53 or 54 or 55 or 56 or 57 or 58 |
| 60. | "systematic review".ti,ab. |
| 61. | 27 and 59 and 60 |

**Pubmed String A:**

("inpatient" OR "admission" OR "hospital" OR "hospitalisation" OR "hospitalization" OR "hospitals" ) AND ("alternative" OR "alternatives" OR "emerging" OR "different" OR "differing") AND ("mental health" OR "mental disorder" OR "mental disorders" OR "psychiatry" OR "psychiatric" OR “psychological” OR "psychosis" OR "schizophrenia" OR "schizophrenic" OR "depression" OR "depressive" OR "anxiety" OR "substance use" OR "addict" OR "addiction" OR "addicted" OR "suicide" OR "suicidal" OR “self-harm” OR “self-harming” OR "bipolar" OR "bulimia" OR "bulimic" OR "anorexic" OR "anorexia" OR "eating disorder" OR "eating disorders" OR "autism" OR "autistic" OR "PTSD" OR "post-traumatic stress disorder" OR "ADHD" OR "attention deficit hyperactivity disorder" OR "conduct disorder" OR "personality disorder" OR "personality disorders" OR "forensic" OR "intellectual disability" OR "intellectual disabilities" OR "crisis") AND ("systematic review*")

**Pubmed String B:**

("mental health" OR "mental disorder" OR "mental disorders" OR "psychiatry" OR "psychiatric" OR "psychological" OR "psychosis" OR "schizophrenia" OR "schizophrenic" OR "depression" OR "depressive" OR "anxiety" OR "substance use" OR "addict" OR "addiction" OR "addicted" OR "suicide" OR "suicidal" OR "self-harm" OR "self-harming" OR "bipolar" OR "bulimia" OR "bulimic" OR "anorexic" OR "anorexia" OR "eating disorder" OR "eating disorders" OR "autism" OR "autistic" OR "PTSD" OR "post-traumatic stress disorder" OR "ADHD" OR "attention deficit hyperactivity disorder" OR "conduct disorder" OR "personality disorder" OR "personality disorders" OR "forensic" OR "intellectual disability" OR "intellectual disabilities" OR "crisis") AND ("acute day unit" OR "acute day units" OR "adu" OR "home treatment team" OR "home treatment crisis teams" OR "mobile crisis" OR "crisis team" OR "crisis teams" OR "community crisis" OR "day hospital" OR "day hospitals" OR "psychiatric decision unit" OR "psychiatric decision units" OR "safewards" OR "crisis house" OR "crisis houses" OR "crisis cafe" OR "crisis case" OR "peer-led service" OR "peer-led services" OR "survivor-led service" OR "survivor-led services" OR "starwards" OR "trieste" OR "soteria" OR "medication-free" OR "drug-free" OR "mother and baby unit" OR "mother and baby units" OR "mbu" OR "peer respite" OR "short stay hospital" OR "short stay hospitals" OR "short-stay ward" OR "short-stay wards" OR "community-based crisis" OR "haven" OR "havens" OR "family based" OR "emergency accommodation" OR "open dialogue") and ("systematic review*")

**Supplementary B: Grey literature databases and search strings**

**Databases:**

UK government official documents: <https://www.gov.uk/official-documents>

NHS UK: <https://www.nhs.uk/>

King’s Fund Library: <https://www.kingsfund.org.uk/consultancy-support/library-services>

CDC Stacks: <https://stacks.cdc.gov/welcome/>

RePORTER: <https://report.nih.gov/>

UCL Discovery: <https://discovery.ucl.ac.uk/>

NDLTD Global ETD Search: <http://search.ndltd.org/>

Medrxiv: <https://www.medrxiv.org/>

Washington State Health Care Authority: <https://www.hca.wa.gov/>

**Search strings:**

Mental health crisis care alternatives

Residential model for severe mental illness crisis

Crisis inpatient mental health

Crisis intervention response team

Inpatient crisis care alternatives

Inpatient mental health alternatives

Mental health inpatient crisis care alternatives

Forensic mental health crisis care

Forensic psychiatric inpatient care alternatives

Forensic mental health alternatives

Young people mental health crisis services

Young people mental health inpatient care alternatives

Young people residential crisis care

**Supplementary C: Call for information template**

Dear [expert],

We are contacting you on behalf of the **National Institute for Health Research Mental Health Policy Research Unit for England (MHPRU)** at University College London and Kings College London. The MHPRU is led by Professors Sonia Johnson and Alan Simpson, and is commissioned to provide evidence to inform NHS policy, as well as publishing academic papers and reports.

We are contacting you because we believe that you are an expert in this field and we hope you can help us identify models that provide alternatives to inpatient care across all age groups. These could include variations on inpatient care involving specific therapeutic models or specialist services for particular groups, or services in the community, including types of crisis houses, services providing crisis treatment at home or acute day units. We are also interested in alternatives to hospital emergency department attendance.

We are especially interested in innovative models that you consider may be substantially improving experiences or outcomes for young people, adults or older adults in mental health crises, or for people receiving longer term inpatient mental health care. We would be really grateful if you could reply to let us know of any model(s) that you think are relevant – web links, reports or contact details of people who can give further information would also be very helpful.

An email reply is fine but, if you prefer, we would also be happy to arrange a short video or phone call to discuss further any models you would like to tell us about. If you aren’t sure whether something is relevant or not, we’d be happy for you to tell us about it anyway so we can decide.

Further details of our project and criteria follow below and a participant information sheet is attached. We’d be delighted to hear from you!

Many thanks,

Jessica Griffiths, Helen Baldwin, Sonia Johnson, Alan Simpson & the MHPRU team

Mapping of evidence on alternatives to inpatient mental health care

**What is the aim of this project?**

This project is primarily being carried out to inform policy makers in England seeking to implement more effective and acceptable alternatives to inpatient care, but will also result in a published report. Our aims are to:

- Map out existing alternative models to standard inpatient mental health care across all age groups (including children and young people, adults and older adults), both nationally and internationally. We are also including longer term hospital care, including in rehabilitation and forensic settings.
- Examine any evidence relating to the effectiveness or outcomes of these alternative approaches (including qualitative literature).

To achieve these aims, we are gathering information via literature scoping and expert consultation. As part of the expert consultation phase, we are reaching out to experts in the field to draw on their knowledge of existing services.

**What information are we requesting from you?**

We would like to invite you to share:

1. **Details of any alternatives to standard inpatient mental health care** that you are aware of, in the UK or internationally, which provide care for children and young people, adults or older adults (which meet the criteria detailed below). This includes both different types of inpatient services and services outside of the hospital setting that aim to prevent admissions or result in early discharge. As well as general acute services, we are also interested in alternatives to secure and longer-term services.
2. **Contact details of any other academics, health professionals, policymakers, charitable representatives, professional networks and/or lived experience experts** who may also be able to assist with this call for evidence.
3. **Academic literature, brief reports, websites and/or other grey literature** which provide a description of services which offer an alternative to standard inpatient mental health care.

We have defined ‘alternatives to standard inpatient mental health care’ as services or initiatives which aim to serve people who would otherwise be admitted to an acute psychiatric ward or receive longer-term inpatient psychiatric care (e.g. in secure or rehabilitation services), and which meet **at least one** of the following criteria:

- Based outside of a hospital setting, including services that support people at home, in day care settings or in community residential settings with the aim of reducing pressure on hospital services
- *AND/OR* Dedicated to a specific diagnostic or sociodemographic group
- *AND/OR* Have a fixed maximum length of stay
- *AND/OR* Have implemented a specific therapeutic model involving changes in the practice of more than one profession within the service, or that involves different types of workers in care
- *AND/OR* A significant change in practice in the management of risk

We are aware of crisis and home treatment teams, crisis houses and acute day hospitals as types of alternatives to hospital: regarding these service models we are interested in implementations that you feel have potential to improve on standard care, by improving outcomes and/or experiences.

**How can you provide this information?**

You can provide this information by replying via email. We are also happy to arrange a brief video call if you would prefer to provide information verbally.

We are able to make a financial contribution towards any participating organisations in the charitable and voluntary sector choosing to participate in a phone or video call.

**How can you help?**

Please send us any information you know of on this topic.

If you have any questions about this request or would like to discuss anything further, please contact either Jessica Griffiths [email address removed] or Helen Baldwin [email address removed] and we can arrange a time to speak.

We thank you for your contribution.

Kind regards,

Jessica and Helen

**Supplementary D:** Professional background and location of experts who responded to the call for information

A total of 78 experts responded to our call for information and fed into our mapping.

**Supplementary Table 1.** The geographical location of experts who responded to the call for information.

| **Location of expert** | **Percentage of respondents (%)** |
| --- | --- |
| United Kingdom | 64% |
| United States | 14% |
| Italy | 5% |
| The Netherlands | 3% |
| Australia | 3% |
| Germany | 1% |
| Belgium | 1% |
| Finland | 1% |
| Norway | 1% |
| New Zealand | 1% |
| Sweden | 1% |
| Unknown | 4% |

**Supplementary Table 2.** The professional occupation of experts who responded to the call for information.

| **Professional background of expert** | **Percentage of respondents (%)** |
| --- | --- |
| Healthcare professionals | 33% |
| Academics | 18% |
| Clinical academics | 18% |
| Senior management | 10% |
| Policy makers | 6% |
| Charity workers | 5% |
| Experts by experience | 4% |
| Unknown | 5% |
