## Additional File 3 for "Alternative approaches to standard inpatient mental health care: development of a typology of service models"

**Additional File 3: Table providing a more detailed description of each alternative model and its associated evidence**

| Service model | Location & sector | Description of the model | Target population | Associated evidence |
| --- | --- | --- | --- | --- |
| Crisis houses (overarching category) | **Locations include:**  Adults: UK, USA, Australia, Switzerland, Israel.  CYP: UK, USA  **Sector:** Public or voluntary/ third sector  **Type of alternative:** Acute | Crisis houses (aka crisis hostels or crisis stabilisation centres) are community-based, residential services that tend to provide care to people in mental health crisis in small, homely settings (usually 8-12 bedded). A range of different types of support may be provided, including peer support, assessment, observation, activities, treatment-planning, medication management, safety-planning and referring on to other appropriate support. Crisis houses can serve as alternatives to inpatient admission, or as a step-down from inpatient care. Their maximum length of stay typically ranges between a few nights to a month (1). | **Age groups:** Adults & CYP (aged 11-18 years). Some are specifically for CYP.  People experiencing a mental health crisis. Exclusion criteria vary by service but can often include people who are involuntarily detained, people with no fixed abode, people who have an active high risk of self-harm or harm to others, active substance misuse, or violent behaviour. | **Adult:**  Systematic reviews (1,2);  Other evidence syntheses (3,4)  RCTs (5–11)  Non-randomised trial (12);  National surveys (13–15)  Personal communications (People USA 2023a; People USA 2023b)^a^;  Case study (16);  Grey literature (17).    **CYP:**  Other evidence synthesis (4);  Policy documents (18); Grey literature (19);  Personal communication (Hope House, 2023)^a^. |
| Clinical crisis houses | **Locations include:** UK, USA    **Sector:** Public  **Type of alternative:** Acute | Clinical crisis houses bear more resemblance to hospital services than non-clinical crisis houses. A high proportion of staff are nurses, and most have at least one staff member on site awake throughout the night. Care programme approach meetings are held in all clinical crisis houses (14). They tend to be integrated with crisis teams. ‘Crisis stabilisation centres’ (predominantly found in the USA) resemble clinical crisis houses. | **Age groups:** Adults and CYP (aged 11-18 years old). Includes services specifically for CYP.    People experiencing a mental health crisis. Exclusion criteria vary by service but can often include people who are involuntarily detained, people with no fixed abode, people who have an active high risk of self-harm or harm to others, active substance misuse, or violent behaviour. | National surveys (14). |
| Non-clinical crisis houses | **Locations include:**  UK, USA, Sweden, Finland, Germany, Switzerland, Hungary, Israel, Japan, France    **Sector:** Voluntary/ third sector  **Type of alternative:** Acute | Non-clinical crisis houses tend to be managed by the voluntary sector, with limited use of nurses or doctors, though can still well integrated with crisis teams. They are less likely than clinical crisis houses to have at least one staff member on site awake throughout the night (14). | **Age group:** Adults    People experiencing a mental health crisis. Exclusion criteria varies by service but can often include: people who are involuntarily detained, people with no fixed abode, people who have an active high risk of self-harm or harm to others, active substance misuse, or violent behaviour. | National surveys (14). |
| Specialist crisis houses | **Locations include:**  UK, USA, Sweden, Finland, Germany, Switzerland, Hungary, Israel, Japan, France    **Sector:** Public, private and voluntary/ third sector  **Type of alternative:** Acute | Specialist crisis houses cater to specific groups, for example specific sociodemographic groups (e.g., women or veterans) or specific clinical groups (e.g., people experiencing early psychosis or suicidal ideation).  Different approaches to care may be implemented in different crisis houses. For example, Soteria houses, a type of specialist crisis house for people experiencing early psychosis, aim to offer a non-coercive environment, with an emphasis on holistic support and person-centred approaches rather than medication. They resemble clinical crisis houses on most characteristics (14). Other crisis houses for people with early psychosis may adopt different approaches (e.g., a ‘early intervention in psychosis’ approach to care) (20). | **Age group:** Adults and CYP (aged 16 and over)    People experiencing a mental health crisis. The target population for specialist houses varies by service (e.g., women, veterans, people with specific difficulties e.g., psychosis).  Exclusion criteria vary by service but can often include people who are involuntarily detained, people with no fixed abode, people who have an active high risk of self-harm or harm to others, active substance misuse, or violent behaviour. | General  National surveys (14).    Soteria houses  Systematic reviews (1,21,22);  Other evidence syntheses (23–26);  Non-randomised trials (27,28); Pre-post studies (29–32); Policy documents (33);  Grey literature (34,35); Other (36,37) Personal communication with Soteria Bern, 2023^a^  Crisis houses for women Qualitative studies (38,39) RCTs (40)  Reports (33) Service evaluations (41) Books (42) |
| Peer-led crisis houses | **Locations include:**  UK, USA    **Sector:** Voluntary/ third sector  **Type of alternative:** Acute | Peer-led crisis houses (aka peer respites) are a type of crisis house which are staffed and run by trained peer support staff (43). They have been established both in the UK, and more widely in the USA. | **Age group:** Adults    Adults experiencing a mental health crisis. Exclusion criteria vary by service and over time. Pelot & Ostrow (2021) found that most do not have a formal policy restricting access to people experiencing suicidal ideation. In 2020, 38% (n = 12) peer respites restricted access to guests who had a plan for suicide (up from 19%, n = 16, in 2018). Most (66%) programs restricted people from staying if they did not have permanent housing (43). | Systematic reviews (2);  RCTs (7);  Pre-post studies (43–45);  Qualitative studies (46,47);  Policy documents (33);  Grey literature (48–50). |
| Crisis team beds | **Locations include:**  UK    **Sector:** Public  **Type of alternative:** Acute | Crisis teams can sometimes offer a small number of beds to people in crisis. They have a short length of stay and high level of integration with crisis resolution teams (14). They tend to support people with less severe mental health needs compared to other crisis services. Their staff teams rarely include nurses and doctors (14). | **Age group:** Adults    People experiencing mental health crisis but not requiring inpatient admission. People accessing crisis team beds tend to have less severe mental health needs than those accessing crisis houses or inpatient psychiatric care (14). | Pre-post studies (6,51);  National surveys (14). |
| Family sponsor homes | **Locations include:**  UK, USA, France. In 2019, only 5/174 (3%) crisis resolution teams in a national survey in England had access to a family sponsor home for adults (13).    **Sector:** Public  **Type of alternative:** Acute | Family sponsor homes (aka crisis family placement schemes) provide people experiencing a mental health crisis with residential support. Support includes involving the person in a normal family environment, and participation in meals and other domestic tasks and activities. Host families can support one or two acutely unwell people at a time. Host families are provided with training and support by local crisis services (1). | **Age group:** Adults    Adults experiencing a mental health crisis. Exclusion criteria include significant levels of risk of harm to self or others, or people who require changes to their treatment (52). | Systematic reviews (1);  RCTs (53);  Pre-post studies (52);  National Surveys (13). |
| Shared Lives | **Locations include:**  UK    **Sector:** Public  **Type of alternative:** Acute, long-term, forensic | Shared Lives is a social care model where the person with support needs moves in with or regularly visits a Shared Lives carer. They share the carer's home, family and community life. Support provided depends on the individual's needs, but can include: personal care (e.g., washing, dressing), practical help (e.g., with cooking, cleaning, shopping), helping them connect with others (e.g., family, friends, the community), support managing money, and accessing education, training, employment and volunteering (54). Different types of arrangement provided include long-term accommodation and support, short breaks, or day support (55). | **Age group:** Adults and CYP (aged 16-18 years old)    People aged 16+ who may have needs that may benefit from receiving day or residential support. People with a range of needs can be supported, including: people with mental health difficulties, people with intellectual disabilities, older adults, care leavers, people who have experienced domestic abuse or modern slavery (54). | Service evaluations (55);  Grey Literature (54,56);  Personal communication (Shared Lives Plus, 2023^a^. |
| Therapeutic foster care | **Locations include:**  UK, USA, Norway, Denmark, Sweden, The Netherlands and elsewhere in Europe.    **Sector:** Public, private  **Type of alternative:** Acute, long-term, forensic | Therapeutic foster care consists of structured therapy within a foster family setting. Children with severe emotional or behavioural are placed with trained foster carers (57), usually for 6-9 months (58). This approach is based on social learning theory and is delivered in a multi-agency context (58). Therapeutic foster care services are staffed by multidisciplinary teams including nurses, psychologists and psychiatrists. The aim is for children to be able to return to their homes after this period of therapeutic foster care (57). | **Age group:** CYP (aged 3-18 years old)    Children and young people with severe emotional or behaviour problems, and significant histories of mistreatment and trauma (59). Therapeutic foster care is also often targeted at young offenders (59). | Other evidence synthesis (57–59);  Grey literature (60)**;**  Other (61,62). |
| Acute day units (ADUs) (overarching category) | **Locations include:**  UK, USA, Europe  **Sector:** Public, private and voluntary/third sector  **Type of alternative:** Acute | ADUs (aka day hospitals, partial hospitalisation services) provide a crisis-focused, time-limited, non-residential service for people experiencing a mental health crisis. They aim to reduce admissions and facilitate earlier hospital discharge. They typically offer an on-site programme of interventions (including psychological treatments, practical and peer support, physical health support, support with daily living activities, relapse prevention, and medication-management) during the day, with service users returning home overnight and at weekends (63). Some ADUs can be specialised (e.g., ADUs for people with eating disorders or diagnoses of personality disorder, or older adults or CYP), whereas others provide more general support. | **Age group:** Adults & CYP (aged 5 years and over). Includes services specifically for CYP or older adults.  People in mental health crisis who are using or would be considered for acute psychiatric inpatient treatment, or other forms of acute care. Specific eligibility criteria may vary between services. Exclusion criteria varies by service, but may include a diagnosis of dementia, personality disorder, brain injury, learning disability or primary alcohol and substance misuse problems or current intoxication. | **Adult**  Systematic reviews (64–67);  RCTs (68);  National surveys (13,69);  Pre-post studies (70–73);  Mixed-methods cross-sectional studies (74);  Qualitative studies (63).    **CYP**  Other evidence syntheses (58,75);  Non-randomised trials (76);  Pre-post studies (77). |
| General acute day units | **Locations include:** UK, USA, Europe  **Sector:** Public, private and voluntary/third sector  **Type of alternative:** Acute | General acute day units provide general mental health support, accepting people experiencing a range of mental health difficulties, as opposed to providing specialist care for people with particular psychiatric diagnoses. | **Age group:** Adults & CYP (aged 5 years and over). Includes services specifically for CYP or older adults.  People in mental health crisis who are using or would be considered for acute psychiatric inpatient treatment, or other forms of acute care. Specific eligibility criteria may vary between services.  Exclusion criteria varies by service, but may include a diagnosis of dementia, personality disorder, brain injury, learning disability or primary alcohol and substance misuse problems or current intoxication. | National surveys (69). |
| Specialist acute day units for people with eating disorders | **Locations include:** UK, Australia, USA, Italy, Germany  **Sector:** Public and private  **Type of alternative:** Acute | Specialist acute day units exist for adults and CYP with eating disorders. These are similar to inpatient treatment in terms of having a similar multidisciplinary approach and offering similar treatments (with similar intensity and duration), but they do not allow overnight stays (78). They offer structured programmes that can include individual and group psychological interventions, group meetings and meal support. There may also be input from a dietician. Specialist acute day units for CYP with eating disorders also incorporate education into their programmes. They can serve as alternatives to inpatient admission, or as a step-down from inpatient care. | **Age group:** Adults & CYP (approximately 11 years and above). Includes services specifically for CYP.  People with eating disorders who are outpatients requiring more intensive support, or inpatient patients requiring a step-down service. People must be medically stable enough to attend day services.  Exclusion criteria vary by service, but services usually exclude people whose physical health needs cannot be met in the community or instances where the eating disorder is not the primary presenting diagnosis. | **Adult**  Systematic reviews (78–80);  RCTs (81);  Pre-post studies (82,83);  Case studies (84).  **CYP**  Systematic review (78,85);  RCT (86);  Grey literature (87). |
| Specialist acute day units for people with personality disorders | **Locations include:**  UK, USA, Canada, Denmark, The Netherlands  **Sector:** Public, private  **Type of alternative:** Acute | Most acute day unit programs for people with a diagnosis of personality disorder include weekly individual therapy and daily group therapy. They can vary in intensity and duration. Acute day units can be part of a step-down care model for people with a diagnosis of personality disorder. In these cases, people receive initial intensive treatment in a day hospital, and this is then followed by less intensive outpatient psychotherapy. | **Age group:** Adults  People with a diagnosis of personality disorder who require more intensive support than outpatient treatment, or a step-down in care from inpatient treatment. Exclusion criteria varies by service, but may often include: active risk to others, or acute substance addiction. | RCTs (88–91);  Non-randomised trials (69,70);  Pre-post studies (94–97). |
| Enhanced acute day unit treatment | **Locations include:** USA  **Sector:** Public  **Type of alternative:** Acute | Acute day unit treatment can be augmented/enhanced (e.g., with respite or outreach services, crisis beds, or extended hours programmes). | **Age group:** Adults  Adults requiring more intensive support than just attending a standard acute day unit programme. | Systematic review (65)**;**  RCT (98). |
| Crisis Resolution and Home Treatment Teams (CRHTTs) | **Locations include:** UK, USA, Norway, Spain, Netherlands, Denmark, Australia, South Korea, Switzerland  **Sector:** Public  **Type of alternative:** Acute | Crisis resolution and home treatment teams (CRHTTs) offer rapid assessment and short-term treatment to people experiencing a mental health crisis (99). They support people when primary care or other secondary mental health service support is insufficient (99). They aim to avoid emergency admissions by offering home-based treatment. They can also provide step-down care, aiding people's transitions from inpatient services back into the community. They also often act as a gatekeeper for inpatient admissions (99). In some places, assessment and home treatment functions are performed by the same team, whereas in others they are provided by separate teams. Some dedicated CRHTTs exist for CYP and older adults. | **Age group:** Adults & CYP. Includes teams specifically for CYP or older adults.  People experiencing acute mental health crisis, who, without intervention, are likely to require inpatient admission. Exclusion criteria varies by service but usually includes: substance or alcohol dependence as the primary presenting issue, intellectual disabilities, dementia or other organic mental health disorders. | **Adult:**  Systematic review (67,99–104);  Other evidence syntheses (3) RCTs (105–107); National surveys (13,108);  Pre-post studies (109–113);  Qualitative literature (114,115);  Grey literature (116).  Other (117)  **CYP:**  Systematic review (85);  RCTs (118,119).  **Older adults:**  Systematic reviews (120,121). |
| Homebuilders model | **Locations include:** USA, UK, Canada  **Sector:** Variable  **Type of alternative:** Acute | Homebuilders (aka the ‘family preservation model’) is a short-term, home-based intensive service for families who need additional support beyond typical outpatient services (57). The goal of the Homebuilders model is to keep children in their home environment. This is achieved by resolving the immediate crisis, teaching caregivers communication and other relevant skills, helping families improve relationships and linking children and families in with needed services (122). | **Age group:** CYP (aged 5-17 years approximately)  Families who need additional support beyond typical outpatient services. | Systematic review (123)**;**  Other evidence synthesis (57);  RCTs (122);  Grey literature (124)**.** |
| Enhanced Homebuilders | **Locations include:** USA  **Sector:** Unclear  **Type of alternative:** Acute | Enhanced Homebuilders (aka enhanced home-based crisis intervention) has the same programmatic goal as the original Homebuilders model (see row above) and offers the same services. In addition to the training that forms part of the original Homebuilders model, Enhanced Homebuilders service providers also receive training in cultural competence and working with families in which interpersonal and/or community violence may be an issue. | **Age group:** CYP (aged 5-17 years approximately)  Families who need additional support beyond typical outpatient services. | Systematic review (123)**;**  RCTs (122). |
| Other intensive home treatment services | **Locations include:**  Germany, Canada, USA  **Sector:** Public  **Type of alternative:** Acute | This category includes other clinician-only intensive home treatment models. These models involve the provision of intensive support to people at home to avoid the need for inpatient admissions. The staff teams do not include police or paramedic staff. Specific examples of models in this category include: ward-equivalent treatment (StäB) (125), the Mannheimer home treatment model (126,127) and a Canadian home treatment model (128,129). | **Age group:** Adults & CYP  People experiencing acute mental health crisis who are likely to require an inpatient admission, or people transitioning from inpatient care back into the community. | Systematic reviews (125,130); Pre-post studies (126–129). |
| Intensive home treatment team with optional brief admission | **Locations include:** The Netherlands  **Sector:** Public  **Type of alternative:** Acute | Intensive home treatment (131) is based on crisis resolution and home treatment team principles. It is provided for a maximum of four months to CYP aged 11-18 years old. Mental health professionals visit patients at their home and offer intervention to the family. The child/young person can have a short admission (maximum 2 weeks) to a psychiatric high and intensive care unit, together with their caregivers (131). During this admission they continue to be supported by the same professionals as in the community. | **Age group:** CYP (aged 11-18 years)  CYP aged 11-18 with severe psychiatric symptoms in need of acute and intensive treatment. Minimum average estimated intelligence (IQ > 70) (131).  Exclusion criteria includes a primary diagnosis of substance abuse disorders or disruptive behaviour. IQ of 70 or less (131). | Pre-post studies (131). |
| Police and/or ambulance street triage (overarching category) | **Locations include:**  UK, USA, Canada, Sweden, Australia  **Sector:** Public  **Type of alternative:** Acute | Police and/or ambulance street triage teams are teams where mental health staff work jointly with police and/or ambulance services to arrange appropriate assessment and support for people with mental health problems who come to the attention of the police and/or ambulance service and provide alternatives to using Section 136 of the Mental Health Act where possible (13). Some teams can include peer workers (132). | **Age group:** Adults & CYP. Includes some services specifically for CYP.  People with mental health problems who come to the attention of the police and/or ambulance service. Sometimes threat of violence to others can be an exclusion criterion (100). | Systematic reviews (100);  Narrative review (3);  National audit/survey (13);  Grey literature (132). |
| Police co-response teams | **Locations include:**  UK, USA, Canada, Australia  **Sector:** Public  **Type of alternative:** Acute | The police co-responder street triage model involves police officers pairing with mental health clinicians to provide appropriate assessment and support to people with mental health problems who come into contact with emergency services (100). Paramedics may also be part of the team (e.g., as in the Police Ambulance Crisis Response (PACER) model). Their aim is to reduce the likelihood of the person in crisis being detained in police custody, and to help reduce their distress (100). | **Age group:** Adults & CYP  People with mental health problems who come to the attention of the police and/or ambulance service. Sometimes threat of violence to others can be an exclusion criterion (100). | Systematic reviews (67,100);  Narrative review (133);  Pre-post studies (134,135);  National audit/survey (136);  Cross-sectional studies (137–140);  Service evaluation (141);  Policy document (142);  Qualitative studies (143) |
| Paramedic street triage teams | **Locations include:** Sweden, UK, USA, Australia, Denmark  **Sector:** Public  **Type of alternative:** Acute | These are mobile crisis unit which include paramedics, but not police (e.g., units comprising paramedics and mental health professionals such as nurses, psychiatrists, or social workers). Examples of paramedic street triage models include units staffed by paramedics and nursing staff (as in the Psychiatric Emergency Response (PAM) model in Sweden (144)) and Support Team Assisted Response (STAR) units staffed by paramedics and behavioural health clinicians (145). | **Age group:** Adults & CYP. Includes some services specifically for CYP.  People with mental health problems who come to the attention of the non-police emergency services (e.g., paramedics). | Systematic reviews (100);  Pre-post (144);  Descriptive studies (145,146);  Grey literature (147). |
| Clinician-only mobile crisis units | **Locations include:**  Adults: USA, Canada, Netherlands  CYP: USA  **Sector:** Public  **Type of alternative:** Acute | This category includes clinician-only mobile crisis units which respond to people experiencing a mental health crisis in the community. They receive calls, review cases, and go out to the person in crisis to help stabilise them, recommend appropriate services, and follow-up with them. These may be staffed by psychiatrists, nurses, psychologists, social workers, therapists or peer workers. They do not include police or ambulance staff. | **Age group:** Adults & CYP  People experiencing a mental health crisis in the community. | **Adult:**  Systematic reviews (100);  Non-randomised trials (148);  Pre-post studies (149,150);  Qualitative literature (151)  **CYP:**  Systematic reviews (100)  Cross-sectional studies (152)  Grey literature (153). |
| Crisis assessment services | **Locations include:**  UK  **Sector:** Public  **Type of alternative:** Acute | Crisis assessment services provide rapid assessment and referral on to other acute mental health services where appropriate. Around 2/3 of crisis assessment teams in England operate 24 hours a day, and 75% accept self-referrals from any member of the public as well as from professionals (13). | **Age group:** Adults & CYP. Includes teams specifically for CYP.  People experiencing acute mental health crisis requiring assessment. | National surveys (13,108). |
| Mental health crisis hubs | **Locations include:**  UK  **Sector:** Public and voluntary/ third sector  **Type of alternative:** Acute | Mental health crisis hubs were introduced during the first wave of the COVID-19 pandemic. They served as alternatives to emergency departments for people in mental health crisis (154). The aim was to divert people from emergency departments in order to reduce COVID-19 infection risk. They provide assessment, short-term management (e.g., whilst awaiting transfer into a clinically appropriate environment), and can signpost to appropriate support (154). | **Age group:** Adults & CYP. Includes some services specifically for CYP.  People experiencing a mental health crisis who do not require additional urgent medical attention (154). Exclusion criteria includes people detained under the Mental Health Act or people requiring urgent medical attention (154). | National audit/report (155);  Service evaluation (154);  Grey literature (156). |
| Crisis cafes (aka safe havens or recovery cafes) | **Locations include:**  Adults: UK, Australia, USA  CYP: UK  **Sector:** Public, voluntary/ third sector  **Type of alternative:** Acute | Crisis cafes provide an alternative to hospital emergency departments to people in crisis. They are usually provided by voluntary sector organisations. They provide out-of-hours assessment and immediate support to people experiencing a mental health crisis and can signpost to other services (13). Approximately a fifth in England are service-user led, and some employ peer workers (13). | **Age group:** Adults & CYP (aged 5 and above). Includes services specifically for CYP.  People experiencing a mental health crisis. Exclusion criteria varies by service but can include: people requiring urgent medical attention, people with active high risk of self-harm or harm to others, and people who are involuntarily detained. | National survey (13);  Cross-sectional studies (157);  Service evaluation (158);  Policy document (142);  Grey literature (159). |
| Peer-led crisis cafes | **Locations include:** UK  **Sector:** Voluntary/ third sector  **Type of alternative:** Acute | A survey showed that approximately a fifth of crisis cafes in England are service-user led (13). | **Age group:** Adults  People experiencing a mental health crisis. Exclusion criteria varies by service but can include: people requiring urgent medical attention, people with active high risk of self-harm or harm to others, and people who are involuntarily detained. | National survey (13). |
| Lifeguard pharmacies | **Locations include:**  UK  **Sector:** Public  **Type of alternative:** Acute | Lifeguard Pharmacy is a supportive signposting service. It is for people who are at risk of harm from themselves or others - including people who are experiencing suicidal thoughts, and people experiencing domestic abuse. People can visit one of the Lifeguard Pharmacies and request a free consultation with a trained member of staff discreetly. Staff then arrange a consultation for them with a trained member of staff (the Lifeguard) in a private consultation room as soon as possible. During this consultation, the Lifeguard provides assessment and signposting to appropriate services (160). | **Age group:** Adults  People who are in danger of harm from themselves or others, including people who are experiencing suicidal thoughts and people who are experiencing domestic abuse (160). | Grey literature (160)**.** |
| Transition to Recovery Program | **Locations include:** Australia  **Sector:** Voluntary/ third sector  **Type of alternative:** Acute | The 'Transition to Recovery' program is an intensive community outreach mental health program that supports peoples' transitions from inpatient mental health care back into the community. It provides people with social support, psychosocial education, and support with managing symptoms and day-to-day tasks. It also links people in with appropriate services and can provide assistance with attending appointments and community-based programmes (161). | **Age group:** Adults  People being discharged from inpatient mental health care or people who are at risk of being hospitalised. It can accept people who are homeless. People are usually excluded from these services if they are considered a danger to themselves or others, or if they require immediate hospitalisation. | Service evaluation (161);  Grey literature (162). |
| Peer Bridger Project | **Locations include:** USA  **Sector:** Public  **Type of alternative:** Acute, long-term | The Peer Bridger Project aims to support people's transitions from psychiatric hospitals into community life through providing intensive individual and group peer support services (163). ‘Peer bridgers’ (people who are successfully managing their own recovery from a psychiatric disability and who have received the required training from the project) are matched with the people accessing the project (usually 2-3 months prior to discharge from the hospital). Participation is voluntary. They collaboratively agree goals, roles and responsibilities. Peer bridgers provide mutual peer support both one-to-one to their 'matches' and by attending regular peer support groups. Support groups are based in hospital for people still admitted and based in the community for people to attend after being discharged from hospital (163). | **Age group:** Adults  Psychiatric inpatients who are willing to consider discharge and open to voluntarily participating in the Peer Bridger Project, especially those who have been hospitalised for a long period of time or have experienced repeated hospitalisations. Those who are unwilling to participate are excluded from these services. | Grey literature (163). |
| Supported Discharge Service (SDS) | **Locations include:**  UK, Germany  **Sector:** Public  **Type of alternative:** Acute, long-term | The SDS model aims to achieve transfer back to the usual community mental health teams. Care can include intensive case management, community (including home) treatment, day care in hospitals or any combination of these approaches. SDS teams operate from 8am-8pm with out-of-hour cover available at other times. They aim to establish contact with each young person admitted to inpatient services within 72 hours of admission. SDS closely follows the principles of assertive community treatment, including: small caseloads, a team approach with a practicing team leader, weekly formalised meetings and daily informal meetings, staff continuity, full responsibility for treatment services and for hospital discharge planning, no dropout policy, assertive engagement mechanisms and working with informal support systems (164). | **Age group:** CYP (adolescents)  Adolescents in inpatient mental health services.  For this specific service, adolescents had to be aged between 12-18 years old. Many were long-term patients with significant barriers to discharge. | Reviews (85);  RCTs (164,165);  Pilot evaluation (166);  Qualitative literature (167). |
| Hot-BITS model (aka BeZuHG-Modell) | **Locations include:** Germany  **Sector:** Unknown  **Type of alternative:** Acute | The Hot-BITS model combines inpatient and outpatient elements in a home treatment setting for CYP with complex mental health problems. It involves shortening people's inpatient stay and following-up with 12 weeks of home treatment (168). The Hot-BITS model includes the following components: thorough assessment; a focus on building the therapeutic alliance early before discharge from the unit; early discharge; establishing individualised treatment plans; and biweekly review of supervised by a child and adolescent psychiatrist; and crisis management available 10 hours a day, 5 days a week by a Hot-BITS team member, in addition to a 24/7 doctor on-call (168). | **Age group:** CYP (aged 5-17 years)  CYP with complex mental health difficulties. In the RCT by Boege et al. (2015) (168) evaluating the Hot-BITS model, to be eligible, participants needed to have been an inpatient for >72 hours, to have had a psychiatric diagnosis at admission, live in a family setting (e.g. with biological parents, relatives, foster family or a single parent), live within the catchment area and have an IQ of 70 or more. As such, those with an IQ below 70 are excluded from these services. | RCTs (168–171). |
| Whole of service stepped-care approach | **Locations include:** Australia  **Sector:** Public  **Type of alternative:** Acute | The 'whole of service stepped-care approach' is a stepped-care model of psychological therapy for people with a diagnosis of personality disorder in between usual crisis care and longer-term treatment in the community (172). The brief intervention clinic is community-based, and integrated with acute crisis services. Eligible people presenting to crisis services (e.g. inpatient units, emergency departments, crisis telephone triage) are identified and referred within 0-36 hours of acute presentation (172). The brief intervention provided by the clinic is manualised, and consists of a month of weekly sessions. People can then receive further outpatient psychological therapy afterwards if needed. | **Age group:** Adults & CYP (12 years and above)  People aged 12+ diagnosed with a personality disorder, who have had at least one inpatient admission within the last 18 months. | RCTs (172,173);  Other (174). |
| Behavioural health crisis care clinic | **Locations include:** USA  **Sector:** Private  **Type of alternative:** Acute | This outpatient service provides support to children and young people (and their families) who are experiencing suicidal thoughts or have a recent history of suicide attempts (but who are not an immediate risk to themselves). Up to four appointments are offered to families, usually one a week. In these sessions, the focus is on: conducting an assessment; working with the child and caregiver to create and follow a safety plan; helping the family to build skills in talking about the child's mental health needs; helping the child cope with difficult feelings and events and find safer ways to act; supporting the family to return to normal after the crisis passes; assessing which treatments are most likely to be helpful; linking the family into follow-up care and coordinating with the child's therapist if they have one (175). | **Age group:** CYP  CYP who are experiencing suicidal thoughts or have a recent history of suicide attempts, and their families.  Children are excluded from this service if they are in an emergency (i.e., at immediate risk of self-harm, or a risk to others), if the child is unable to participate in safety-planning to avoid self-harm or if they cannot take care of themselves. Cases are also excluded where the parent/guardian is concerned that they cannot keep their child safe until they have an appointment. | Grey literature (175). |
| Assertive Community Treatment (ACT) | **Locations include:**  UK, Norway, Australia, Switzerland, Germany, USA  **Sector:** Public  **Type of alternative:** Acute, long-term | Assertive community treatment (ACT) is a community-based alternative to hospital for people with severe mental health difficulties and complex needs. It takes a multidisciplinary team approach (usually at least two case managers, a nurse and a psychiatrist) and staff have small, shared caseloads (10:1 ratio of staff: cases). Staff are available 24/7, and support is delivered to people in their home environments. A range of services are provided (e.g., mental health support, housing, daily living skills, socialisation, employment, substance abuse treatment and crisis intervention). | **Age group:** Adults & CYP (aged 10 years and above). Includes services specifically for CYP and older adults.  People with severe mental health difficulties and complex needs, especially people who have experienced multiple inpatient admissions and have high levels of service use (176). | **Adult:**  Systematic review (104);  Other evidence syntheses (176);  RCTs (177–179);  Policy document (180);  Qualitative literature (181).  **CYP:**  Other evidence syntheses (58);  RCTs (182);  Policy document (183). |
| Flexible Assertive Community Treatment (FACT) | **Locations include:** Sweden, UK  **Sector:** Public  **Type of alternative:** Acute, long-term | Flexible assertive community treatment enables people to access intensive support delivered in the community using a team caseload and assertive community treatment principles, as and when they require it. In this model, care coordinators manage individual caseloads, but also work together to provide shared care for people at times of increased need, enabling seamless transition between high- and low-intensity care (184). | **Age group:** Adults & CYP (aged 12 years and over). Includes teams specifically for CYP.  People with severe mental health difficulties. | Other evidence syntheses (185);  Service evaluation (184);  Case studies (186);  Grey literature (187). |
| Intensive case management | **Locations include:**  UK, USA, Denmark, Sweden, Canada, Netherlands, Asia  **Sector:** Public and private  **Type of alternative:** Acute, long-term | Intensive case management is a community-based model designed to meet the needs of people with high levels of service use (188). It is a long-term intensive approach which provides service users with a comprehensive range of treatment, rehabilitation and support services (189). It has low staff to client ratios, provides outreach to clients in their natural environments, and practical assistance in a variety of areas (188). Staff caseloads are not shared. It aims to support people's recovery and psychosocial functioning in the community, to reduce hospitalisations and to prevent loss of contact with services (189). | **Age group:** Adults  People with long-term severe mental health difficulties and complex needs, especially people who have experienced multiple inpatient admissions and have high levels of service use. | Systematic reviews (189);  NICE review (190);  Other evidence syntheses (188,191);  RCT (192);  Policy document (180). |
| Peer-delivered case management | **Locations include:** USA  **Sector:** Public and voluntary/ third sector  **Type of alternative:** Acute | Peer-delivered case management (aka peer- or consumer-provided case management) involves peer workers (who have their own lived experience of mental health difficulties and/or substance use disorders) providing case management alongside other healthcare professionals (e.g., psychiatrists, nurses). | **Age group:** Adults  Adults with severe/chronic mental illness (193). They may have comorbid substance misuse difficulties. | Other evidence syntheses (193);  RCTs (194–196). |
| Crisis case management | **Locations include:** USA  **Sector:** Public  **Type of alternative:** Acute | Crisis Case Management (CCM) is a short-term intensive case management model which provides community-based support to children and young people with mental health difficulties in crisis (and their families). It is an adapted form of intensive case management. Compared to intensive case management, it is less labour intensive, more short term (supporting families for 4-6 weeks) and does not involve delivering clinical services in service users' homes (197). | **Age group:** CYP (aged 5 years and over)  CYP living at home who are at risk of inpatient admission as a result of psychiatric crisis (122). | Systematic review (123);  RCTs (122,197). |
| ‘Enhanced’ community mental health teams | **Locations include:**  UK, Australia  **Sector:** Public  **Type of alternative:** Acute | Enhanced community mental health teams, which may also be known as ‘extended hours’ teams, are extended community mental health teams which offer standard ongoing case management at home or in the community, as well as integrated crisis response and intervention. The professionals in these teams are drawn from local community mental health teams on a rota basis to provide crisis responses in and out of office hours (198). | **Age group:** Adults  People with any psychiatric disturbance requiring secondary mental health care input. No exclusion criteria based on type of psychiatric disturbance (198). | Service description (198) |
| Early Intervention in Psychosis (EIP) services | **Locations include:** Widespread (e.g. UK, USA, Australia, Canada, Netherlands, Scandinavia, Spain, China)  **Sector:** Public  **Type of alternative:** Acute | Early Intervention in Psychosis (EIP) services are community-based multidisciplinary teams that support people for up to three years after their first episode of psychosis. They aim to reduce treatment delays at the onset of psychosis, and promote recovery by reducing the probability of relapse following a first episode of psychosis. They can provide a range of support including: assessment; a range of pharmacological and psychological interventions; support, information and advice for families and carers; support with employment, training and/or education; physical health monitoring; support with other comorbid mental health difficulties (e.g., low mood, anxiety, substance misuse); crisis planning; relapse prevention work; and support with social care issues such as housing or debt management (199). | **Age group:** Adults & CYP (aged 14-65 years)  People who are experiencing a first episode of non-organic psychosis, or suspected psychosis. People at-risk of developing psychosis can also be supported. People may be excluded in cases where psychosis is not the primary disorder (e.g., primary personality disorder with psychotic symptoms) (200). | Systematic review (201);  RCTs (202,203);  Pre-post studies (204);  Policy document (205);  Other (200);  Grey literature (199,206). |
| Early Intervention in Eating Disorder (FREED model) | **Locations include:**  UK, Australia  **Sector:** Public  **Type of alternative:** Acute | FREED is an evidence-based package of care for 16–25-year-olds experiencing a first episode of an eating disorder (of less than 3 years in duration). FREED aims to reduce the duration of an untreated disorder and intervene before the behaviours become more embedded. It operates within existing community eating disorder services by providing eligible service users with fast-tracked access to specialised treatment which is adapted to the specific needs of this age group. Each service has a ‘FREED champion’ which coordinates the care of these individuals. | **Age group:** Adults & CYP (aged 16 years and over)  Individuals who have had an eating disorder for three years or less. As such, people who have had an eating disorder for three or more years are excluded from these services. | Qualitative literature (207);  Pre-post studies (208–212)  Grey literature (213–215). |
| Enhanced psychiatric liaison services (overarching category) | **Locations include:** Widespread (e.g. UK, USA, Europe, Australia, China, Japan)  **Sector:** Public  **Type of alternative:** Acute | Liaison psychiatry services provide assessment and treatment to people with comorbid physical and mental health needs. They may provide input to acute general hospitals, other specialist hospitals and primary care. In the UK, they tend to be hospital based teams which provide on demand consultation and treatment for patients in acute hospital settings (including in emergency departments and medical wards), but some also provide some outpatient work or specialist in-reach to particular medical teams. Enhanced psychiatric liaison models (e.g., 'Enhanced 24' and 'Comprehensive' models) offer more specialised care, increased psychiatric consultant input, and enhanced follow-up support , in part aiming to help prevent avoidable inpatient admissions (216). | **Age group:** Adults & CYP. Includes specific services for CYP and older adults.  Patients in acute hospitals (including emergency departments) with mental health problems (e.g., comorbid physical and mental health problems, self-harm, dementia, drug and alcohol related problems, behavioural disturbance). | Systematic review (217);  NICE review (218);  Narrative review (3);  National survey (219);  Qualitative literature (220);  Service evaluation (216);  Policy documents (221); Grey literature (222). |
| Enhanced 24 (including the RAID model) | **Locations include:**  UK  **Sector:** Public  **Type of alternative:** Acute | The ‘enhanced 24’ model performs the same functions as the ‘core 24’ model, but provides more specialist care (e.g., in addictions and drug and alcohol use, older adults, and mental health problems in people with learning disabilities). ‘Enhanced 24’ model services have more consultant psychiatrist input and are able to provide more follow-up care. Support may extend to medical outpatients. An example of the 'enhanced 24' model is the Rapid, Assessment, Interface and Discharge (RAID) model. | **Age group:** Adults & CYP (aged 16 years and above)  Patients in acute hospitals with comorbid mental health and physical health needs, including those for which they have specialist expertise (e.g., younger people, frail older adults, people with substance misuse difficulties, people with intellectual disabilities) (223). | Systematic review/ health technology assessment (67);  Policy document (221,223);  Service evaluation (216); Pre-post (224). |
| Comprehensive psychiatric liaison services | **Locations include:**  UK  **Sector:** Public  **Type of alternative:** Acute | Comprehensive model psychiatric liaison services deliver core 24 services and provide enhanced expertise and input to planned care pathways across all inpatient and outpatient settings. This includes assessment and treatment for conditions such as chronic pain and medically unexplained symptoms. This model is usually used in large secondary care centres with regional and supra-regional services. | **Age group:** Adults & CYP (aged 16 years and above)  Patients accessing large secondary medical care centres who have comorbid physical health and mental health needs. Specific eligibility criteria can vary by service. | Policy documents (221,223). |
| Paediatric psychiatric liaison services | **Locations include:**  UK  **Sector:** Public  **Type of alternative:** Acute | Paediatric psychiatric liaison services are psychiatric liaison services that specifically serve children and young people under the age of 18. Liaison psychiatry support for CYP at general hospitals is provided by child and adolescent mental health services (CAMHS). It is usually provided on a case-by-case outreach basis - only a minority of areas have a dedicated liaison psychiatry service. | **Age group:** CYP (under 18)  Children and young people at general hospitals who have co-occurring physical and mental health needs | Policy document (225). |
| Brief-stay crisis units (overarching category) | **Locations include:** UK, USA, Canada, Australia, Belgium, The Netherlands, France, Singapore  **Sector:** Public, private  **Type of alternative:** Acute | Brief-stay crisis units include a variety of different service models, such as emergency psychiatry assessment, treatment and healing units (EmPATH units), behavioural assessment units, psychiatric observation units, psychiatric decision units, 23-our crisis stabilisation units and psychiatric emergency service centres. They are hospital-based units which provide support for short, time-limited periods (typically between 24-72 hours in duration). They provide environments for stabilisation, assessment and onward referrals. They aim to reduce emergency department mental health presentations and wait times, and/or psychiatric admission. | **Age group:** Adults & CYP. Includes services specifically for CYP.  People experiencing mental health crisis but not requiring a full inpatient admission. Exclusion criteria of brief-stay units identified in a systematic review included: people under the influence of or dependent on drugs or alcohol, aggressive behaviour, medical issues, residing out of the catchment area, pattern of self-harming or requiring an inpatient admission (226). | Systematic review (226)**.** |
| Psychiatric observation units | **Locations include:** USA  **Sector:** Public, private  **Type of alternative:** Acute | Psychiatric observation units are hospital-based units that allow for overnight stays for a limited time period, that provide stabilisation, assessment and further referral, with the goal of reducing emergency room wait times and psychiatric hospitalisations. | **Age group:** Adults  People in crisis who likely require an observation stay of 48 hours or less or who require inpatient admission but no bed availability. | Systematic review (226)**;**  Service evaluations (227). |
| Psychiatric decision units | **Locations include:**  UK, France, Singapore, USA, Australia  **Sector:** Public  **Type of alternative:** Acute | Psychiatric decision units are short-stay facilities, typically based within psychiatric hospitals, for referred voluntary patients in acute mental health crisis. They are open 24/7, accept only voluntary patients, usually provide recliner chairs in partitioned rooms for sleeping rather than beds, and limit stays to 24-72 hours (228). They offer enhanced assessment and short-term support, which is typically cognitive or psychosocial. They are often nurse-led. | **Age group:** Adults  Targeted at people who experience excessive stays in emergency departments, frequently use other services (e.g., police or ambulance services) and who have frequent crisis-related needs. All studies in Goldsmith et al.’s (2021) systematic review would only accept voluntary admissions (228). | National surveys (13,228);  Service evaluations (229). |
| 23-hour crisis stabilisation units | **Locations include:** USA  **Sector:** Private  **Type of alternative:** Acute | 23-hour crisis stabilisation units provide up to 23 consecutive hours of crisis respite and observation in the community. They provide assessment, stabilisation and determination of the level of care needed. They aim to provide an alternative to emergency department presentations, and to avoid unnecessary hospitalisations for people whose crisis may resolve with time and observation (4,18). Some may employ peer workers (People USA, personal communication). | **Age group:** Adults & CYP (aged 5 years and above). Includes services specifically for CYP.  People in crisis experiencing mental health and/or substance use problems. The following exclusion criteria may be applied: lack of voluntary consent to admission/treatment; if the person can be treated in a less restrictive settings; if they can only be treated in an inpatient setting; if their medical needs cannot be met in this setting; people requiring one-to-one support; and people with co-occurring substance use disorders where the substance use is the primary presenting problem. | **Adult:**  Other evidence synthesis (230);  Personal communication (People USA, 2023)^a^; Grey literature (231–234).  **CYP:**  Other evidence synthesis (4);  Policy documents (18,235). |
| Psychiatric emergency service centres | **Locations include:** USA, Belgium  **Sector:** Public, private  **Type of alternative:** Acute | Psychiatric emergency service centres are stand-alone emergency departments specifically for psychiatric patients. They provide immediate psychiatric assessment and provide a therapeutic environment for people in crisis to receive psychiatric, medical and social support (67,236). Patients are triaged by a separate staff of mental health professionals and medically assessed in conjunction with the emergency department. Patients can typically stay for up to 24 hours. They are considered outpatient services. They must support walk-ins. | **Age group:** Adults & CYP. Includes services specifically for CYP.  People who pose a high level of risk of harm to themselves or others and who may have a high level of functional impairment with behavioural symptoms that cannot be stabilized in a less restrictive setting. | Health technology assessment (67);  Cross-sectional studies (237,238);  Pre-post implementation studies (239,240);  Naturalistic comparison studies/ quasi experimental (241,242);  Retrospective chart review (236);  Grey literature (234,243). |
| Behavioural assessment units | **Locations include:**  Australia  **Sector:** Public  **Type of alternative:** Acute | The ‘Behavioural assessment unit’ model of care aims to create a safe and therapeutic environment for people experiencing behavioural disturbances, including people with acute and chronic substance abuse difficulties, people in psychosocial crisis, and people with acute psychiatric conditions (244). Behavioural assessment units are specifically designed to allow close observation and provide timely access to specialist expertise and facilities required for sedation and restraint when required. | **Age group:** Adults & CYP (aged 16 years and above)  Patients with acute behavioural disturbance, specifically behaviour influenced by drugs and alcohol, drug intoxication, mental health difficulties and social crises. A diagnosis of a mental health condition is not a prerequisite for admission. People at risk of requiring intubation are excluded from these services. | Pre-post design (244). |
| Emergency psychiatric assessment, treatment and healing (EmPATH) units | **Locations include:** USA  **Sector:** Private  **Type of alternative:** Acute | An outpatient hospital-based program that accepts emergency department patients in a psychiatric crisis. Once the patient arrives to the EmPATH unit, they are placed on suicide precautions to ensure safety. Each patient receives a nursing evaluation, a psychosocial evaluation from social workers, and a psychiatric evaluation from a psychiatrist. The interdisciplinary treatment team works to identify the underlying aetiology and contributing factors to the individual's suicidal ideation and treatment is initiated. Treatment modalities include individual and family therapy, medication management, referrals for ongoing treatment of substance use disorders, and assistance with financial and housing insecurity. | **Age group:** Adults  Adults in acute mental distress. People are excluded from these services if they are under 18 years old, medically unstable, need co-management of a medical condition, incarcerated, actively violent, or intoxicated (245). | Systematic review (226)**;**  Economic evaluation (246);  Pre-post implementation study (245). |
| Time-limited admission to a general medical ward with specialist community eating disorder team input | **Locations include:**  UK  **Sector:** Public  **Type of alternative:** Acute | Sometimes people who are unwell as a result of an eating disorder are admitted to general medical wards for medical stabilisation, instead of being admitted to general psychiatric inpatient units or specialist eating disorder units (247). Admissions are time-limited (up to three weeks generally, although this can vary in practice). Specialist community eating disorder services work closely with the medical wards to ensure the safe treatment of people with eating disorders close to home, and without needing to wait for a bed in a general inpatient psychiatric ward or specialist eating disorder unit. | **Age group:** Adults & CYP. Includes services specifically for CYP.  People with eating disorders who have medical needs that require inpatient medical care (e.g., very low weight or high medical risk) (247). There may be some patients who are so physically ill that their needs cannot be met within a specialist eating disorder inpatient unit. In these cases, they may also be admitted to general medical wards. | NICE guidelines (248); RCT (86);  Other (247);  Grey literature (249). |
| Safewards | **Locations include:**  UK, Australia, Denmark, Germany, Canada, Finland, Poland  **Sector:** Public, private  **Type of alternative:** Acute, long-term, forensic | Safewards is an organisational approach to delivering inpatient mental health services, which aims to reduce levels of conflict and restrictive practices in mental health inpatient settings. It consists of ten core interventions that were identified through a rigorous testing and refining processes. | **Age group:** Adults & CYP. Including in CYP-specific settings.  People on inpatient psychiatric wards (acute, long-term or forensic). | **Adult:**  Systematic reviews (250,251);  Other evidence reviews (252–254);  RCTs (255);  Pre-post studies (256,257);  Descriptive paper (258);  Policy documents with an integrated pre-post research component (259);  Mixed-methods paper (260).  **CYP:**  Pre-post implementation study (261);  Policy documents with an integrated pre-post research component (259). |
| Star Wards | **Locations include:**  UK  **Sector:** Public, private  **Type of alternative:** Acute | Star Wards is a quality improvement initiative which was set up by Marion Janner in 2004 and run by the charity Bright until 2020. The aim was to work with a full range of mental health wards to promote the implementation of 75 practical, low-cost ideas to improve mental health inpatient care - 80% of UK mental health wards are members, and membership is free for NHS wards (262). | **Age group:** Adults  People on acute inpatient mental health wards. | Survey report (262). |
| Talk 1st | **Locations include:** UK  **Sector:** Public  **Type of alternative:** Acute | Talk 1st is an amalgamation of the Safewards and Star Wards models (see separate Safewards and Star Wards models for further detail). It aims to reduce the need for restrictive interventions in inpatient psychiatric wards. | **Age group:** Adults  People on inpatient mental health wards. | Grey literature (263). |
| Tidal model | **Locations include:**  UK, Ireland, New Zealand, Canada, Japan, Australia  **Sector:** Public  **Type of alternative:** Acute, forensic | The Tidal Model attempts to avoid a reductionist approach where people are viewed as patients with symptoms to be treated. Instead, it focuses on people’s own narratives of illness and their own perception of their problems (1). It aims to respect individuals’ knowledge and expertise about their own lives. In the model, staff are encouraged to take a curious approach as a way of validating people’s experiences. They also aim to find common language to describe a person’s situation (264). Frequent, collaborative contact between staff and patients is encouraged through regular assessment of problems and goals. This involves documenting individuals' expressed needs and problems verbatim (1). | **Age group:** Adults  People on acute and forensic inpatient mental health wards. | Systematic review (1);  Other evidence synthesis (265);  Pre-post implementation studies/service evaluations (266,267);  Qualitative literature (268);  Service description (264). |
| Bradford refocusing model | **Locations include:**  UK, Germany  **Sector:** Public  **Type of alternative:** Acute | The Bradford Refocusing Model was developed on acute wards in Bradford, UK in the late 1990s. The model increased nurses' authority to take risk management decisions, thus minimising or eradicating formal observations on wards, instead allowing more time to spend on more 'collaboratively agreed contact between staff and patients' (1). | **Age group:** Adults  People on acute inpatient mental health wards. | Systematic review (1);  Other evidence synthesis / model description (269);  Case studies (270). |
| Six Core Strategies | **Locations include:** UK, Australia, USA, Finland  **Sector:** NA  **Type of alternative:** Acute, forensic | Six Core Strategies aims to reduce seclusion and mechanical restraint. It involves: senior management commitment to change, using audit to inform practice, workforce training, use of assessment tools, patient involvement, and debriefing techniques. | **Age group:** Adults & CYP. Including in services specifically for CYP.  People on an acute inpatient mental health ward, or in forensic settings. | Scoping review (271);  Cluster RCT (272);  Pre-post implementation studies (273–275);  Qualitative literature (276);  Policy documents (277). |
| The Sanctuary model | **Locations include:** USA  **Sector:** NA  **Type of alternative:** Acute, long-term, forensic | The Sanctuary model is a 'blueprint for clinical and organisational change which, at its core, promotes safety and recovery from adversity through the active creation of a trauma-informed community' (278). It was initially developed for adult trauma survivors in short-term inpatient treatment settings. However, it has since been adapted for a variety of settings including adolescent residential treatment programmes and forensic settings (271). | **Age group:** Adults & CYP. Including services specifically for CYP.  Anyone accessing an organisation implementing the Sanctuary Model (which can include inpatient, residential, day hospital, and community settings). | Scoping review (271).  Grey literature (278). |
| Drug-free or minimal medication wards | **Locations include:** Norway  **Sector:** Public, private  **Type of alternative:** Acute | In Norway, medication-free inpatient services have been introduced into national policy for reducing coercion, after being suggested by a service user organisation arguing for patients' rights not to be coercively treated with antipsychotic medication. The Norwegian government made medication free services mandatory. | **Age group:** Adults  Mostly people who have previously tried mainstream psychiatry services. | Qualitative literature (279);  Qualitative analysis of policy document (280);  Grey literature (281,282)**.** |
| Brief Admission model | **Locations include:** Sweden, the Netherlands, Norway  **Sector:** Public  **Type of alternative:** Acute | Brief admission (BA) by self-referral is an intervention which allows people to hospitalize themselves in a pre-negotiated way (283). It may be used with service users with emotional instability and self-harm, including people with a diagnosis of borderline personality disorder. When the service user is not in crisis, they draw up a contract with clinicians. This contract specifies the agreed frequency and duration of admissions (with a maximum stay of three days). It also agrees the service user's goals, preferred approaches from staff, and stress-reducing activities. The contract also states the service user's responsibilities (e.g., to not harm themselves or others, or to not be under the influence of drugs or alcohol). Not following the commitments of this contract results in premature discharge, alongside a discussion with a clinician about what went wrong for the purpose of future learning. | **Age group:** Adults (aged 18-60) & CYP (adolescents). Including in services specifically for CYP.  Individuals with recurrent self-harming and/or suicidal behaviour, with at least three symptoms of borderline personality disorder and a history of at least 7 days of admission to a psychiatric ward or presenting to a psychiatric emergency department at least three times, during the last 6 months. Exclusion criteria vary between services, but usually include: no regular contact with outpatient psychiatric services; unstable housing; somatic disorder or need or medication management that significantly contributes to inclusion criteria (284). | **Adult:**  RCT (283);  Qualitative literature (285–287,287);  Pre-post implementation study (288);  Other literature / intervention manual (284).  **CYP:**  RCT protocol (289). |
| Preventative admission model | **Locations include:** the Netherlands  **Sector:** Unknown  **Type of alternative:** Acute | In the Preventative Admission model, service users are offered a series of pre-arranged admissions over the next 6 months, instead of repeated negotiations in times of crises (usual care) (290). The frequency of admissions depends on the service user's previous inpatient service use. The aim is to give service users’ more control, reduce disagreements between service users and staff, and help service users delay crisis until the next scheduled hospitalisation, thereby improving their ability to cope in the community. | **Age group:** Adults (aged 18-60)  Adults with a diagnosis of borderline personality disorder and a history of repeated or long-term admissions. | Retrospective pre-post implementation pilot study (290). |
| HOPES model | **Locations include:** UK  **Sector:** Public  **Type of alternative:** Acute | HOPES is a clinical model based on principles of person-centred and human rights-based care. It involves an unconditional, relentlessly positive approach to reducing long-term segregation experienced by autistic adults, adults with an intellectual disability, and children and young people in inpatient mental health settings (291). | **Age group:** Adults & CYP. Including services specifically for CYP.  Children and young people, adults with an intellectual disability and autistic adults in inpatient settings at risk of long-term segregation in inpatient settings. | Grey literature (291)**.** |
| Inpatient wards for deaf people | **Locations include:** UK  **Sector:** Public  **Type of alternative:** Acute, long-term, forensic | Some specialist inpatient wards exist specifically for deaf adults and CYP with mental health problems. They provide assessment, treatment and interventions. They are staffed by deaf and hearing staff who can use British Sign Language. | **Age group:** Adults & CYP. Including services specifically for CYP.  People with a hearing impairment requiring psychiatric inpatient admission. Other eligibility criteria vary by service (e.g., some are acute wards, some secure, some for people with specific needs e.g., people with a diagnosis of personality disorder). | Quality standards (292) ;  Grey literature (293–295). |
| Inpatient wards for children under the age of 13 | **Locations include:** UK  **Sector:** Public  **Type of alternative:** Acute | Some inpatient psychiatric services exist specifically for children under the age of 13 years old. They provide intensive assessment and treatment for children under 13 who have complex emotional, behavioural and psychological difficulties. Whilst admitted, children can engage with schooling and take part in therapeutic activities. Parents/carers may also be offered support. | **Age group:** CYP (aged 5-13)  Children aged between 5-13 years old with complex mental health problems. Exclusion criteria include: a primary diagnoses of substance misuse, need for secure care in forensic services, children whose primary reason for referral is breakdown of family or placement, children who may present in a way that detrimentally impacts the care of other children in the service (e.g., extreme violence or behaviour) and young people who are unlikely to benefit on clinical grounds, and where the parents/caregivers or the child have not validly consented to the treatment If they are able to do so (296). | Grey literature (296)**.** |
| Specialist personality disorder wards | **Locations include:**  UK  **Sector:** Public, private  **Type of alternative:** Acute, long-term, forensic | Specialist inpatient wards for people with a diagnosis of personality disorder exist, including acute, longer-term rehabilitation, and secure services. They are staffed by multidisciplinary teams. They can provide assessment, medication, a range of evidence-based psychological therapies, occupational therapy, social therapy and physical healthcare. | **Age group:** Adults  Adults with a diagnosis of personality disorder who have complex mental health needs. | Pre-post studies (297–300);  Grey literature (301,302)**.** |
| Specialist eating disorder wards | **Locations include:** Widespread (e.g. UK, Norway, Italy, USA**)**  **Sector:** Public, private  **Type of alternative:** Acute | Specialist eating disorder units provide specialised inpatient care to people with eating disorders. They can provide: assessment; liaison with other services; expert oral refeeding; nasogastric tube feeding; evidence-based psychological interventions; and a daily programme of groups and activities (which may focus on areas such as motivational enhancement, psychoeducation, dietetic psychoeducation, emotional coping skills, independent living skills, social skills, and recreational and social activities). | **Age group:** Adults & CYP. Includes services specifically for CYP.  People with an eating disorder requiring more intensive input than outpatient treatment, especially those requiring specialist eating disorder input (e.g., refeeding, nasogastric tube feeding). Thresholds for admission vary across services. They are usually not able to accept patients with extreme medical risk and multiple organ failure (these individually are usually admitted to general medical hospitals instead) (303). Separate inpatient services exist for adults and children. | **Adult:**  Systematic reviews (78,79);  NICE guideline (303); RCT (86); Policy documents (304).  **CYP:**  Policy documents (305). |
| Specialist wards for early psychosis | **Locations include:**  UK, Canada  **Sector:** Public  **Type of alternative:** Acute | Specialist wards for early psychosis are usually offered as part of the early intervention services. These provide assessment, treatment and care which may include group psychotherapy programmes which focus on psychosis education, cognitive behavioural therapy skills, self-management, skill-building and recreation. | **Age group:** Adults & CYP (aged 16 and over)  People experiencing psychosis for the first time requiring inpatient admission. | Pre-post prospective cohort study (306);  Grey literature (307,308)**.** |
| Short-stay acute inpatient wards | **Locations include:** UK, USA, Canada  **Sector:** Public  **Type of alternative:** Acute | Short-stay acute inpatient wards (aka brief-stay wards, triage wards or assessment wards) are inpatient wards with shorter stays than acute wards (admitting patients for up to approximately a week). They usually do not admit patients detained under the Mental Health Act but can accept people detained under a 72-hour section (13). They aim to provide more intensive support than on standard acute psychiatric wards (including assessment, medication review, help with solving psychosocial problems and after-care planning), but do not provide distinctly different interventions to standard psychiatric wards (1). | **Age group:** Adults & CYP (aged 16 years and above). Includes services specifically for CYP.  Adults requiring acute admission, some specifically require a diagnosis of schizophrenia. Additional criteria for some brief-stay wards include: one or more previous admissions, stable housing, no current substance use or major medical problems, no diagnosis of brain injury, living with a responsible adult, admission from community, voluntary or on 72-hour section. | Systematic reviews (1);  RCTs (309,310);  Non-randomised trials (311);  Pre-post studies (6,312–314);  National surveys (13). |
| Open Dialogue | **Locations include:** Finland initially (now Europe [including the UK], USA, Australia)  **Sector:** Variable  **Type of alternative:** Acute | Open Dialogue is a therapeutic intervention as well as a way of organising services. It takes a person- and network-centred approach to treating mental health problems. It incorporates elements of systemic family therapy as well as psychodynamic principles. It emphasises the importance of transparent decision making and bringing together social and professional networks. Families are encouraged to meet immediately and regularly after a person has been referred to a service. The Open Dialogue approach aims to provide continuity of psychological care across services. It requires that services organise themselves so that they can facilitate immediate help, social network perspectives, flexibility and mobility, responsibility and psychological continuity (315). | **Age group:** Adults & CYP (aged 14 years and above)  Largely applied to individuals experiencing psychosis. | Systematic reviews (201);  Other evidence reviews (316–318);  Cluster RCT protocol (315);  Feasibility study (319);  Qualitative literature (320,321);  Pre-post implementation studies (322–325);  Other quantitative analysis (326);  Policy document (33). |
| Peer-supported open dialogue (POD) | **Locations include:** Finland, UK, Netherlands  **Sector:** Unknown  **Type of alternative:** Acute | Peer-supported open dialogue (POD) adds an explicit role of peer support workers to the Open Dialogue approach. These peer support workers are paid professionals with expertise gained through their own lived experience of psychological distress. This POD approach was introduced into daily ambulatory care for people diagnosed with severe mental health problems in five Dutch mental health care organisations (327). | **Age group:** Adults  Largely applied to individuals experiencing psychosis. | Qualitative literature (327). |
| Need-adapted treatment | **Locations include:** Finland, Sweden  **Sector:** Public  **Type of alternative:** Acute | Need-adapted treatment is a psychotherapeutic approach to schizophrenia. Therapeutic activities are tailored to each case so that they meet the changing needs of service users and family members. A psychotherapeutic approach to assessment and treatment is emphasised. It also highlights a family-centred initiation of treatment (328). Two examples of the application of need-adapted treatment in early psychosis include the Swedish Parachute Project and Acute Psychosis Integrated Treatment. | **Age group:** Adults and CYP (aged 15 and over).  Adults diagnosed with psychosis.  In the Swedish Parachute Project, people with a primary diagnosis of substance misuse or a diagnosed brain disorders were excluded.  In the Acute Psychosis Integrated Treatment study, exclusion criteria included previous treatment with neuroleptics, previous psychotherapy (more than 30 visits), serious physical illness, pregnancy, serious threat of suicide or violence (329). | General Pre-post implementation studies/ case time series (328,330).  Swedish Parachute Project Systematic reviews (21,201);  Non-randomised trial (331–333);  Pre-post design (334,335).  Acute Psychosis Integrated Treatment  Non-randomised trial (329)**.** |
| Specialist consultancy | **Locations include:**  UK  **Sector:** Private  **Type of alternative:** Acute, long-term, forensic | Some specialist consultancies exist which specifically aim to promote alternative approaches to standard inpatient care and to avoid and reduce inpatient mental health admissions. They may achieve this through direct clinical work or more structural approaches providing consultation, training and supervision to healthcare organisations. This consultancy may be provided by survivor-led, not-for profit or private organisations. Examples identified include the Leeds Survivor-Led Crisis Service (336,337), a ‘cultural consultation service’ (338), efforts of Killaspy et al. to reduce out-of-area long-term inpatient placements (339), co-PACT (340), ImROC (341) and Beam Consultancy (342). | **Age group:** Adults  Organisations providing specialist consultancy may work with a variety of people and organisations, including healthcare organisations, and some may also provide support directly to people in crisis. | Mixed-methods service evaluation (338);  Evaluation (339);  Grey literature (336,337,340–342). |
| Therapeutic Communities | **Locations include:** Widespread (e.g. UK, USA, Europe, Latin America)  **Sector:** Public, private, voluntary/ third sector  **Type of alternative:** Acute, long-term, forensic | Therapeutic Communities are structured, psychologically informed environments – they are places where the social relationships, structure of the day and joint activities together are all deliberately designed to help people’s health and well-being (343). Therapeutic communities are not defined by specific methods or programme elements, but the common shared values underlying them. They have been implemented in day centre and residential settings (including acute, long-term and forensic inpatient mental health wards). | **Age group:** Adults & CYP. Includes services specifically for CYP.  Anyone experiencing mental health problems. | **Adult:**  Systematic review (344); NICE reviews (345);  Other evidence synthesis (24);  Pre-post design (346–348);  Service evaluation/local audit (346,349,350);  Personal communication (Cockersell, 2022)^a^;  Qualitative literature (351);  Grey literature (343)**.**  **CYP:**  Other evidence synthesis (352,353);  Grey literature (354)**.** |
| Wraparound with intensive services | **Locations include:** USA  **Sector:** Variable  **Type of alternative:** Acute | Wraparound services are designed to help families develop a plan to address their child's individual needs at home and at school. The services are provided through teams that implement comprehensive support plans. These aim to link children, families or carers and their support networks with health, social services, education and youth justice services (355). | **Age group:** Adults & CYP (aged 0-20 years)  Children and adolescents with severe emotional and behavioural problems at-risk of out-of-home placement and their families, but also 'at risk' children. | Other evidence synthesis (58,75)**;**  RCTs (355). |
| Multisystemic therapy (MST) | **Locations include:** USA  **Sector:** Variable  **Type of alternative:** Acute, forensic | Multisystemic therapy (MST) is a model that was originally developed for treating serious antisocial behaviour in children and young people (aged 10-17 years old). MST teams provide a range of systemic interventions that address high risk personality traits associated with conduct disorder and antisocial personality disorder, as well as interventions to tackle offending behaviour, reparation work, interpersonal skills and family support (58). Interventions are provided at family's homes or at other community locations. It has been implemented with forensic and non-forensic populations. | **Age group:** CYP (aged 10 years and above)  Children with serious emotional disturbance who had been referred for emergency hospitalisation, who have access to a non-institutional residential environment (e.g., a family or relative's home, foster home). | Systematic review (356);  Other evidence synthesis (58,85);  RCTs (357–361);  Qualitative literature (362). |
| Enabling environments | **Locations include:**  UK  **Sector:** All sectors  **Type of alternative:** Acute, long-term, forensic | Enabling Environments are places where there is a focus on creating a positive and effective social environment and where healthy relationships are seen as the key to success. They can be found in any kind of service, and in a wide variety of sectors. The Royal College of Psychiatrists have published a set of standards to work towards in order to be accredited with an Enabling Environment. | **Age group:** Adults & CYP. Including specific CYP settings.  Enabling Environments can be applied in a wide variety of settings (including acute, forensic and long-term rehabilitation settings), and so there is no particular target population. | Grey literature (363). |
| Trieste | **Locations include:** Italy (with efforts made to implement in USA, Wales, Scotland, Japan)  **Sector:** Public  **Type of alternative:** Acute, long-term and forensic | In Trieste, Italy, there is a whole-systems approach to mental health care. When Trieste's psychiatric hospital closed in 1980, psychiatric institutions were replaced by a network of services that would be fully embedded in the community, highly accessible, and with a low threshold of access. There are no locked doors and restrictive care is avoided (364,365). | **Age group:** Adults & CYP.  All people with mental health needs in Trieste. There are no exclusion criteria around diagnoses, severity thresholds, etc. It is a whole-system approach intended to be able to cater to everyone's mental health needs. | Other evidence syntheses (365–369);  Pre-post studies (370,371);  Policy documents (372,373);  Qualitative literature (374,375);  Grey literature (364). |
| Buurtzorg model | **Locations include:** Netherlands, Switzerland, Germany, UK, Finland, USA  **Sector:** Public  **Type of alternative:** Acute | Buurtzorg (Dutch for 'neighbourhood care'), founded in the Netherlands, involves small, self-managed teams of nursing staff providing a range of personal, social and clinical care to people in their own homes in a particular neighbourhood. They aim to provide holistic, person-centred care. Buurtzorg T teams have been established which implement the Buurtzorg model in the treatment of psychiatric patients. These teams (which consist of a psychiatrist and multiple psychiatric nurses) can offer psychiatric patients mental health advice and treatment. | **Age group:** Adults & CYP  The Buurtzorg model can provide care to people with physical health needs or mental health needs. Specialist Buurtzorg T teams exist which provide psychiatric care. | Other evidence synthesis (376);  Qualitative literature (377,378);  Grey literature (379,380). |
| Geel family foster care model | **Locations include:** Belgium    **Sector:** Public  **Type of alternative:** Long-term | Geel is a family foster care model which has been practiced for centuries in a small town in Belgium. Guests often stay with families for long periods of time, with an average length of stay of approximately 30 years (381). After being placed with a foster family, the person receiving care becomes part of the foster family. They are encouraged to participate in the day-to-day running of the household (e.g., cleaning, cooking, engaging in other household tasks) according to their ability levels. The aim is to surround the person with normal expectations and demands in a compassionate environment. There is a team of district nurses who support these placements. During difficult times, they may visit daily. The community of Geel is a crucial part of the model (381). Foster families receive a monthly allowance of 500 euros (381). | **Age group:** Adults and CYP    People (all age groups) with mental health problems requiring residential support, often with chronic mental health difficulties (e.g. long-term psychotic, mood or personality disorder). A significant group of people receiving care have comorbid intellectual disabilities.  People are not eligible to receive care from the Geel foster care project if they are acutely unwell, displaying aggressive behaviour that is not reasonably under control, or if they have a history of sexual offences or serious crime (381). | Other evidence synthesis (381). |
| Healing Homes | **Locations include:** Sweden    **Sector:** Public  **Type of alternative:** Long-term | 'Healing homes' is a model in Sweden where people with psychosis are placed with host families for upwards of a year or two. It aims to help people with psychosis recover without medication. The person becomes a part of the family. Staff members offer clients intensive psychotherapy and provide supervision and support to host families (382). | **Age group:** Adults    Adults with psychosis who would like to be supported to recover from psychosis without medication. | Grey literature (382). |
| Intensive residential services (overarching category) | **Locations include:**  UK, Italy, USA, Australia, Spain, Sweden, Canada, New Zealand, Denmark, Germany  **Sector:** All sectors  **Type of alternative:** Long-term, forensic | Intensive residential services include ‘residential rehabilitation services’ and ‘intensive supported housing’ models, including ‘integrated supported housing’ and ‘housing with external intensive community support’ (e.g., Housing First and Full Service Partnership). They provide graduated support after discharge from hospital to the community. Intensive residential services aim to help people develop skills to enable more independent living. Individuals often move from a placement with higher to lower support every few years, with the ultimate aim of successfully managing an independent tenancy (383). | **Age group:** Adults & CYP. Includes services specifically for CYP and older adults.  People with complex, longer-term mental health problems and significant impairment of social functioning. Specific eligibility criteria varies by service. | Systematic reviews (13,384);  NICE reviews (345,385,386);  Other evidence syntheses (383);  Feasibility RCTs (387).  National surveys (388,389);  Retrospective cohorts (390);  Qualitative studies (391,392);  Other (393,394). |
| Residential rehabilitation services | **Locations include:**  UK, USA, Australia, Canada, Europe  **Sector:** All sectors  **Type of alternative:** Long-term, forensic | Residential rehabilitation services are communal facilities, staffed 24 hours a day, where day-to-day needs are provided (including meals, supervision of medicines and cleaning). Placements are not time-limited, and people do not hold a tenancy in a residential care home (190). | **Age group:** Adults & CYP. Includes services specifically for CYP and older adults.  People with complex, longer-term mental health problems and significant impairment of social functioning who require 24-hour residential care, but not inpatient admission. Specific eligibility criteria vary by service. | NICE reviews (190,385); Other evidence syntheses (383);  National surveys (389); Retrospective cohorts (390);  Cross-sectional studies (395);  Personal communication with Rethink Mental Illness 2023a^a^. |
| Residential rehabilitation services for eating disorders | **Locations include:**  UK, USA, Australia, Canada, Europe  **Sector:** Public, private  **Type of alternative:** Long-term | Residential rehabilitation services for people with eating disorders provide full-time housing and multidisciplinary treatment in a non-hospital-based setting. Support provided normally includes individual and group therapy, meal support and various forms of recreational activities (396). Unlike in inpatient eating disorder wards, residential rehabilitation services for people with eating disorders usually do not offer medical refeeding or monitoring (396). | **Age group:** Adults & CYP. Includes services specifically for CYP.  People with an diagnosed eating disorder who are medically stable but require more intensive support than outpatient treatment. | Systematic reviews (396). |
| Residential rehabilitation services for complex psychosis | **Locations include:**  UK  **Sector:** Public  **Type of alternative:** Long-term | Residential rehabilitation services for people with complex psychosis are communal facilities, staffed 24-hours a day, where residents’ day-to-day needs are provided for (including meals, supervision of medicines and cleaning). Placements are not time-limited. Residents do not hold a tenancy (190). | **Age group:** Adults  Adults with complex psychosis requiring 24-hour residential care, but not inpatient admission. | NICE reviews (190). |
| Integrated supported housing | **Locations include:**  UK, USA, Canada, Australia, Europe  **Sector:** All sectors  **Type of alternative:** Long-term, forensic | Integrated supported housing is shared or individual self-contained, time-limited tenancies with staff based on site up to 24 hours a day (190). Staff help residents to gain skills to move on to less supported accommodation. Integrated supported housing varies in intensity and nature of support. The intended length of stay for residents is two years, though only about a third of people move on in this time (190). | **Age group:** Adults  People with complex, longer-term mental health problems and significant impairment of social functioning. Some supported housing may be for specific sociodemographic groups (e.g., veterans, homeless people with serious mental illness, or dual serious mental illness and substance misuse). | Systematic reviews (13,397,398);  NICE guidelines (190);  Feasibility RCTs (399);  Cohort studies (390,400);  National surveys (389);  Qualitative literature (392);  Policy documents (401,402). |
| Housing First | **Locations include:** USA, Canada, Denmark, Finland, France, Italy, Sweden, Spain, UK, Czech Republic, Japan  **Sector:** All sectors  **Type of alternative:** Long-term, forensic | Housing First provides homeless people with mental health problems immediate access to a permanent tenancy with intensive outreach support from a specialist multidisciplinary mental health team (383). Compared to traditional floating outreach or supported housing models, in the Housing First model, workers have smaller caseloads (initial caseload of 5-7 people) which enables them to offer more intensive and personalised supported to service users (403). Support teams have two possible structures: intensive case management and assertive community treatment. | **Age group:** Adults  People who have experienced repeat housing instability, particularly those experiencing multiple disadvantage and experiencing, or at risk of, repeat homelessness. This can include people with offending histories. An unwillingness to take up an independent tenancy is regarded as a key exclusion criterion for Housing First services. | Systematic reviews (13)  NICE reviews (386);  Other evidence syntheses (383);  RCTs (404,405);  National surveys (389);  Policy documents (403,406). |
| Full Service Partnership | **Locations include:** USA  **Sector:** Voluntary/ third sector  **Type of alternative:** Long-term, forensic | Full Service Partnerships provide people with severe mental health difficulties who are homeless or at risk of homelessness with a combination of subsidised permanent housing and multidisciplinary team-based services with a focus on rehabilitation and recovery (407). They typically follow either an intensive case management model or a modified assertive community treatment model. They can provide medication management, vocational services, substance abuse services and other services to support clients (408). In this model, crisis intervention services are available 24/7. | **Age group:** Adults & CYP (aged 16 and over)  Eligibility criteria can vary by service, but generally the target population is adults with a serious mental and persistent mental illness that results in difficulty functioning and who are either homeless (or at risk of being homeless), justice-involved or high utilisers of mental health services. | Systematic reviews (13);  Quasi-experimental studies (407,408). |
| Specialist community mental health rehabilitation teams | **Locations include:**  UK, USA, Turkey, Israel, Sweden, Denmark, Singapore  **Sector:** Public and voluntary/ third sector  **Type of alternative:** Long-term | Community rehabilitation teams provide specialist skills and care coordination to identify and address people's rehabilitation needs in the community. Each person is assigned a designated care coordinator, though a shared team caseload approach is taken. The team should support and oversee people's progression through the rehabilitation pathway and liaise with other involved providers in the statutory and voluntary sectors (190). They also help people to manage transitions (e.g., as they move between different settings) aiming to reduce the chances of placement breakdown or relapse (409). | **Age group:** Adults  People over the age of 18, with complex mental health and social care needs who require longer-term multidisciplinary input. Teams commonly work with people living in supported accommodation, often over many years, though can work in all community settings. | Systematic reviews (13);  NICE guidelines (190);  Mixed-methods (410);  Policy documents (409,411). |
| Hostel-wards | **Locations include:**  UK  **Sector:** Public  **Type of alternative:** Long-term | Hostel-wards provide residential care to long-stay patients in inpatient mental health services who are not able to manage in the community. Hostel-wards provide domestic style accommodation but with staffing levels and support normally only found in a conventional hospital setting. Residents are legally inpatients in receipt of 24-hour nursing care. Residents are given a programme of domestic chores and self-care activities judged as being within their abilities, as well as personal target behaviours agreed with a psychologist. A proportion of residents, after a number of years, gain sufficient independence to move to less supported accommodation in the community. | **Age group:** Adults  Adults below the age of 65 with psychiatric disorders, already in standard inpatient care hospital for over 6 months - "long-stay patients". | Systematic review (412);  Non-randomised trials (413). |
| Housing and Accommodation Support Services (HASS) | **Locations include:**  UK  **Sector:** Public and voluntary/ third sector  **Type of alternative:** Forensic | Housing and Accommodation Support Services (HASS) form part of the Offender Personality Disorder (OPD) pathway. HASS provide support and accommodation to people on probation who have been released from prison and secure healthcare settings or who are moving on from Approved Premises. Residents are offered weekly key work sessions, house meetings and a range of group interventions and activities to help them develop living skills, promote socialisation, build confidence and establish positive community links. | **Age group:** Adults  Offenders with likely Personality Disorder who have been screened into the Offender Personality Disorder (OPD) pathway. The service is for people who would benefit from high levels of specialist support to enable successful resettlement. There may be eligibility criteria such as minimum remaining lengths of time on licence (e.g., 12 months). | Policy documents (414); Grey literature (415,416). |
| Residences for the Execution of Security Measures (REMS) | **Locations include:** Italy  **Sector:** Public  **Type of alternative:** Forensic | Residences for the Execution of Security Measures (REMS) are regional-based community residential facilities in Italy for people who have committed crimes who have serious mental health difficulties. These individuals are given custodial security orders and serve the period of admission at REMS. Each facility accommodates no more than twenty people, and they are designed to have a therapeutic/rehabilitative purpose. Duration of treatment is limited and there is an absence of police officers (417). | **Age group:** Adults  REMS are for people who have committed crimes who also have serious mental health difficulties (e.g., psychotic-spectrum disorders, major depression, personality disorders), requiring high-intensity psychiatric treatment (418). | Cross-sectional or descriptive studies (417–419). |
| Secure children’s homes | **Locations include:**  UK  **Sector:** Public and voluntary/ third sector  **Type of alternative:** Long-term, forensic | Secure children's homes include full residential care, formal on-site education, and healthcare provision. They operate with high staffing ratios. They aim to support residents to develop their personal skills and support them to be able to manage safely and responsibly on exit. They offer residents input from a range of mental health and associated professionals to help them to reintegrate into the community successfully (420). | **Age group:** CYP (aged 10-17 years)  CYP who are sentenced or on remand through the justice system or placed due to local authority concerns that they are a serious risk to themselves or others and there is no other type of placement that could keep them safe. Some secure children’s homes are welfare-only (where CYP are placed for their own welfare by their local council and the court), whereas others are for sentenced young people via the Youth Custody Service, some accept both. | Other evidence reviews (421);  National audits (422);  Policy documents and reports (423–426);  Grey literature (420,427). |
| Secure training centres | **Locations include:**  UK  **Sector:** Usually private, sometimes public  **Type of alternative:** Forensic | Secure training centres are places of detention for young people who are vulnerable and have been sentenced to custody following conviction of a criminal offence, or who are being held on remand awaiting trial. They are usually run by private companies, though the Prison and Probation Service can take over management. Children should receive 30 hours of education and training each week and follow a school-day timetable (428). | **Age group:** CYP (12-17 years of age)  Children who have been sentenced to custody following conviction of a criminal offence, who are a significant risk to themselves or others, and are more vulnerable than those in Youth Offender Institutions but less than in secure children's homes. | Other evidence reviews (421);  Policy documents (423,425);  Grey literature (428). |
| Forensic community mental health teams | **Locations include:**  UK  **Sector:** Public  **Type of alternative:** Forensic | Core functions of the forensic community mental health teams include: providing case management of a defined caseload (especially those leaving secure care); providing treatment; providing liaison, advice, specialist interventions, educational and skills development; active involvement in care pathway management into and out of secure settings and prison; liaison with Multiagency Public Protection Arrangements and links with criminal justice liaison and court diversion services (429). They are staffed by multidisciplinary teams. Individuals have care co-ordinators with caseloads of 12-20 patients. Case management may be longer-term (429). There are two main models for delivering specialist forensic care: parallel and integrated (429). | **Age group:** Adults  High risk offenders with mental health problems who require support transitioning from forensic inpatient care or prisons back into the community or who require support in the community. | Other evidence syntheses (429);  Grey literature (430,431). |
| Forensic assertive community treatment | **Locations include:** USA, Canada, The Netherlands, New Zealand, Belgium  **Sector:** Variable  **Type of alternative:** Forensic | Forensic assertive community treatment aims to reduce criminal justice involvement and reoffending among justice-involved individuals with mental health difficulties (432). It is an adaptation of the 'assertive community treatment' model which attempts to also address criminogenic risks and needs. It involves the provision of crisis assessment and intervention, pharmacotherapy and behavioural interventions, recovery and social support services, vocational and housing services, and substance use interventions. It may also involve coordination with criminal justice entities (e.g., law enforcement, courts, jails, community corrections), provision of legal advocacy and assistance and support with applying for financial support (432). | **Age group:** Adults  Eligibility varies depending on the team, e.g., mental health diagnosis and duration, status of not guilty by reason of insanity, types of crimes, history of repeated incarceration, co-occurring substance use disorders, history of non-adherence to treatment etc. | Other evidence synthesis (433,434);  Qualitative literature (435);  Policy documents (432). |
| Adolescent forensic community services | **Locations include:**  UK  **Sector:** Public  **Type of alternative:** Forensic | Community adolescent forensic services provide assessment and treatment interventions for young people with a high risk of offending behaviour and experiencing mental distress. These interventions can be provided in community, residential and custodial settings. They may operate as dedicated multidisciplinary teams, provide in-reach to juvenile secure estate and local authority secure children's homes or include special interest or expertise as part of CYP mental health services (58). | **Age group:** CYP (adolescents)  Young people with complex, persistent or serious mental disorders who are in contact with the criminal justice system or who have a high risk of offending behaviour. | Narrative review (58); Policy document (436);  Grey literature (430). |
| Intensive Intervention and Risk Management Services (IIRMS) | **Locations include:**  UK  **Sector:** Public  **Type of alternative:** Forensic | Intensive intervention and risk management services (IIRMS) represent the community intervention part of the Offender Personality Disorder (OPD) pathway. They adopt a psychologically-informed case management approach and focus on direct work with individuals who are being released from prison into the community on probation license and who have a history of high risk, high harm violent convictions linked to pervasive psychological and interpersonal problems (437). Approaches to engagement are informed by psychological formulations which consider individuals' personality styles, relational issues and possible triggers. IIRMS provide in-reach to prison and outreach support in the community, with opportunities for psychoeducational learning. They aim to enhance skills and self-management necessary for successful resettlement (437). | **Age group:** Adults  Offenders with likely personality disorder who were screened into the OPD pathway, who are being released from prison into the community on probation licence and who have a history of high-risk, high harm violent convictions linked to pervasive psychological and interpersonal problems. | Descriptive literature (437). |
| Transitional support and liaison services | **Locations include:** UK  **Sector:** Public  **Type of alternative:** Forensic | This model is a specific form of IIRMS. These services provide support over a few months to help people access services and support provided by statutory and voluntary organisations, both during and after their transition from custody to the community (438). It forms part of the OPD pathway. | **Age group:** Adults  Offenders with likely personality disorder who have been screened into the OPD pathway and who are transitioning from custody into the community. | Service evaluation (438). |
| Ter Beschikking Stelling (TBS) | **Locations include:** The Netherlands  **Sector:** Public  **Type of alternative:** Forensic | In The Netherlands, offenders with severe mental disorders with high risk of recidivism can be detained in specialist high-secure forensic psychiatric institutions (so-called ‘TBS hospitals’) after they have served their prison sentence. The main aims of this penal hospital order are protection of society and treatment of the offender, in order to rehabilitate patients into the community (439). Key differences between the Dutch and English forensic inpatient care systems include: the ability of the Dutch system to detain people with substance-related disorders only under mental health legislation; risk being a specific criterion for detention; and TBS hospitals having multiple wards with different levels of security within the same hospital, meaning that offenders remain under the same clinical team as they transition between different levels of security, in contrast to England, where offenders are transferred between different hospitals. The Dutch system is also less restrictive (e.g., allowing more patients unsupervised leave and family visits) (440). | **Age group:** Adults  Adult offenders who have a mental disorder who have been declared partially or fully unaccountable for a serious sexual or violent offence and who have been judged to have a high risk of recidivism and so have been given a penal hospital order. | Historical/ narrative reviews (439)  Cross-sectional studies (440). |
| Democratic therapeutic communities | **Locations include:**  UK  **Sector:** Public  **Type of alternative:** Acute, long-term, forensic | Democratic therapeutic communities (DTCs) represent a whole-system approach to treatment or rehabilitation (441). It refers to an institution (or a unit within an institution), including its staff, residents and all rules, processes and activities that take place within it (441). They operate according to four principles: democratisation, communalism, permissiveness and reality confrontation (441). It usually encompasses all 24-hours of the day. Specific treatment programmes (e.g., CBT) may be delivered within the overall context of a therapeutic community. The involvement of the peer group is an essential and defining feature of the DTC approach. It is expected that residents become involved in each other's treatment, becoming 'auxiliary therapists' (441). | **Age group:** Adults & CYP.  Offenders in prisons and forensic psychiatric settings in high and medium security. | Other evidence syntheses (441,442);  RCTs (443);  Non-randomised trials (444);  Pre-post designs (445). |
| Psychologically Informed Planned Environments (PIPES) | **Locations include:**  UK  **Sector:** Public  **Type of alternative:** Forensic | Psychologically Informed Planned Environments (PIPEs) aim to support the progression of offenders, as part of the Offender Personality Disorder (OPD) pathway. They have a particular focus on the environment and on recognising the importance and quality of relationships and interactions. In order to help create a safe and supportive environment, staff members receive additional training to develop an increased psychological understanding of their work. To help approach situations in a psychologically informed way, six key components are utilised: developing an 'Enabling Environment', a combination of structured groups between staff and residents, focused PIPE keywork, socially creative sessions and training, and supervision and reflective sessions for staff (446). | **Age group:** Adults  Offenders who have complex needs and personality related difficulties and have been screened into the Offender Personality Disorder (OPD) pathway and are in prisons or Approved Premises (community-based hostel settings which support people who have just been released from custody). | Policy document/service evaluation (446). |
| Offender Personality Disorder treatment services | **Locations include:**  UK  **Sector:** Public  **Type of alternative:** Forensic | OPD treatment services are another component of the OPD pathway in the UK. They offer support to people in secure settings or individuals in the community who are subject to probation supervision. Support offered may include Mentalisation Based Treatment for people with Anti-Social Personality Disorder (MBT-ASPD) or Male-Trauma Recovery Empowerment Model (M-TREM) services. MBT-ASPD service users attend weekly group therapy sessions and receive additional monthly individual therapy sessions. Whereas, M-TREM services offer an 18-week educational and discussive group for men who have a history of trauma and who are currently supervised by the National Probation Service. The programme strongly considers the influence of male gender roles and stereotypes and how these might be related to trauma experiences, as well as coping styles an individual strengths and strategies to build upon these coping skills (447). | **Age group:** Adults  Offenders who have been screened into the OPD pathway who are in secure settings (including prison or forensic inpatient services) or who are in the community and subject to probation supervision. | Grey literature (447,448). |
| Psychologically Enhanced Resettlement Services (PERS) | **Locations include:**  UK  **Sector:** Public  **Type of alternative:** Forensic | Pathways Enhanced Resettlement Services (PERS) are one component of the OPD pathway. Their core function is to support OPD service users through a programme with three phases: 'Transition-In', 'Graduation' and 'Transition-Out'. These phases aim to support service users as they transition in and out of open conditions. Service users have the opportunity to consolidate skills through a rolling psycho-educational group programme, to engage in regular key work, and to use the 'open-door' policy for ad hoc support from staff (449). | **Age group:** Adults  PERS are intended for people who are: Category D prisoners; managed by the National Probation Service; subject to restricted Release on Temporary License; assessed as posing a high or very high risk of harm to others; and who have personality difficulties that meet the Offender Personality Disorder screening criteria. | Qualitative literature (449). |

^a^ Personal communication, offering further information on service models, were provided during the expert consultation phase in either written or oral format. Shading has been used to indicate models that group together: the first darker-shade row in each block indicates the over-arching category, whilst the lighter-shade rows underneath indicate models within that over-arching category. ACT; Assertive Community Treatment, ADU; Acute Day Unit, BA; Brief Admission, CAMHS; Child and Adolescent Mental Health Services, CMHT; Community Mental Health Team, CBT; Cognitive Behavioural Therapy, CCM; Crisis Case Management, CRHTT; Crisis Resolution and Home Treatment Teams, CYP; Children and Young People, DTC; Democratic Therapeutic Communities, EIP; Early Intervention in Psychosis, FACT; Flexible Assertive Community Treatment, FREED; First episode and Rapid Early intervention for Eating Disorders, HASS; Housing and Support Services, ICM; Intensive Case Management, IIRMS; Intensive Intervention and Risk Management Services; LGBTQ+; Lesbian, Gay, Bi, Trans, Queer, Questioning and Ace, MBT-ASPD; Mentalisation Based Treatment for people with Anti-Social Personality Disorder, M-TREM; Male-Trauma Recovery Empowerment Model, MST; Multisystemic Therapy, NHS; National Health Service, NICE; National Institute for Care Excellence, OPD; Offender Personality Disorder, PACER; Police Ambulance Crisis Response, PAM; Psychiatric Emergency Response, PA; Preventative Admission, PERS; Pathways Enhanced Resettlement Services, PIPEs; Psychologically Informed Planned Environments, POD; Peer-Supported Open Dialogue, RAID; Rapid, Assessment, Interface and Discharge, RCT; Randomised Controlled Trial, REMS; Residences for the Execution of Security Measures, SDS; Supported Discharge Service, STAR; Support Team Assisted Response, STC; Secure Training Centre, TBS; Ter Beschikking Stelling, TCs; Therapeutic Communities, UK; United Kingdom, USA; United States of America

17.	Therapeutic alternatives to hospital [Internet]. Tŷ Cynnal Support House & Linden House. n.d. Available from: https://platfform.org/project/alternatives-to-hospital/

18.	SAMHSA. Substance Abuse and Mental Health Services Administration: National Guidelines for Child and Youth Behavioral Health Crisis Care [Internet]. Substance Abuse and Mental Health Services Administration; 2022. Available from: https://store.samhsa.gov/sites/default/files/SAMHSA_Digital_Download/pep-22-01-02-001.pdf

19.	The Hope Service ©. The HOPE Service [Internet]. 2023. Available from: http://www.hopeservice.org.uk/

20.	Future Health and Social Care. Birmingham and Solihull Mental Health Foundation Trust, Forward Thinking Birmingham and Future Health and Social Care: Crisis House. Future Health and Social Care website. Available from: https://www.futurehsc.com/crisis-house/

34.	Pathways Vermont. SOTERIA HOUSE [Internet]. n.d. Available from: https://www.pathwaysvermont.org/what-we-do/our-programs/soteria-house/

35.	Soteria Bradford. Soteria Bradford [Internet]. n.d. Available from: https://soteriabradford.wixsite.com/soteria-bradford

36.	Charity Commission. Soteria Network: Charity Commission Decision dated 29 February 2021 [Internet]. Available from: https://assets.publishing.service.gov.uk/government/uploads/system/uploads/attachment_data/file/335438/soteria_dec.pdf

37.	Ciompi L, Mosher LR. SOTERIA CRITICAL ELEMENTS [Internet]. n.d. p. 1–2. Available from: https://psychrights.org/education/SoteriaCriticalElements.pdf

50.	Canterbury Medical Research Foundation. Mental Health Research. Evaluating a peer-led acute community residential service for severe mental illness. Available from: https://cmrf.org.nz/research/evaluating-a-peer-led-acute-community-residential-service-for-severe-mental-illness/

56.	A guide to Shared Lives for mental health crisis [Internet]. Wales, United Kingdom: Shared Lives South East Wales; n.d. Available from: https://www.caerphilly.gov.uk/caerphillydocs/adults-and-older-people/shared-lives/15535-leaflet-1-e.aspx

62.	Multidimensional treatment foster care program components and principles of practice. In: Treating chronic juvenile offenders: Advances made through the Oregon Multidimensional Treatment Foster Care Model. Washington DC: American Psychological Association.; n.d.

87.	Maudsley Centre for Child and Adolescent Eating Disorders. The Intensive Treatment Program [Internet]. Available from: https://mccaed.slam.nhs.uk/young-person-and-families/our-services/intensive-treatment-program/

115.	MIND. Listening to Experience: An Independent Inquiry into Acute and Crisis Mental Healthcare. London: MIND; 2011.

124.	Bradford S. The Building Blocks of Social Work Practice. In: 9th edition. Pearson Education (US); 2011.

132.	People USA. Forensic Mobile Crisis Response Team [Internet]. Available from: https://people-usa.org/program/forensic-mobile-team/

141.	Edmondson D, Cummins I. Oldham Mental Health Phone Triage/RAID Pilot Project. Evaluation Report. Manchester: Manchester Metropolitan University: University of Salford; 2014.

147.	Climer BA, Gicker B. Psychiatric Times. CAHOOTS: A Model for Prehospital Mental Health Crisis Intervention. [Internet]. Available from: https://www.psychiatrictimes.com/view/cahoots-model-prehospital-mental-health-crisis-intervention

153.	Association to Benefit Children (ABC). Children’s Mobile Crisis Team [Internet]. Available from: https://www.a-b-c.org/store/crisis-intervention

154.	Health Innovation Network, NHS England. Evaluating NHS Mental Health Crisis Hubs in London: Final Report [Internet]. 2022. Available from: https://healthinnovationnetwork.com/wp-content/uploads/2022/11/MH-Crisis-Hubs-Evaluation-Final-Report.pdf

155.	Northover G. Children and Young People’s Mental Health Services GIRFT Programme National Specialty Report [Internet]. 2022. Available from: https://future.nhs.uk/connect.ti/GIRFTNational/view?objectId=130556421

156.	NHS North Central London Integrated Care Board. Child & Adolescent Mental Health Service [Internet]. n.d. Available from: https://gps.northcentrallondon.icb.nhs.uk/services/child-and-adolescent-mental-health-service-camhs-islington

162.	Woden Community Service. Woden Community Service [Internet]. Available from: https://www.wcs.org.au

163.	New York Association of Psychiatric Rehabilitation Services Inc. Peer Bridger Project [Internet]. Available from: https://www.nyaprs.org/peer-bridger

187.	CENTRUM FÖR EVIDENSBASERADE PSYKOSOCIALA INSATSER (CEPI). Ups and Downs in Mental Health-A new flexible care and support model for increased health and functioning in everyday life among people with alarming and complex mental health need [Internet]. Available from: https://www.cepi.lu.se/delta-i-studie

199.	West London NHS Trust. Early Intervention in Psychosis [Internet]. n.d. Available from: https://www.westlondon.nhs.uk/our-services/adult/mental-health-services/early-intervention-psychosis

205.	Northover G. Getting It Right First Time (GIRFT): Mental Health - Children and Young People’s Services. 2022.

213.	FREED. Free from Eating Disorders (FREED) [Internet]. n.d. Available from: https://freedfromed.co.uk/

214.	FREED From ED [Internet]. 2023. FREED Shared Learning Conference Slides. Available from: https://freedfromed.co.uk/news-and-stories/73/freed-network-shared-learning-conference-march-2023

215.	FREED From ED. FREED Shared Learning Conference Video (Vimeo) [Internet]. Available from: https://vimeo.com/showcase/10321714

231.	State of Tennessee Department of Mental Health & Substance Abuse Services website [Internet]. n.d. Available from: https://www.tn.gov/behavioral-health/need-help/crisis-services/csu.html

249.	Central & North West London NHS Foundation Trust. Community Eating Disorder Service for Children and Young People [Internet]. Available from: https://www.cnwl.nhs.uk/services/mental-health-services/eating-disorders/community-eating-disorder-service-children-and-young-people

278.	The Sanctuary Institute [Internet]. Available from: https://www.thesanctuaryinstitute.org/about-us/the-sanctuary-model/

284.	Liljedahl SI, Helleman M, Daukantaitė D, Westling S. Brief Admission: Manual for training and implementation developed from the Brief Admission Skåne Randomized Controlled Trial (BASRCT). Lund, Sweden: Media-Tryck, Lund University.; 2017.

291.	Mersey Care NHS Foundation Trust website [Internet]. n.d. Available from: https://www.merseycare.nhs.uk/hopes-model

292.	Souza R, Palmer L, Tarant E, Kaselionyte J, editors. Standards for Adult Inpatient Mental Health Services for Deaf People [Internet]. The Royal College of Psychiatrists; 2015. Available from: https://www.rcpsych.ac.uk/docs/default-source/improving-care/ccqi/quality-networks/deaf-services-qnmhd/qnmhd_standards_2nd_edition_2015.pdf?sfvrsn=b9b9be33_2

293.	South West London and St George’s Mental Health NHS Trust [Internet]. n.d. Corner House - National Deaf CAMHS. Available from: https://www.swlstg.nhs.uk/our-services/find-a-service/service/corner-house-national-deaf-camhs#:~:text=Corner%20House%20is%20a%20six,complex%20emotional%20and%20psychological%20problems.

294.	Novakova L. Providing effective mental health services for deaf and hard of hearing patients [Internet]. 2020. Available from: https://www.england.nhs.uk/blog/providing-effective-mental-health-services-for-deaf-and-hard-of-hearing-patients/

295.	Cygnet Group website [Internet]. n.d. Available from: https://www.cygnetgroup.com/locations/cygnet-hospital-bury/bridge-hampton-mens-low-secure-and-deaf-mental-health/

301.	Cygnet Health Website [Internet]. n.d. Available from: https://www.cygnethealth.co.uk/services/personality-disorder/

302.	West London NHS Trust Website [Internet]. n.d. Available from: https://www.westlondon.nhs.uk/our-services/adult/mental-health-services/cassel-hospital

303.	Eating disorders: recognition and treatment [NICE Guideline No. 69] [Internet]. The National Institute for Health and Care Excellence; 2020. Available from: https://www.nice.org.uk/guidance/ng69

304.	NHS Standard Contract for Specialised Eating Disorders (Adults): Schedule 2 - The Services - Service Specifications [Internet]. NHS England; 2013. Available from: https://www.england.nhs.uk/commissioning/wp-content/uploads/sites/12/2014/12/c01-spec-eat-dis-1214.pdf

305.	Batchelor A. Adult Critical Care: Getting It Right First Time (GIRFT) Programme National Specialty Report. National Health Service; 2021.

306.	Siebert S, Leopold K, Baumgardt J, von Hardenberg LS, Burkhardt E, Bechdolf A. Specialized inpatient treatment for young people with early psychosis: acute-treatment and 12-month results. Eur Arch Psychiatry Clin Neurosci. 2022 Oct;272(7):1–14.

307.	South London and Maudsley NHS Foundation Trust website [Internet]. n.d. Available from: https://slam.nhs.uk/service-detail/service/leo-unit-early-intervention-27/

308.	CAMH. Crisis and Critical Care Unit 5 Early Psychosis Unit (EPU - CCC5) [Internet]. n.d. Available from: https://www.camh.ca/en/your-care/programs-and-services/crisis-and-critical-care-unit-5#:~:text=The%20Early%20Psychosis%20Unit%20(EPU)%20is%20a%20specialist%20inpatient%20unit,mental%20health%20issues%20with%20psychosis.

336.	Leeds Survivor-Led Crisis Service website [Internet]. n.d. Available from: https://www.lslcs.org.uk/additional-services/

337.	Leeds Survivor Led Crisis Services: Trustees’ Report and Financial Statements for the Year Ended 31 March 2018 [Internet]. Leeds Survivor Led Crisis Services; 2018. Available from: https://register-of-charities.charitycommission.gov.uk/charity-search?p_p_id=uk_gov_ccew_onereg_charitydetails_web_portlet_CharityDetailsPortlet&p_p_lifecycle=2&p_p_state=maximized&p_p_mode=view&p_p_resource_id=%2Faccounts-resource&p_p_cacheability=cacheLevelPage&_uk_gov_ccew_onereg_charitydetails_web_portlet_CharityDetailsPortlet_objectiveId=A9335399&_uk_gov_ccew_onereg_charitydetails_web_portlet_CharityDetailsPortlet_priv_r_p_mvcRenderCommandName=%2Faccounts-and-annual-returns&_uk_gov_ccew_onereg_charitydetails_web_portlet_CharityDetailsPortlet_priv_r_p_organisationNumber=3956536

338.	Bhui KS, Owiti JA, Palinski A, Ascoli M, De Jongh B, Archer J, et al. A cultural consultation service in East London: Experiences and outcomes from implementation of an innovative service. Int Rev Psychiatry. 2015 Jan 2;27(1):11–22.

339.	Killaspy H, Rambarran D, Harden C, Fearon D, Caren G, McClinton K. A comparison of service users placed out of their local area and local rehabilitation service users. J Ment Health. 2009 Jan;18(2):111–20.

340.	University of Oxford website [Internet]. Co-PACT summary. Available from: https://www.psych.ox.ac.uk/research/chimes/co-pact/co-pact_summary

341.	ImROC [Internet]. n.d. Available from: https://imroc.org/resource/co-production-sharing-our-experiences-reflecting-on-our-learning/

348.	Solís-Román C, Knickman J. Project to evaluate the impact of Fountain House programs on Medicaid utilization and expenditures. Health Evaluation and Analytics Lab: New York University; 2016.

380.	Treatment at home for psychological and psychiatric problems [Internet]. The Buurtzorg Model. Available from: https://www.buurtzorg.com/innovation/buurtzorg-te/

411.	Kalidindi S, Killaspy H, Edwards T. Community psychosis services: the role of community mental health rehabilitation teams: Faculty Report. Royal College of Psychiatrists; 2012 Nov.

412.	Macpherson R, Edwards TR, Chilvers R, David C, Elliott HJ. Twenty-four hour care for schizophrenia. Cochrane Schizophrenia Group, editor. Cochrane Database Syst Rev [Internet]. 2009 Apr 15 [cited 2023 Jun 23]; Available from: https://doi.wiley.com/10.1002/14651858.CD004409.pub2

413.	Hyde C, Bridges K, Goldberg D, Lowson K, Sterling C, Faragher B. The Evaluation of a Hostel Ward: A Controlled Study using Modified Cost-Benefit Analysis. Br J Psychiatry. 1987 Dec;151(6):805–12.

414.	NHS England. The offender personality disorder pathway strategy. National Offender Management Service. [Internet]. 2015. Available from: https://www.england.nhs.uk/commissioning/wp-content/uploads/sites/12/2016/02/opd-strategy-nov-15.pdf

415.	Social Interest Group. Penrose OPD HASS [Internet]. Available from: https://socialinterestgroup.org.uk/our-services/penrose-opd-hass-catford/

423.	Youth Justice Board. Developing the Secure Estate for Children and Young People in England and Wales–Young People’s Consultation Report. [Internet]. Youth Justice Board; Available from: https://dera.ioe.ac.uk/14532/1/Young%20Peoples%20Consultation%20Report.pdf

424.	UNLOCKING THE FACTS: YOUNG PEOPLE REFERRED TO SECURE CHILDREN’S HOMES: SUMMARY REPORT [Internet]. Cardiff University: What Works for Children’s Social Care; 2020 Dec. Available from: https://whatworks-csc.org.uk/wp-content/uploads/WWCSC_Unlocking_the_Facts_SCH_summary_report_Dec2020_Acc.pdf

425.	Ministry of Justice. National Audit Office: Children in custody: secure training centres and secure schools. [Internet]. 2022. Available from: https://www.nao.org.uk/wp-content/uploads/2022/04/Children-in-custody-secure-training-centres-and-secure-schools.pdf

426.	Hart D, La Velle I. Secure children’s homes: placing welfare and justice children together. [Internet]. 2021. Available from: https://assets.publishing.service.gov.uk/government/uploads/system/uploads/attachment_data/file/983619/Secure_children_s_homes_placement_review_report.pdf

427.	Nuffield Family Justice Observatory. Number of applications to deprive children of their liberty in unregulated placements rises by 462% in three years [Internet]. 2022. Available from: https://www.nuffieldfjo.org.uk/news/number-of-applications-to-deprive-children-of-their-liberty-in-unregulated-placements-rises-by-463-in-three-years

428.	Family Rights Group. Helping Families Helping Children [Internet]. Available from: https://frg.org.uk

429.	Latham R, Williams HK. Community forensic psychiatric services in England and Wales. CNS Spectr. 2020 Oct;25(5):604–17.

430.	West London NHS Trust. Forensic Community Services [Internet]. Available from: https://www.westlondon.nhs.uk/our-services/adult/mental-health-services/forensic-community-services

431.	Kent and Medway NHS and Social Care Partnership Trust. Forensic Outreach and Liaison service [Internet]. Available from: https://www.kmpt.nhs.uk/our-services/forensic-outreach-and-liaison-service/

432.	Substance Abuse and Mental Health Services Administration. Forensic Assertive Community Treatment (FACT): A Service Delivery Model for Individuals with Serious Mental Illness Involved With the Criminal Justice System [Internet]. Substance Abuse and Mental Health Services Administration (SAMHSA); n.d. Available from: https://store.samhsa.gov/sites/default/files/d7/priv/pep19-fact-br.pdf

438.	O’Meara A, Morgan L, Godden S, Davies J. A model of a specialist transitional support and liaison service within the Offender Personality Disorder Pathway in Wales: Learning from a regional pilot service. J Community Crim Justice. 2019;

447.	Midlands Partnership University NHS Trust. Forensic Mental Health: Offender Personality Disorder Pathways [Internet]. Available from: https://forensics.mpft.nhs.uk/offender-personality-disorder-pathways

448.	Craissati J, Joseph N, Skett S, editors. Working with offenders with personality disorder: A practitioners guide [Internet]. NHS England; 2015. Available from: https://www.england.nhs.uk/commissioning/wp-content/uploads/sites/12/2015/10/work-offndrs-persnlty-disorder-oct15.pdf

449.	Jarvis D, Shaw J, Lovell T. Service user experiences of a psychologically enhanced resettlement service [PERS] in an English open prison. J Forensic Pract. 2022 Jun 28;24(3):241–52.
