## Supplementary figures and images for "Alternative approaches to standard inpatient mental health care: development of a typology of service models"

### Additional File 4

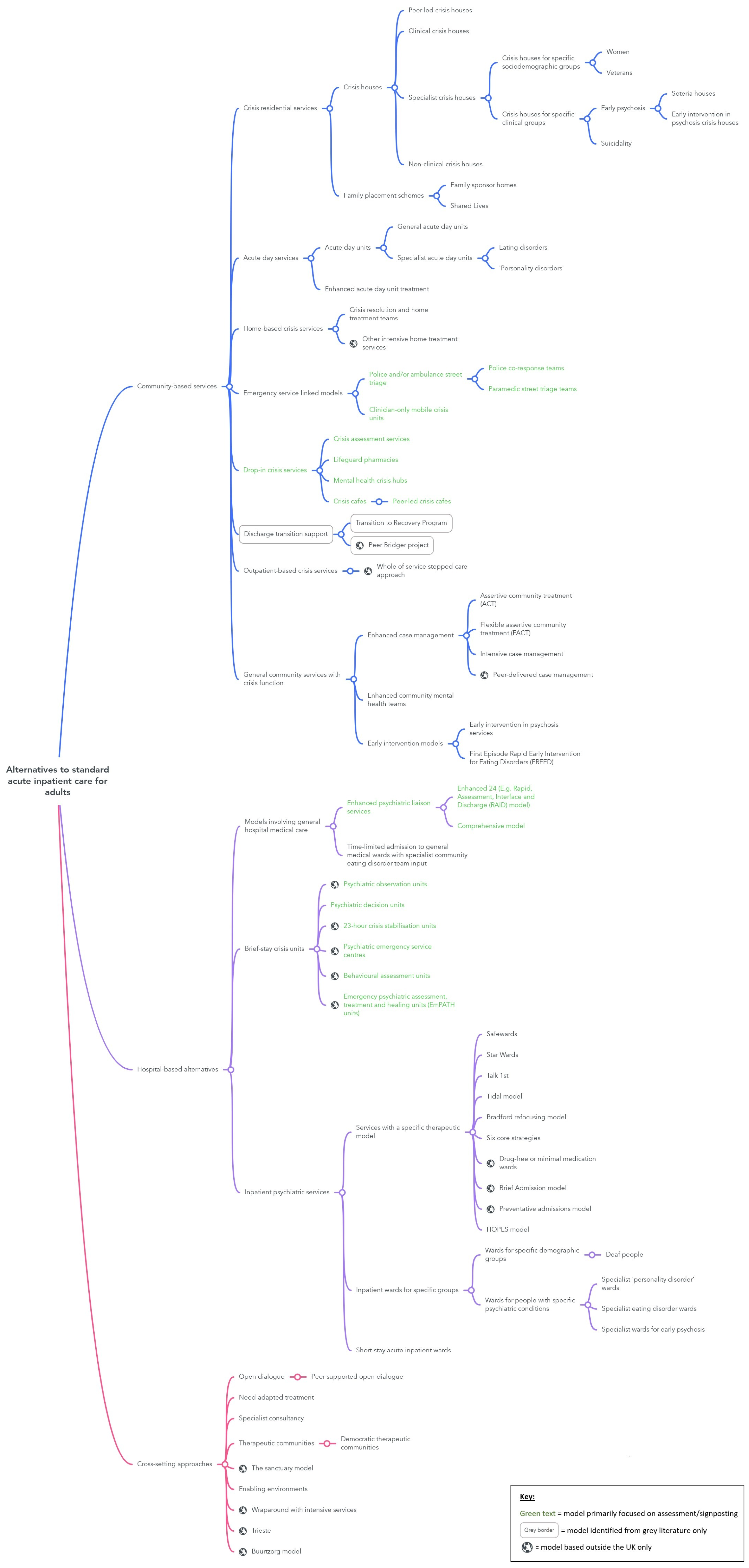

### Additional File 5

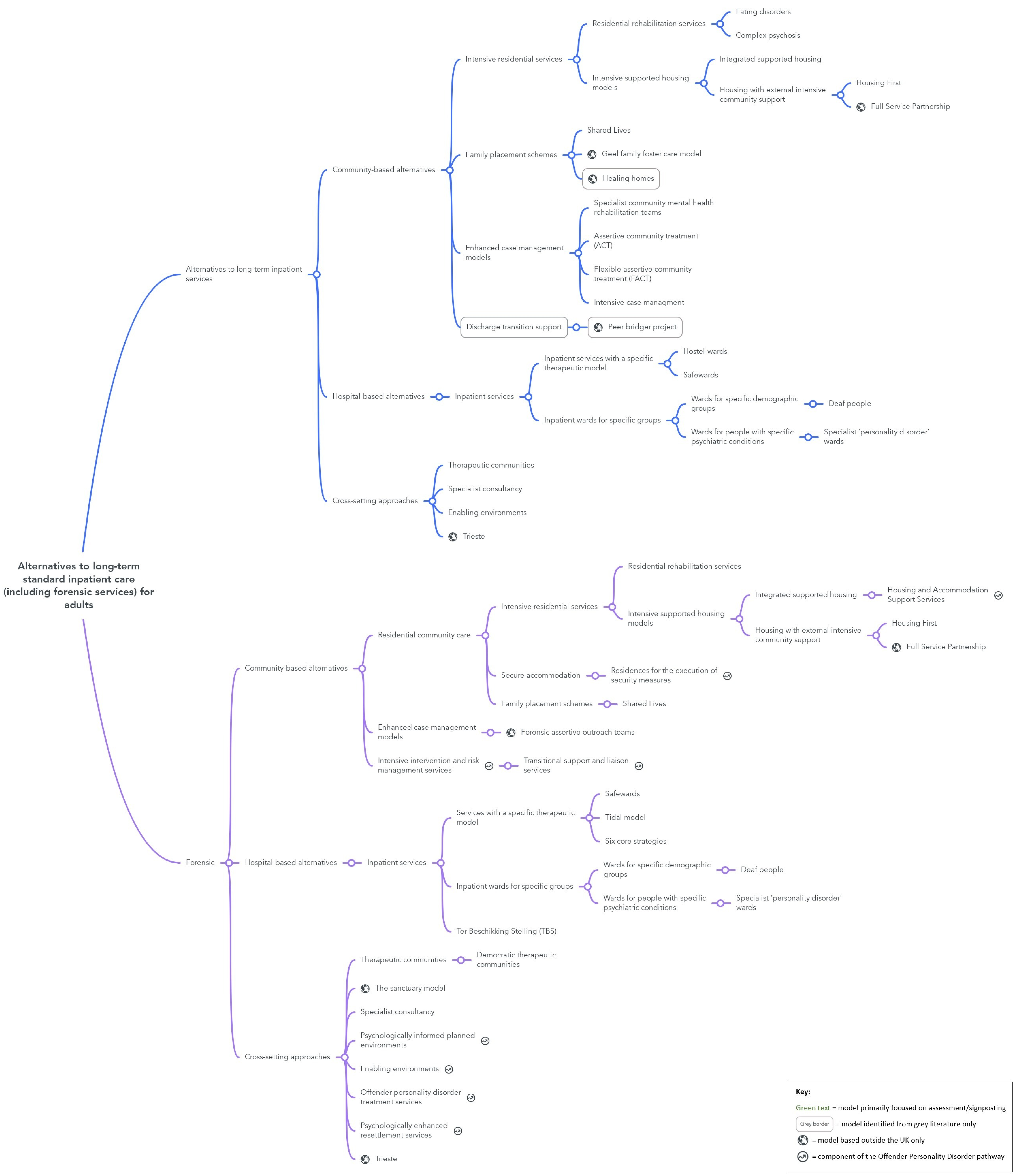

### Additional File 6

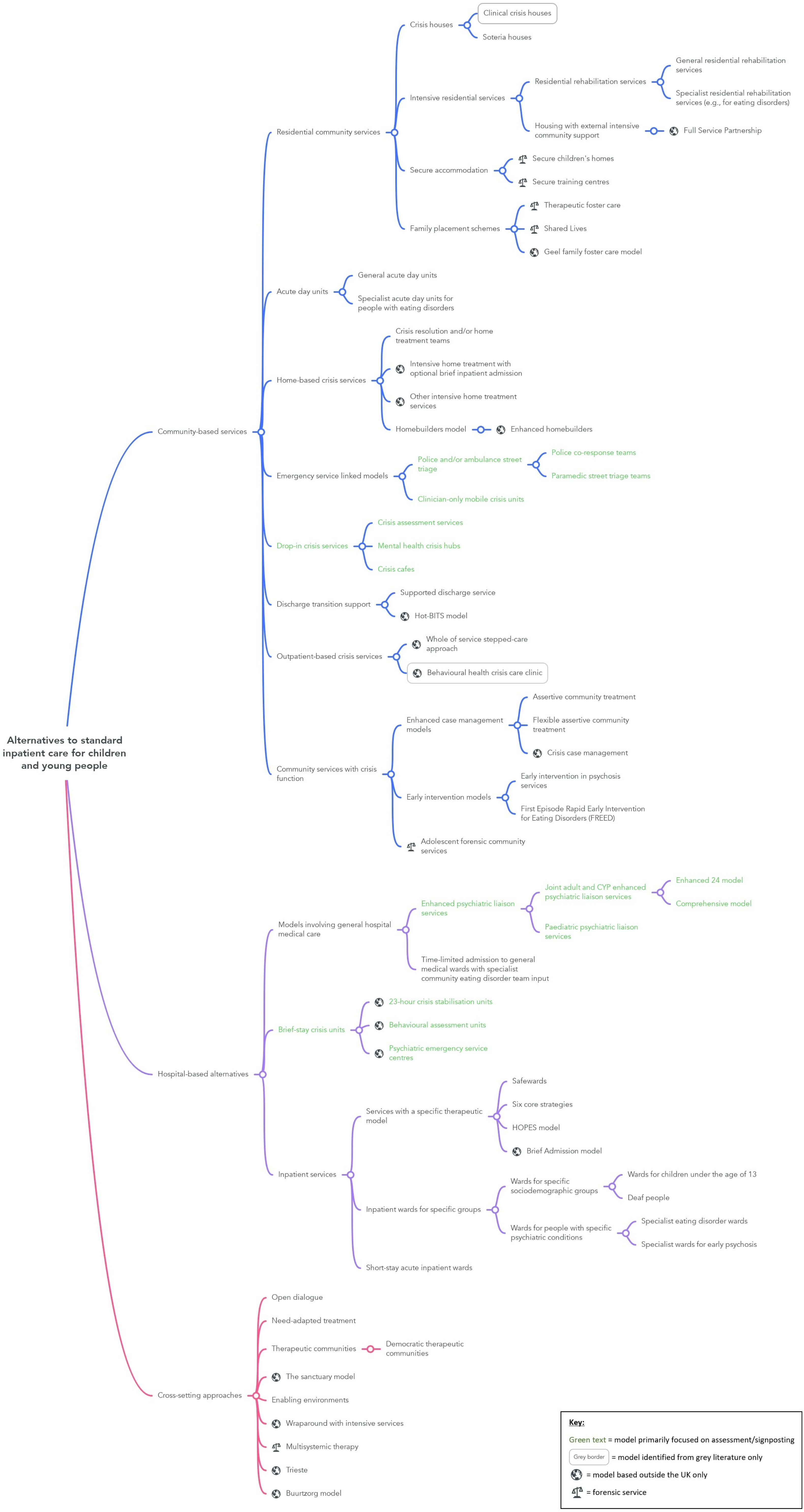
